# Identifying genetic differences between bipolar disorder and major depression through multiple GWAS

**DOI:** 10.1101/2024.01.29.24301816

**Authors:** Georgia Panagiotaropoulou, Kajsa-Lotta Georgii Hellberg, Jonathan R. I. Coleman, Darsol Seok, Janos Kalman, the Bipolar Disorder Working Group of the Psychiatric Genetics Consortium, the Major Depressive Disorder Working Group of the Psychiatric Genetics Consortium, the iPSYCH Study Consortium, Philip B. Mitchell, Peter R. Schofield, Andreas J. Forstner, Michael Bauer, Laura J. Scott, Carlos N. Pato, Michele T. Pato, Qingqin S. Li, George Kirov, Mikael Landén, Lina Jonsson, Bertram Müller-Myhsok, Jordan W. Smoller, Elisabeth B. Binder, Tanja M. Brückl, Darina Czamara, Sandra Van der Auwera, Hans J. Grabe, Georg Homuth, Carsten O. Schmidt, James B. Potash, Raymond J. DePaulo, Fernando S. Goes, Dean F. MacKinnon, Francis M. Mondimore, Myrna M. Weissman, Jianxin Shi, Mark A. Frye, Joanna M. Biernacka, Andreas Reif, Stephanie H. Witt, René R. Kahn, Marco M. Boks, Michael J. Owen, Katherine Gordon-Smith, Brittany L. Mitchell, Nicholas G. Martin, Sarah E. Medland, Lisa Jones, James A. Knowles, Douglas F. Levinson, Michael C. O’Donovan, Cathryn M. Lewis, Gerome Breen, Thomas Werge, Andrew J. Schork, Roel Ophoff, Stephan Ripke, Loes Olde Loohuis

## Abstract

**Background:** Accurate diagnosis of bipolar disorder (BD) is difficult in clinical practice, with an average delay between symptom onset and diagnosis of about 7 years. A key reason is that the first manic episode is often preceded by a depressive one, making it difficult to distinguish BD from unipolar major depressive disorder (MDD).

**Aims:** Here, we use genome-wide association analyses (GWAS) to identify differential genetic factors and to develop predictors based on polygenic risk scores that may aid early differential diagnosis.

**Methods:** Based on individual genotypes from case-control cohorts of BD and MDD shared through the Psychiatric Genomics Consortium, we compile case-case-control cohorts, applying a careful merging and quality control procedure. In a resulting cohort of 51,149 individuals (15,532 BD cases, 12,920 MDD cases and 22,697 controls), we perform a variety of GWAS and polygenic risk scores (PRS) analyses.

**Results:** While our GWAS is not well-powered to identify genome-wide significant loci, we find significant SNP-heritability and demonstrate the ability of the resulting PRS to distinguish BD from MDD, including BD cases with depressive onset. We replicate our PRS findings, but not signals of individual loci in an independent Danish cohort (iPSYCH 2015 case-cohort study, N=25,966). We observe strong genetic correlation between our case-case GWAS and that of case-control BD.

**Conclusions:** We find that MDD and BD, including BD with a depressive onset, are genetically distinct. Further, our findings support the hypothesis that Controls – MDD — BD primarily lie on a continuum of genetic risk. Future studies with larger and richer samples will likely yield a better understanding of these findings and enable the development of better genetic predictors distinguishing BD and, importantly, BD with depressive onset from MDD.

## 1. Introduction

Bipolar disorder (BD) affects more than 1% of the world’s population irrespective of nationality, ethnic origin, or socioeconomic status^1,2^. In WHO’s World Mental Health surveys, BD was ranked as the illness with the second greatest effect on days out of role^3,4^. Accurate diagnosis of BD is difficult in clinical practice: mean delay between symptom onset and diagnosis is around 7 years^5^. One of the main reasons for this delay is that onset is often characterized by a depressive episode and until the onset of mania it is difficult to distinguish these BD patients from patients with unipolar major depressive disorder (MDD) ^6,7,8,9,10,11,12^. For example, in studies that have followed-up patients with an initial MDD diagnosis, approximately between 10-20% demonstrate conversion to BD over follow-up periods of about 5-10 years^13,14^. The misdiagnosis of BD can have significant detrimental consequences, including prescription of antidepressants in the absence of mood-stabilizing drugs, which can lead to mania^15^, poor clinical outcomes and ultimately high healthcare costs. Family-based studies^8,16^ and our recent GWAS ^17^ demonstrate independent patterns of inheritance for mania and depression and initial presentation of bipolar disorder^10^. Several recent studies identified BD genetic liability as a predictor of conversion to BD^18,19^. Together, these findings suggest that scrutinizing the genetic relationship between these two core phenotypes will be valuable in understanding risk for BD. While several summary-statistics-based genetic studies have evaluated genetic similarities and differences between BD and MDD^19^, no study has yet been performed directly assessing the genetic differences between these two phenotypes using a systematic approach of combining individual-level genetic data from different cohorts.

Here, we aim to characterize genetic differences between BD patients and patients with MDD using data from the Psychiatric Genomics Consortium (PGC total N=68,612 participants)^20,21^ with a replication in the iPSYCH case-control study (total N=25,966)^22,23^. In a follow up analysis, we focus specifically on patients with a first onset of depression, *depression-first BD,* who are most difficult to differentiate from MDD in clinical settings.

## 2. Methods

### 2.1. Sample Description

Our analyses are based on 17,673 BD and 14,346 MDD cases of European ancestry from Europe, North America and Australia from the Psychiatric Genomics Consortium (PGC) BD and MDD Working Group, which comprised our discovery data^20,21^. For a list of included cohorts, their sample sizes and case control breakdown, see the Supplementary Material (Supp. Tables 1, 2). The individual studies were approved by the respective local ethics committees and all participants provided written informed consent.

**Table 1.** Replication results of PRS analysis using iPSYCH as the target cohort. Top panel: AUC and Nagelkerke’s R^2^ achieved by each model (i.e. null model - principal components only, full model - BDvsMDD GWAS and full model with combined predictor) for BD vs. MDD status classification; bottom panel: similar for BD-D vs. MDD status. Note: Nagelkerke R2 adjusted = (Nagelkerke R2 Full model) - (Nagelkerke R2 Null model)

|  |  | Null model (predicted by<br>10 PCs) | Full model (predicted by<br>BDvsMDD + 10 PCs) | Full model combined<br>predictor (predicted by<br>BDvsMDD + BD + MDD +<br>10 PCs) |
| --- | --- | --- | --- | --- |
| **BDvsMDD** | **AUC** | 0.563 | 0.578 | 0.587 |
|  | **Nagelkerke R2** | 0.99% | 1.39% | 1.83% |
|  | **Nk R2 adjusted** | - | 0.40% | 0.84% |
| **ConvertBDvsMDD_0m** | **AUC** | 0.547 | 0.562 | 0.578 |
|  | **Nagelkerke R2** | 0.33% | 0.65% | 1.02% |
|  | **Nk R2 adjusted** | - | 0.32% | 0.69% |

Additionally, summary statistics of GWAS based on ICD-10 secondary care contacts from national health registers^24,25^ for both disorders were provided for the iPSYCH case-cohort study^22,23^, which were used for replication. All individuals were born in Denmark between 1981 and 2008 and enrolled based on a secondary care contact recorded in national health registers for BD (ICD-10: F30-F31) or MDD (ICD-10: F32-F33) before 2016. Individuals with a schizophrenia (ICD-10: F20) diagnosis were excluded. For iPSYCH samples, retrieved from the Danish Neonatal Screening Biobank, parents were informed at the time of sampling and given the option to withdraw the sample from inclusion in research studies^22^.

Polarity at onset (PAO) was available for a subset of participants with a BD diagnosis in the PGC cohorts. For these patients, as in our previous study^17^, PAO was determined by selecting the earliest age between the onset of mania/hypomania and depression, or as provided by the cohorts. Patients for whom PAO was available were categorized into two subgroups: depression before mania/hypomania (depression-first), and mania before depression of a mixed onset (mania-first). The latter category includes both participants whose onset was marked by an episode with mixed features and participants who had their first manic and depressive episode within the same year. For the iPSYCH data, depression-first PAO was indirectly inferred based on the presence of a registered MDD contact prior to first registered BD contact.

### 2.2. Genotype data merge, quality control and imputation

All PGC cohorts in our analysis ascertained patients with a single main diagnosis; either MDD or BD. To perform direct case-case genetic analyses at the genotype level, a first step is to combine multiple independent cohorts into unified cohorts including both MDD and BD case participants. To do so, great care needs to be taken to avoid introducing population stratification and technical artifacts while combining distinct data sources. We developed and applied an iterative procedure for merging, quality control and imputation in Ricopili^26^, described in detail in the Supplementary Notes A section. We thereby compiled 13 grouped case-case cohorts including 15,532 BD cases and 12,920 MDD cases in total.

We created a similar set of 13 grouped cohorts, adding 40,160 control participants from the original merged cohorts, performing a similar quality control procedure. The resulting 13 pairs of case-control cohorts contained 14,513 BD cases vs. 22,697 controls and 12,259 MDD cases vs. 17,463 controls, after additional outlier and overlap exclusions.

We also leveraged available information about BD POA (manic episode first - BD-M or depressive episode first - BD-D) to compile 7 case-case cohorts with 2,597 depression- first BD cases (BD-D) and 9,217 matching MDD cases. For BD-M, the sample size was too small (1,300 cases) and the overall observed heritability did not meet the recommended significance criteria (z=2.45, P>0.01)^27^, so we have not included the BD-M-based stratification in further analyses.

### 2.3. Genome wide association analyses

To evaluate genetic differences between BD and MDD, we performed three primary GWAS analyses and one replication analysis:

#### 2.3.1. Genotype-based Case-Case GWAS Meta-analysis

To identify genetic risk factors differentiating BD and MDD, we first compare BD and MDD cases directly, similar to a previous comparison of schizophrenia to BD^28^. Specifically, we perform GWAS on each of the 13 grouped case-case cohorts based on dosage genotypes, followed by standard inverse-SE weighted meta-analysis across all grouped cohorts, whereby individuals with BD were coded as cases, MDD cases as controls. The first 20 principal components were used as covariates. We refer to this primary GWAS analyses as *BDvsMDD GWAS*. We repeat this analysis using only depression-first BD cases and matched MDD cases (7 case-case cohorts) and refer to it as *BD-DvsMDD GWAS*.

#### 2.3.2. Meta-regression analysis

For this second GWAS analysis we introduce control individuals and aim to identify genetic differences between BD and MDD *relative to controls.* To do so, for each of the 13 cohorts, we first generated summary statistics for two GWAS: one of BD vs. controls and one of MDD vs. controls. Note that the controls for each group are split between BD and MDD cases proportionally (see previous section). We then used a meta-regression approach to model the effect size of each SNP as a function of a single fixed covariate: a binary indicator of phenotype (BD or MDD, see also Supp. Notes B). This GWAS is referred to as *MetaRegr GWAS*.

We also performed separate random effects meta-analyses of the BD and MDD GWAS summary statistics to evaluate which phenotype appeared to have more heterogeneity in SNP effect sizes using the respective meta-regression r^2^ estimates.

#### 2.3.3. CC-GWAS

We also performed a GWAS based on the CC-GWAS method^29^, using BD vs. controls and MDD vs. controls summary statistics. For this, we compiled a version of our grouped cohorts based on a set of completely overlapping controls, as CC-GWAS covariance matrix estimation benefits from control overlap. LD score regression ^30^ was used to calculate the set of parameters required as input by the method (see Supp. Table 4). In addition to applying a genome-wide threshold for p-value, CC-GWAS includes an “stress test” to determine whether a SNP is considered significant, accounting for any indication of differential tagging of a shared causal allele (i.e. SNPs with similar allele frequency for both disorders), arising from subtle ancestry differences in the input. We thus also filter our results accordingly, including hits which pass this additional filter.

#### 2.3.4 Reverse GWAS

In Coleman et al.^31^, summary statistics were used to identify loci with differential signals between the two disorders (“reverse-effect” analysis). We evaluated concordance between loci identified through this analysis and our results, by evaluating the genome-wide significant hits in the “reverse-effect” analysis (three in total) in our three GWAS.

#### 2.3.5. Replication analysis with iPSYCH

To replicate our findings from the BDvsMDD GWAS, we performed a similar case-case association analysis in the iPSYCH 2015 case-cohort study (2,524 BD cases and 23,442 MDD cases). GWAS was performed using Plink2 v2.00a2^32^ in two independent samples (iPSYCH-2012, N_BD=1,452, N_MDD=15,920 and additional iPSYCH-2015i, N_BD=1,072, N_MDD=7,522) and meta-analyzed.

For our onset analysis, we also utilize a constrained set of 976 individuals who had an MDD diagnosis registered on the same day or prior to their BD diagnosis (BD-D), against the set of 23,442 individuals with MDD diagnosis. To evaluate the degree of replication of LD independent index SNPs from our primary GWAS, we performed a sign test, grouping variants with p-value smaller than 1e-05, to determine whether the percentage of variants in the original analysis retaining their direction of effect in the replication analysis is significantly higher than chance.

### 2.4. Heritability and genetic correlation

For all GWAS, heritability and genetic correlations were estimated with LD score regression. In addition, we estimated genetic correlations between our GWAS and well-powered (SNP heritability z-score>5 and more than 10,000 cases) psychiatric GWAS made publicly available by the PGC (https://pgc.unc.edu/for-researchers/download-results/). The following traits were included: Schizophrenia (SCZ), ADHD, Cannabis Use Disorder (CUD), Alcohol Dependence (AD), Alcohol Use Disorder (AUD), Anorexia Nervosa (AN), Autism Spectrum Disorder (ASD), Post-traumatic Stress Disorder (PTSD). Since our analysis is currently limited to European ancestry, we used summary statistics limited to the European population subset.

### 2.5. Polygenic score analyses

To evaluate whether our GWAS can help distinguish between patients with MDD and those with BD on an individual level, we compute polygenic risk scores (PRS). We calculate leave-one-out (LOO) summary statistics based on our set of GWAS and use SBayesR^33^ to calculate polygenic scores for each of the 13 grouped cohorts respectively. We thus create a number of different polygenic predictors, including combinations of those using multiple regression. We report the area-under-curve (AUC) score as a metric for performance, as well as the percentage of variance explained, expressed in terms of Nagelkerke’s R^2^.

Specifically, we calculate polygenic scores based on summary statistics of 4 different GWAS: i) BDvsMDD GWAS, ii) BD vs. controls GWAS (BD GWAS), iii) MDD vs. controls GWAS (MDD GWAS) and iv) MetaRegr GWAS. We compare the ability of each of these scores, based on different GWAS designs, as well as a combination of (i), (ii) and (iii) (combined using multiple regression), to predict the target phenotype, namely to classify BD vs. MDD status.

To obtain within-cohort standard errors and calculate confidence intervals for the AUC, we bootstrap the process based on 100 samples for each cohort.

To compare classification performance across different predictors, we further performed paired (across cohorts) weighted t-tests, with weights based on the effective sample size of the target cohorts, to determine the statistical significance of the difference in performance between individual predictors and the combined predictor (CC+BD+MDD). Since using t-tests we do not rely on confidence intervals, these performance comparisons between predictors were based on the AUC values reported for each of our cohorts, and not the ones obtained via the bootstrapping process.

To further quantify the impact of sample size, we compared our predictors to the BD GWAS of the Psychiatric Genetics Consortium in ^34^. As each of our grouped cohorts contains multiple BD and MDD studies, it is an involved process to create LOO summary statistics while removing overlap; we therefore limit this comparison to one cohort (“grp5_neth”).

Since we are most interested in distinguishing BD patients with an onset of depression from those with unipolar MDD, we repeat the above analysis using BD-D vs. MDD cohorts as target datasets. Finally, we test the reproducibility of our PRS results on the iPSYCH cohort.

### 2.6. Polygenic risk scores based on other psychiatric traits

Using SBayesR, we also calculated polygenic scores based on public summary statistics for each of the psychiatric GWAS included in our genetic correlation analysis. We report mean weighted AUC calculated across our 13 cohorts.

## 3. Results

### 3.1. GWAS does not identify significant loci

Our GWAS results using our three different GWAS methods are summarized in Supp. Table 5, after visual inspection of region plots produced by Ricopili for reasonable LD patterns. Overall, we observe no genome-wide significant hits for BDvsMDD or meta-regression, while one locus passes the genome-wide threshold for CC-GWAS. While our primary GWAS (BDvsMDD) did not yield significant loci, we observed significant heritability (observed h2 = 0.23 (se 0.02), intercept 1.001 (se 0.01)). For the BD-D vs. MDD GWAS, we observed similar results (observed h2 = 0.18 (se 0.04), intercept 1.01 (0.01)). Our two secondary GWAS (meta-regression, CC-GWAS) were strongly correlated with BDvsMDD and with each other (rg 0.91-1, Figure 1a), but they were less well-powered than BDvsMDD (meta-regression: h2 = 0.05 (se 0.01) with intercept 0.96 (0.01), CC-GWAS: h2 = 0.17 (se 0.01) with intercept 0.98 (0.01)).

**Figure 1.**
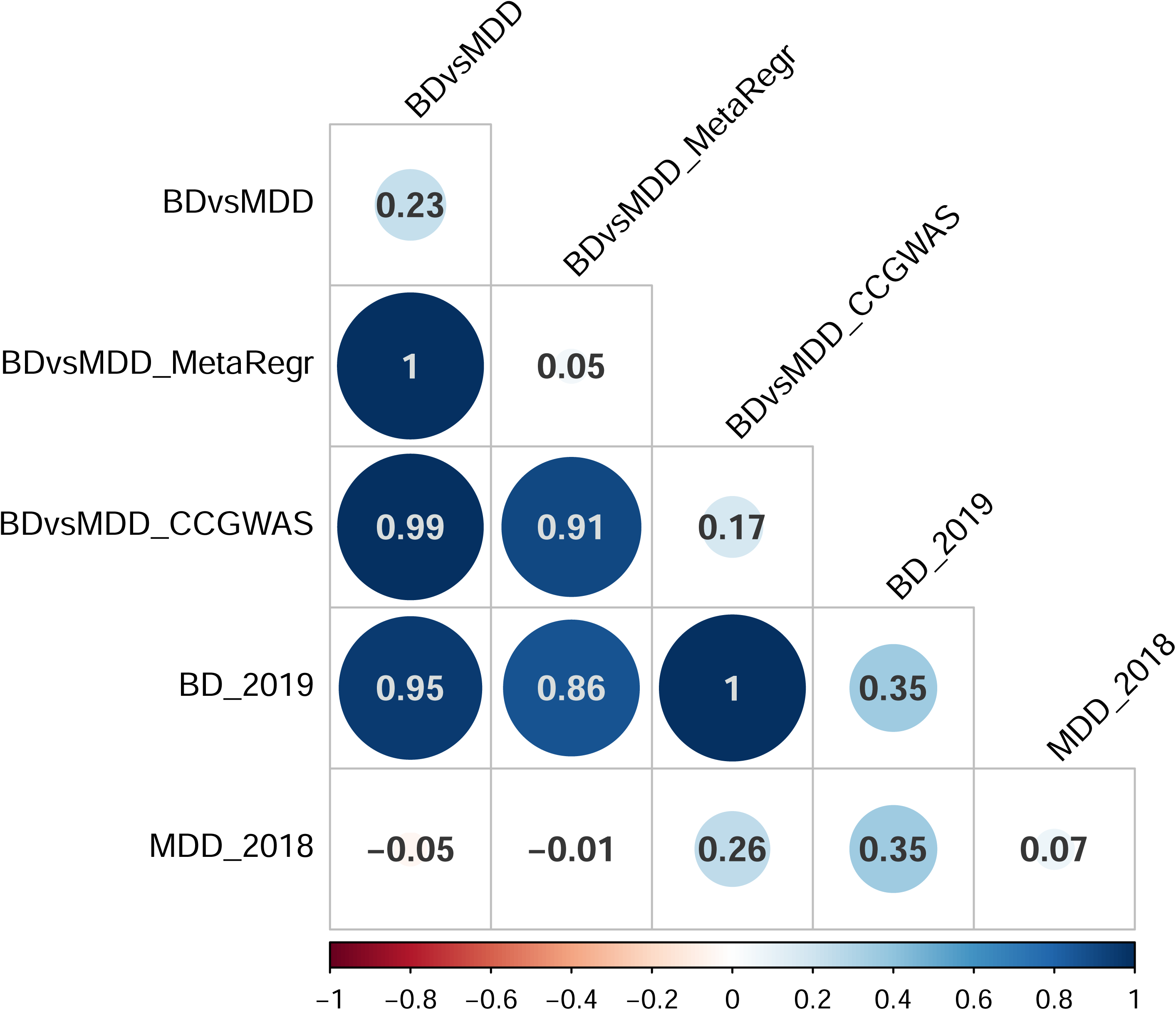
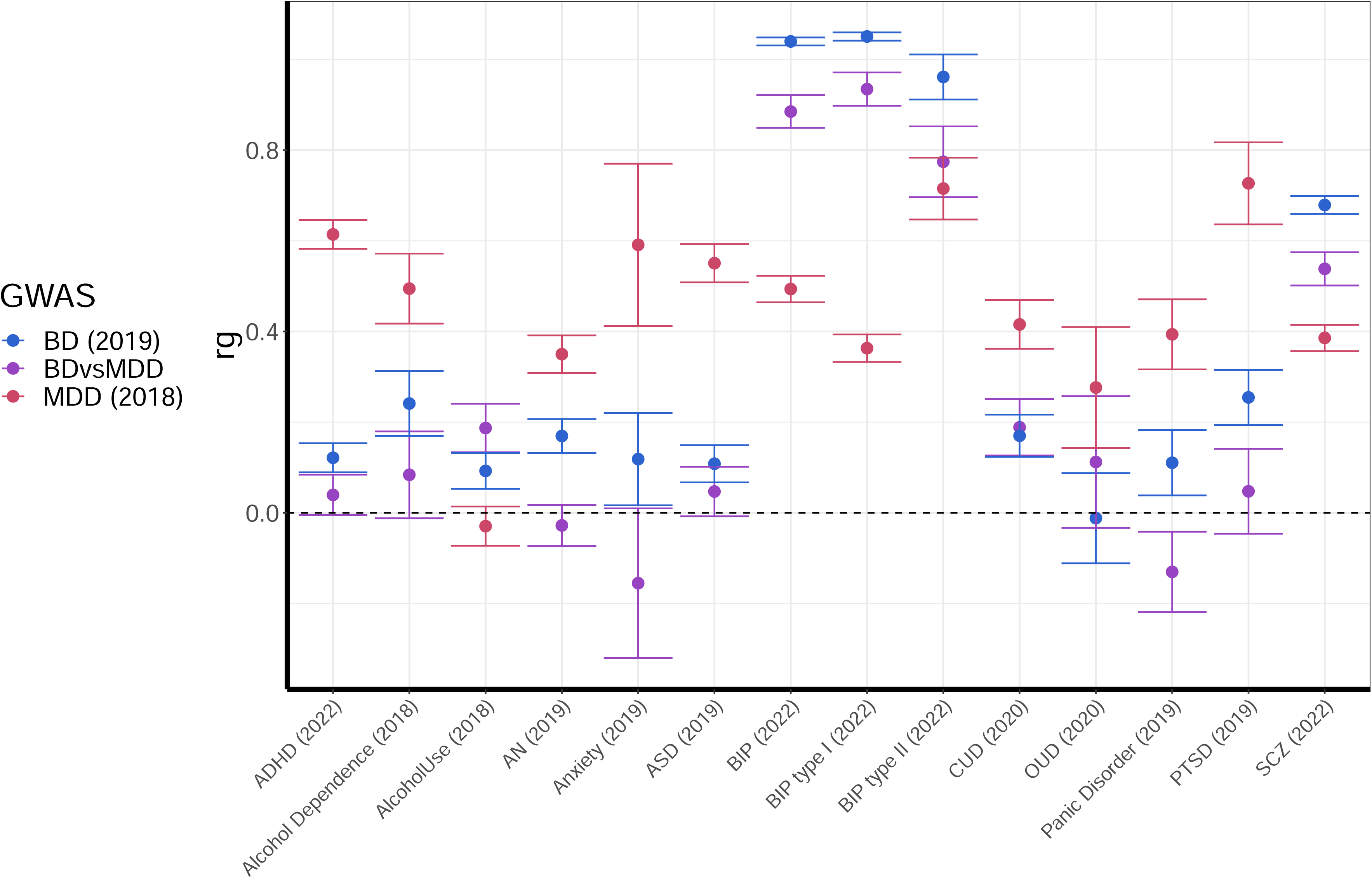
A) Genetic correlations between the different GWAS methods performed B) Genetic correlations between the case-case GWAS (BDvsMDD purple), our BD case-control GWAS (blue) and our MDD case-control GWAS (red) on the y-axis and GWAS of other psychiatric traits from the PGC on the x-axis.

A total of eight loci reached a suggestive p-value of less than 1e-06 (Supp. Table 5) in BDvsMDD, two of which, marked in bold face, fall within known BD loci ^34^. The Manhattan, quantile-quantile (Q-Q), region and region forest plots for this analysis as well as the corresponding Manhattan and QQ plots for the BP-D vs. MDD GWAS can be found in Supp. Figures 2a-b, 3a-b, 4a and 5. As seen in the region plots (Supp. Figure 4a), one of the loci (in chromosome 11) harbors two potentially independent signals.

Respectively, four loci reached suggestive genome-wide significance for the meta-regression analysis, none of which coincide with those of the BDvsMDD GWAS (Supp. Figures 2c, 3c and 4b). For CC-GWAS, we do not report suggestive loci, since we do not have differential tagging information for those.

For the single hit (rs174601 on chromosome 11, P=6.43e-09, with OR 0.99) identified through this analysis, we also report results on BDvsMDD, BD, MDD and meta-regression (Supp. Figures 2d, 3d and 4c). For both BDvsMDD and meta-regression we observe a similar effect P<1.0e-05 and a larger effect size (OR of 0.93 for BDvsMDD GWAS and 0.89 for meta-regression), while for BD this SNP is genome-wide significant with P = 8.0e-10 and maps onto a known BD locus, close to the FADS1 gene^34^. We observe a signal in the same direction for MDD, though the effect is not significant (P>0.1).

In none of the three different GWAS do we observe genetic signal (at P < 1e-04) for the three SNPs reported to differentiate BD and MDD in ^31^ (Supp. Table 6).

The phenotype-specific meta-regression analysis allowed us to compare effect size heterogeneity between MDD and BD cohorts. We observed slightly elevated effect size heterogeneity in MDD cohorts compared to BD, indicating that across all SNPs tested, MDD cohorts are slightly more heterogeneous; however, the observed difference is minimal (mean r^2^ values of 3.0e-02 for MDD vs. 2.6e-02 for BD, P<1.0e-16 paired t-test in all 6.9 million SNPs).

### 3.2. Heritability and genetic correlation indicates a strong correlation with PGC BD GWAS

We observe a strong genetic correlation between the BDvsMDD GWAS summary statistics and the GWAS of PGC BD: rg = 0.95 with BD^20^ (Figure 1a and b), primarily BD type I (Figure 1b n; Note that genetic correlation estimates above 1 between PGC analyses occur. These may be due to overlapping individuals in the studies involved.) The correlation between BDvsMDD GWAS and our BD GWAS, using only matched individuals, is also strong: rg = 0.88 (se 0.03). On the other hand, the correlation estimate with PGC MDD^21^ is negative rg = −0.05 (se 0.06), but the standard error overlaps with zero. The negative direction of effect is expected, given that MDD cases were coded as “controls” in our case-case analyses (where “cases” correspond to individuals with BD).

Genetic correlations with other psychiatric traits tracks are presented in Figure 1b, alongside BD and MDD (See also Supp. Table 7). Mostly, the observed genetic correlations follow an expected pattern that matches the observations above: When a trait is strongly correlated with BD, and less so with MDD (e.g., SCZ), the genetic correlation of BDvsMDD falls in between. When a trait is strongly correlated with MDD, and less so with BD (e.g., PTSD, ADHD), the genetic correlation of BDvsMDD is driven towards zero (or a negative correlation) due to the relative strength of the MDD signal. An exception to this “rule” is Alcohol Use, which is more strongly correlated with BDvsMDD (rg=0.19, se=0.05) than with PGC BD (rg=0.09, se=0.04), indicating that genetic risk factors for alcohol use could represent additional independent risk for conversion from MDD to BD.

### 3.3. Polygenic risk scores can distinguish between MDD and BD, including BD-D

Figure 2A shows the classification score in terms of AUC (see also Supp. Figure 6 for Nagelkerke’s R^2^) for all 13 grouped cohorts, for polygenic scores based on BDvsMDD GWAS (BDvsMDD), BD GWAS (BD), MDD GWAS (MDD) and a combination of these three predictors (BDvsMDD+BD+MDD). The mean AUC (over 100 bootstrapped samples per cohort), weighted by cohort sample size is 0.62 (2.29% adjusted Nagelkerke R^2^), 0.63 (R^2^ = 4%), 0.59 (R^2^ = 0.29%) and 0.64 (R^2^ = 4.56%) respectively. Similar results are shown on Figure 2B for depression-first BD, as discussed later. For all cohorts in both plots, it can be deduced from the standard error bars that the AUC is significantly higher than chance level (0.5) and also significantly higher than the bootstrapped model using principal components only (null model – AUC of 0.58), with the exception of the MDD; here, the confidence intervals overlap the null model (for AUC) or zero (for adjusted R^2^) in seven cohorts. However, using paired t-tests, weighted by effective sample size, we show that the weighted mean across all 13 cohorts is significantly higher than that of the covariates-only “null” model (see Supp. Table 8).

**Figure 2.**
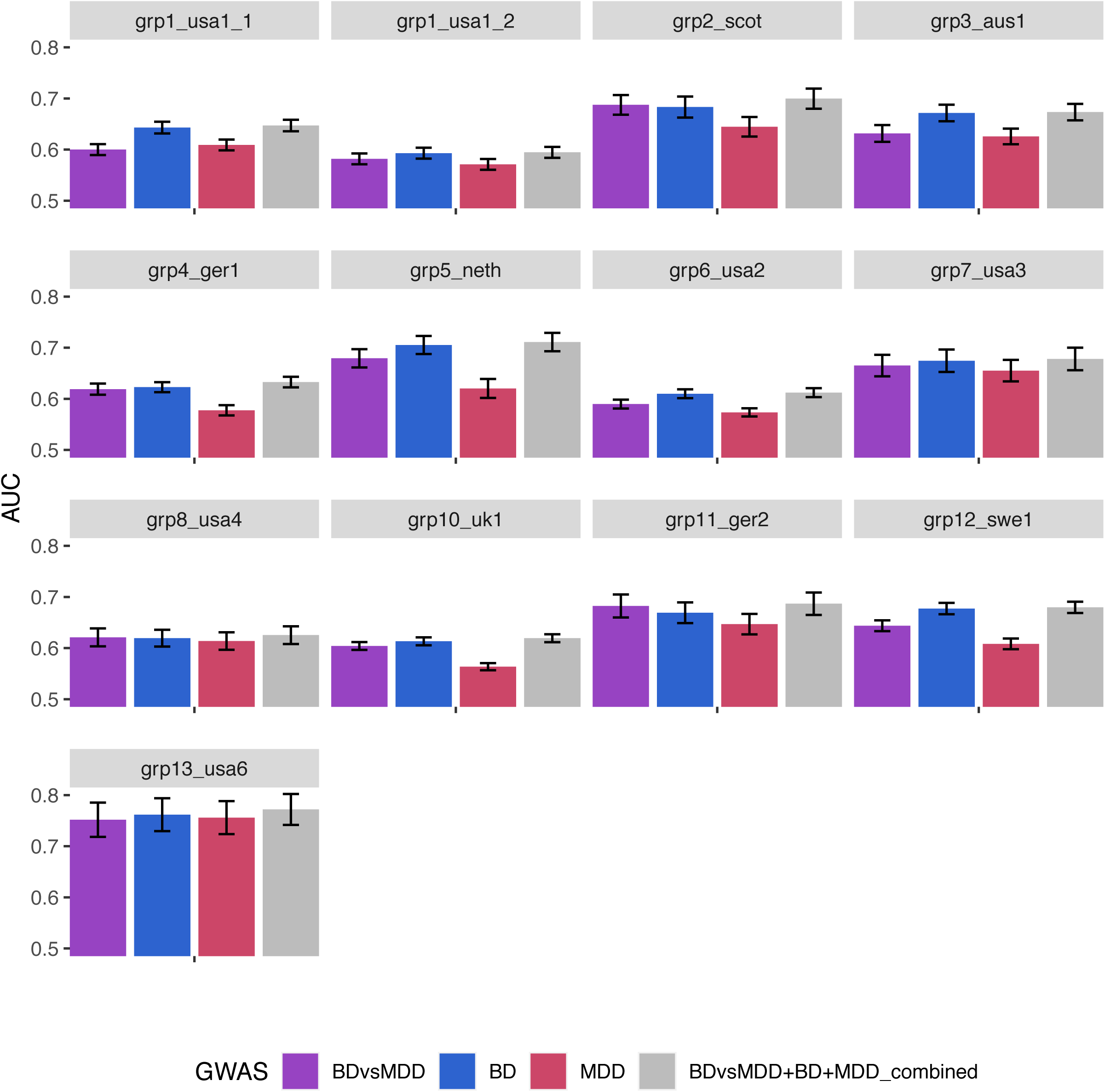
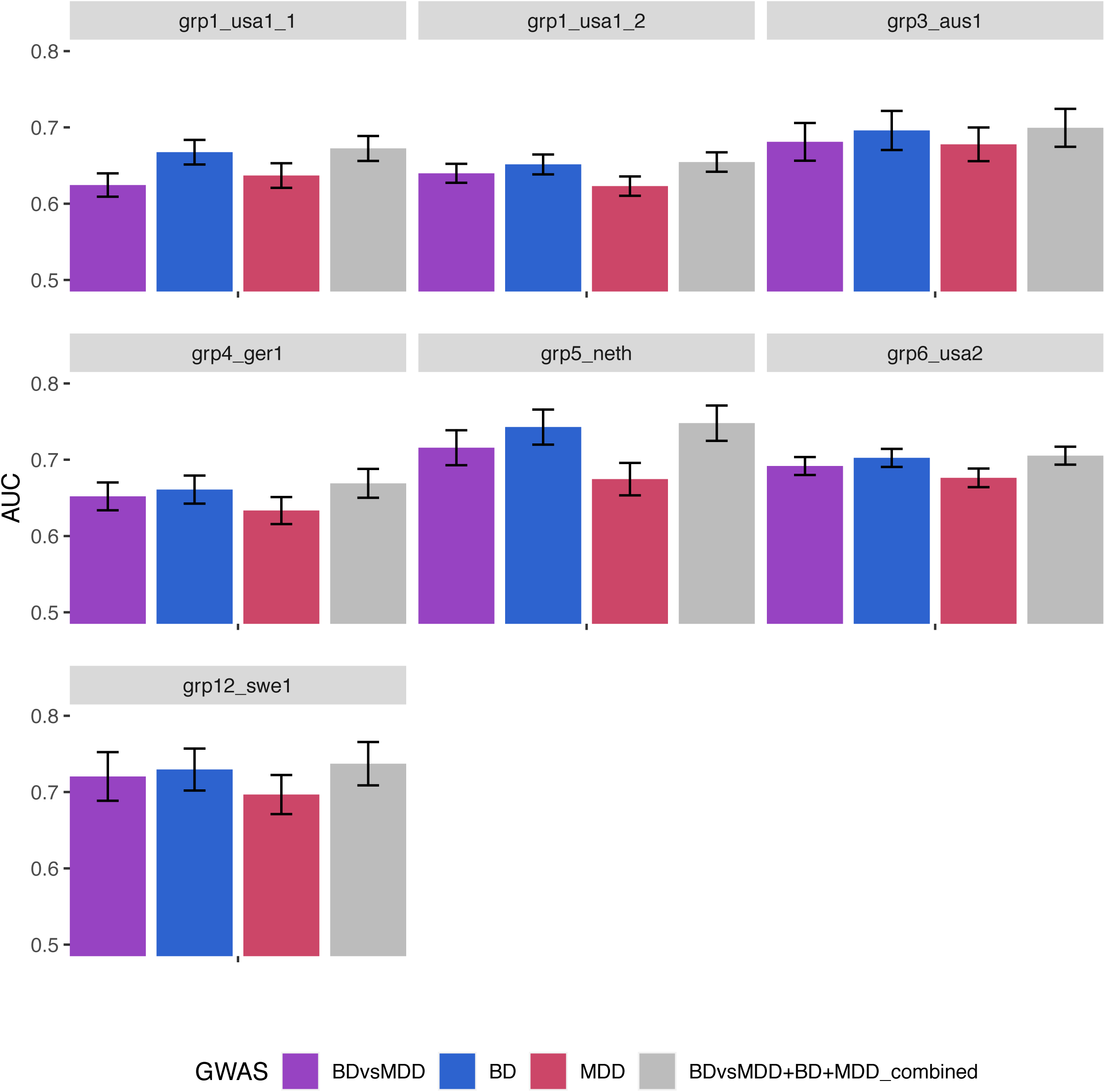
Ability of our GWAS to distinguish BD vs. MDD status in our cohorts: Area under the ROC curve (AUC) of PRS analysis using SBayesR for the BDvsMDD GWAS (A) and the BD with depressive onset (BD-D) vs. MDD GWAS (B) for all cohorts.

Interestingly, the BD predictor outperforms the predictor built on BDvsMDD cohorts. However, this is likely due to differences in sample size of the underlying GWAS: when we compare the BDvsMDD predictor to a version of the BD predictor based on a GWAS of equal sample size (BD-subN, see Supp. Notes C, Supp. Figure 7), the performance difference initially observed is no longer significant (p = 0.28 for equal sample size, paired weighted t-test).

Our comparison of the BD and BDvsMDD predictors to a more recent PGC BD collection^34^, including 41,917 cases and 371,549 controls, while attempted only for cohort “grp5_neth”, demonstrates the power advantage of the PGC BD GWAS-based predictor in classification performance (13.15% R^2^ for the PGC BD predictor, compared to 7.16% for the BDvsMDD predictor and 10.98% for our combined BDvsMDD+BD+MDD predictor, Supplementary Figure 8). However, combining our BDvsMDD predictor with the PGC BD one, yields even better performance (R^2^ = 14.81%), thus confirming the value of utilizing a predictor based on case-case GWAS.

Using a paired weighted t-test (one-tailed), we observed significantly increased performance of the combined predictor relative to each of the individual predictors: mean weighted AUC BDvsMDD = 0.60, BD = 0.62, MDD = 0.5 and combined = 0.63 (P-value of 3.5e-05 (BDvsMDD), 1.9e-03 (BD) and 6.5e-07 (MDD)).

To delineate the contribution of the signals attributable to each disorder, we further broke down the combined predictor to two-way combinations and found that the MDD signal contributes little orthogonal signal to the BDvsMDD+BD combination: mean AUC 0.62 for BDvsMDD+BD (compared to 0.63 for BDvsMDD+BD+MDD, as mentioned above, with P = 0.04, see Supplementary Figure 9A,B).

We next limited our analysis to the subgroup of patients with depression onset, testing the ability of BDvsMDD (and BD and MDD) PRS to distinguish between depression-first BD cases (BD-D) and MDD cases. We found that the classification accuracy is similar to that including all BD cohorts (Figure 2B and Supplementary Figure 9C,D). Our available sample size did not permit a similar analysis for manic-first episode BD (heritability z-score of 2.4).

Finally, Figure 3 shows the classification performance of all different psychiatric traits listed above (see Methods) with respect to differentiating between BD and MDD cases. Only SCZ is able to provide substantial differentiation between BD and MDD, comparable to our BDvsMDD GWAS (AUC = 0.61, se = 0.02), while for the rest of the available psychiatric traits, the performance is very poor.

**Figure 3.**
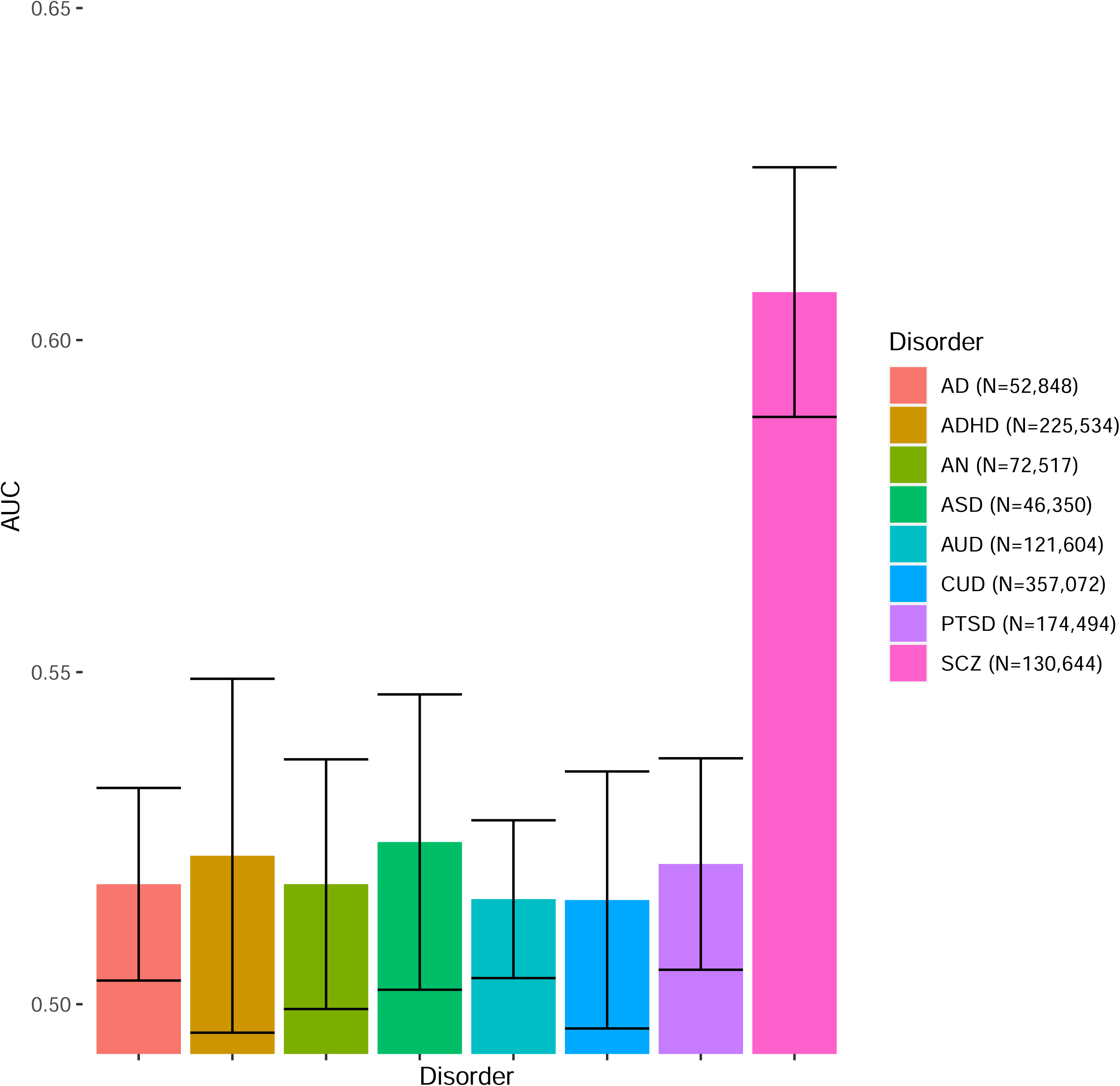
Ability of different psychiatric traits from the PGC to classify BD vs. MDD status in our cohorts. Mean AUC weighted by cohort effective sample size is reported.

### 3.4. Replication with iPSYCH

#### Sign tests

We tested 39 independent SNPs (P-value < 1.0e-05) from BDvsMDD, of which 22 (56%) had the same direction of effect in discovery and replication samples, indicating an accumulation of the same direction of effect in our replication sample, though this test does not reach nominal significance. We observe minimal SNP heritability of BDvsMDD in the iPSYCH cohort (h2=0.02 (se = 0.02), with intercept 1.003 (0.01)), which may account, in part, for this lack of replication.

#### Polygenic risk scoring

Polygenic scores based on our full PGC BDvsMDD GWAS, calculated using SBayesR, yielded an AUC of 0.62 and an incremental Nagelkerke R^2^ score of 0.40% on iPSYCH, after adjusting for population covariates in the regression model. Although it displays limited power, the PRS predictor is highly significant (P<1.0e-16), and an ANOVA between the full PRS model against the null model using covariates only is significant (P=1.9e-12), confirming the additional classification accuracy conferred by the PRS predictor.

Using our combined predictor in a multiple regression setting yields improved results, with an AUC of 0.63 and adjusted Nagelkerke R^2^ of 0.83%. After examination of the individual predictors, we see that the BD predictor has the strongest contribution (P=3.3e-07), while the BDvsMDD and MDD predictors are not statistically significant in the presence of the BD predictor (P>0.1). As before, the full model using BD, MDD and BDvsMDD outperforms the null model using only covariates (ANOVA, P<1.0e-16, also see Table 1) and the model outperforms using the BDvsMDD predictor only (ANOVA, P=2.8e-12).

Constrained to individuals with an MDD diagnosis prior to BD diagnosis, our models have similar classification performance, with an AUC of 0.61 and adjusted Nagelkerke R^2^ of 0.32% for the BDvsMDD and an AUC of 0.62 and adjusted Nagelkerke R^2^ of 0.69% for the combined predictor.

## 4. Discussion

With the goal of identifying genetic differences between MDD and BD, we performed three GWAS: a direct comparison between cases of both disorders, a meta-regression testing whether effect sizes differ between BD vs. Controls and MDD vs. Controls across cohorts, and CC-GWAS using case-control summary statistics.

While we found that MDD and BD are genetically distinct, with an estimated heritability of 23% on the observed scale in the direct comparison GWAS (5% by meta-regression, and 17% by CC-GWAS), our primary GWAS yielded no genome-wide significant loci. This lack of signal is likely due to a lack of power. While we were able to include 76% of PGC participants available for these analyses, with the resulting sample sizes they are still relatively underpowered to yield genome-wide significant hits for psychiatric traits, given their polygenicity and sizes of underlying effects, among other factors^35^. Compared to our primary analysis, both secondary GWAS (meta-regression and CC-GWAS), require additional power beyond a standard inverse-weighted meta-analysis, for different reasons. The meta-regression framework benefits from the addition of control individuals, but as a mixed effect model also requires more power to fit additional parameters. On the other hand, CC-GWAS relies solely on summary statistics, which can facilitate access to larger sample sizes as they become available. However, using our data, we obtained one genome-wide significant hit with CC-GWAS, which has support from both BDvsMDD and meta-regression, as well as BD GWAS. The lack of signal in MDD underlines the BD-specificity of this locus.

Somewhat surprisingly, we observed that the BDvsMDD GWAS was strongly correlated with BD GWAS (ranging between 0.88-0.95). Genetic correlations between BDvsMDD and other psychiatric traits are consistent with this observation.

Our leave-one-out polygenic risk scoring analysis confirms the ability of our BDvsMDD GWAS to differentiate between BD and MDD status, which is enhanced when adding multiple predictors from the corresponding case-control GWAS in a multiple regression setting (combined BDvsMDD+BD+MDD predictor). Although it is possible that this is attributable to the increased effective sample size rather than orthogonal signal, we found that the BD and MDD predictors (of similar sample size) contribute differently.

Consistent with the observation that the BDvsMDD GWAS has a high genetic correlation with BD, we found that including the MDD predictor (based on the MDDvsControls GWAS) did not add substantial orthogonal information over and above the BDvsMDD+BD predictors.

Moreover, we observed that our BDvsMDD predictor, which relies on careful matching of cases across cohorts originally designed for case-control studies, does not outperform our BD GWAS predictor, even when the latter, originally of larger sample size, is subsampled for comparison. We did not observe a similar effect for MDD, for which the training GWAS sample size is also larger than the BDvsMDD GWAS: the MDD GWAS was a worse predictor than either the BD GWAS or the BDvsMDD GWAS alone.

Our BDvsMDD and combined predictors had lower performance than a predictor built on the latest BD GWAS^34^, which is derived from a much larger sample size, although this comparison was limited to one dataset due to extensive sample overlap between the GWAS being compared. In this dataset, the BD GWAS does not saturate classification accuracy: using our BDvsMDD in conjunction with the well-powered latest BD GWAS from the PGC yielded the highest accuracy for the dataset tested. This is expected, since the overall variance explained by PRS is not yet close to the observed heritability.

Finally, we tested the ability of PRS to differentiate between patients with unipolar depression and BD patients who are most difficult to diagnose: those with a depressive onset. Given that depression-first BD cases have stronger depressive features than those with a manic POA^36,17^, one may hypothesize that the ability of PRS to distinguish between depression-first BD cases and MDD cases is lower than that including all BD cases. To the contrary, we observe that the classification accuracy of PRS is statistically indistinguishable to that including all BD patients, in all cohorts. This finding is encouraging, as it opens the possibility of future genetic studies to aid in precision psychiatry efforts, including the differential diagnosis of mood disorders.

Our replication effort in iPSYCH did not show strong signals of replication. This may be due to lack of power, but also may be impacted by the differences in ascertainment strategies. Patients in the iPSYCH samples are ascertained in secondary care hospitals where only ∼15% of MDD cases in Denmark are treated ^37^, which may mean the PGC MDD cases, comprising our discovery sample, may be less representative of them. This is consistent with previous work^38^, showing that the genetic correlation between iPSYCH-PGC for MDD is lower than for BD and that the MDD-BD cross-disorder genetic correlation is higher in iPSYCH than in prior PGC studies, potentially limiting the power to identify discriminating genetic signals. In the PGC data available to us, 83% of BD case participants have BD-I, indicating a selection for severity, whereas this number is not known in iPSYCH. Despite these differences, polygenic risk scores effects were replicated in iPSYCH.

Taken together, our results support the hypothesis that Controls – MDD — BD primarily lie on a continuum of genetic risk, with little specific MDD vs. BD signal detectable at the current sample sizes.

However, larger sample sizes are needed to further investigate the similarities and differences between BD and MDD. Since disease prevalence and heritability differ between BD and MDD (BD has higher heritability and lower prevalence compared to MDD), relatively larger sample sizes are needed to detect MDD-specific signals^39^. Our genetic correlation and PRS results suggest that additional orthogonal signals are yet to be identified.

In addition to larger sample sizes, future studies with richer phenotypic information and multi-diagnostic cohorts, as well as more direct case-case analyses, will likely yield a better understanding of these findings and enable the development of better genetic predictors distinguishing BD from MDD and more specifically depression-first BD from MDD.

Here, leveraging the dataset currently available, we provide an approach to carefully match and compile case-case-control cohorts from existing case-control cohorts, which enable more comprehensive analyses of underlying genetic architecture such as the one provided here. Specifically, the collection of 13 case-case-control cohorts compiled here will be a valuable resource for the research community in psychiatric genomics. Information on accessing these data from studies shared with the PGC will be available on the PGC website. Summary statistics data from case-case GWAS analysis will also become available upon publication.

## Data Availability

Information on accessing merged genotype data from studies shared with the PGC will be available on the PGC website (https://pgc.unc.edu/). Summary statistics from case-case GWAS will be made available on the PGC website upon publication.

## List of Supplementary Figures and Tables

Supplementary Figure 1. PCA plots for each cohort, showing PCA1 (x-axis) against PCA2 (y-axis), corresponding to the case-case PCA (A) and the case-control (B) analysis.

Supplementary Figure 2. Manhattan plots for the BDvsMDD GWAS (A), the BD-DvsMDD GWAS (B), the meta-regression GWAS (C) and the CC-GWAS (D).

Supplementary Figure 3. Quantile-quantile plots for the BD vs. MDD GWAS (A), the BD-DvsMDD GWAS (B), the meta-regression GWAS (C) and the CC-GWAS (D).

Supplementary Figure 4. Region plots for the BD vs. MDD GWAS (A), the meta-regression GWAS (B) and the CC- GWAS (C).

Supplementary Figure 5. Region forest plots for the BD vs. MDD GWAS.

Supplementary Figure 6. Nagelkerke’s R^2^ of PRS analysis using SBayesR for the BD vs. MDD GWAS (A) and the BD-D vs. MDD GWAS (B) for all cohorts.

Supplementary Figure 7. Comparison of classification accuracy between the PRS predictors based on the BD vs. MDD GWAS (BDvsMDD - red), the BD GWAS (BD - dark blue) and the BD GWAS with its sample size made equal to the BD vs. MDD GWAS (BD-subN, light blue). A) AUC with the BD vs. MDD cohorts as target, B) Ng R^2^ with the BD vs. MDD cohorts as target, C) AUC with the BD-D vs. MDD cohorts as target, D) Ng R^2^ with the BD-D vs. MDD cohorts as target.

Supplementary Figure 8. Comparison of PRS predictors based on our BD vs. MDD GWAS (blue), our combined predictor (magenta), the latest PGC BD GWAS (orange), and a predictor based on the combination of the two predictors based on our BD vs. MDD GWAS and the PGC BD GWAS (yellow). AUC with cohort “grp5_neth” as target is reported.

Supplementary Table 1. Merging and quality control results for case-case cohorts: constituent case-control cohorts for each of the 13 grouped case-case cohorts are reported, together with pre- and post-QC number of cases for each disorder and number of SNP.

Supplementary Table 2. Description of our quality control procedure with flags and corresponding values.

Supplementary Table 3. Summary of results introducing controls to the 13 grouped cohorts: Post-QC number of cases for both disorders as well as control individuals, inflation factor lambda, as well as number of post-QC SNPs are reported.

Supplementary Table 4. Full list of CC-GWAS input parameters used. Heritability estimates were obtained from LDSC.

Supplementary Table 5. List of genome-wide significant hits (P<5×10e-08) and suggestive hits (P<1×10e-6) for all three different GWAS methods: case-case BD vs. MDD, meta-regression and CC-GWAS. For the CC-GWAS hit, the corresponding statistics for other GWAS are reported as well.

Supplementary Table 6. List of hits from the “reverse-GWAS” analysis from Coleman et al. 2020: results from our case-case BD vs. MDD are reported.

Supplementary Table 7. List of genetic correlations between our GWAS and GWAS of other psychiatric traits from the PGC.

Supplementary Table 8. Paired t-test comparing the classification accuracy of models based on our PRS predictors against the null model based on principal components only.

## Supplementary Notes

### A. Merging, Quality Control and Imputation

Using Ricopili^26^, we developed and applied the following stringent and iterative procedure to match cohorts based on country and genome chip and perform quality control. We first evaluated potential merges of cohorts with cases of both phenotypes (“case-case” cohorts, with BD coded as “cases” and MDD coded as “controls”) based on matched genotyping platform and country. After performing the standard Ricopili QC pipeline, we evaluated each potential match based on nine criteria, to avoid technical artifacts, extremely unbalanced cohorts and inflation due to population structure, as well as to secure adequate number of good-quality variants: 1) the number of SNPs post-QC (fewer than 200,000 not considered) 2) the number of SNPs post-QC per platform (threshold is platform-dependent) 3) and 4) the number of MDD and BD cases (fewer than 100 not considered), 5) the number of participants lost relative to the total sample size (above 10% not considered) 6) number of individuals without phenotype (more than 10 not considered), 7) the ratio of cases failing sex-check (greater than 2.5% not considered) 8) lambda of post-QC GWAS without covariates (greater than 1.2 not considered) 9) number of genome-wide significant SNPs (not considered if any, given the small sample size of individual cohorts). Acceptable values for these criteria on all cohort-merges were obtained by iteratively excluding variants or individuals, while as few cohorts as possible were dropped altogether. We were able to match about 63% of BD cases and 65% of MDD cases of the total PGC collection, grouped into 13 case-case cohorts, and included 15,532 BD cases with 12,920 MDD cases from 22 and 19 cohorts respectively (Suppl. Tables 1, 2).

Principal component analysis of all the final merged cohorts as well as the final QC criteria can be found in Supp. Figures 1a, 1b and Supp. Tables 2 and 3.

Consistent with prior PGC analyses, QC was followed by imputation to the European subset of the Haplotype Reference Consortium (HRC) reference panel^40^. Specifically, genotype imputation was performed using the pre-phasing/imputation stepwise approach implemented in EAGLE / MINIMAC3 (with a variable chunk size of 132 genomic chunks and default parameters). The imputation reference set consisted of 54,330 phased haplotypes with 36,678,882 variants from the publically available HRC reference. For subsequent analysis we imposed a filter of INFO>0.8 on imputation quality score.

### B. Meta-regression framework

We use a meta-regression approach to model the effect size of each SNP (the coefficient of the SNP from the GWAS) as a function of a single fixed covariate: a binary indicator of phenotype (BD or MDD). Specifically, we used a random-effects meta regression, where group was treated as a random effect. In this random-effects model, a groups’s observed effect size 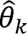 deviates from θ because of sampling error *ɛ_k_* and an additional error term ζ_k_ (varianceτ^2^) related to the variability of individual groups around 2 their true effect sizes. This regression uses maximally 26 estimates of effect size (two for each of the 13 groups). We analyzed only SNPs where there were GWAS summary statistics for at least 10 of the 13 groups.

The variable τ^2^ from the meta-regression estimates the variance of distribution of true SNP effect sizes after accounting for the fixed covariate (phenotype). We also performed a separate random effects meta-analyses of the BD and MDD GWAS summary statistics, obtaining two estimates of τ^2^, one for BD and one for MDD, to evaluate which phenotype appeared to have more heterogeneity in SNP effect sizes.

### C. Comparison between the BDvsMDD and BD predictors with equal sample sizes

Since the BD GWAS has a larger sample size than the BDvsMDD GWAS, due to the fact that control individuals (included in the former) outnumber MDD individuals (included in the latter), we also calculated scores based on a subsampled version of the former, whereby the number of controls included in the BD GWAS is downsampled to the same number as that of MDD cases in the BDvsMDD GWAS, to provide a fair comparison between the two.

Our comparison of BD and BDvsMDD with similar sample sizes is shown in Supp. Figure 3A (also see Supp. Figure 6C,D). In this scenario, the performance difference is no longer significant (p = 0.28 for equal sample size, paired weighted t-test), meaning that the BDvsMDD predictor still does not outperform the BD predictor, even after accounting for sample size.

### D. Full list of members of the Bipolar Disorder Working Group of the PGC, the Major Depressive Disorder Working Group of the PGC and the iPSYCH Consortium

#### Bipolar Disorder Working Group of the Psychiatric Genomics Consortium

Niamh Mullins^1,2,235†^, Andreas J. Forstner^3,4,5,235^, Kevin S. O’Connell^6,7^, Brandon Coombes^8^, Jonathan R. I. Coleman^9,10^, Zhen Qiao^11^, Thomas D. Als^12,13,14^, Tim B. Bigdeli^15,16^, Sigrid Børte^17,18,19^, Julien Bryois^20^, Alexander W. Charney^2^, Ole Kristian Drange^21,22^, Michael J. Gandal^23^, Saskia P. Hagenaars^9,10^, Masashi Ikeda^24^, Nolan Kamitaki^25,26^, Minsoo Kim^23^, Kristi Krebs^27^, Georgia Panagiotaropoulou^28^, Brian M. Schilder^1,29,30,31^, Laura G. Sloofman^1^, Stacy Steinberg^32^, Vassily Trubetskoy^28^, Bendik S. Winsvold^19,33^, Hong-Hee Won^34^, Liliya Abramova^35^, Kristina Adorjan^36,37^, Esben Agerbo^14,38,39^, Mariam Al Eissa^40^, Diego Albani^41^, Ney Alliey-Rodriguez^42,43^, Adebayo Anjorin^44^, Verneri Antilla^45^, Anastasia Antoniou^46^, Swapnil Awasthi^28^, Ji Hyun Baek^47^, Marie Bækvad-Hansen^14,48^, Nicholas Bass^40^, Michael Bauer^49^, Eva C. Beins^3^, Sarah E. Bergen^20^, Armin Birner^50^, Carsten Bøcker Pedersen^14,38,39^, Erlend Bøen^51^, Marco P. Boks^52^, Rosa Bosch^53,54,55,56^, Murielle Brum^57^, Ben M. Brumpton^19^, Nathalie Brunkhorst-Kanaan^57^, Monika Budde^36^, Jonas Bybjerg-Grauholm^14,48^, William Byerley^58^, Murray Cairns^59^, Miquel Casas^53,54,55,56^, Pablo Cervantes^60^, Toni-Kim Clarke^61^, Cristiana Cruceanu^60,62^, Alfredo Cuellar-Barboza^63,64^, Julie Cunningham^65^, David Curtis^66,67^, Piotr M. Czerski^68^, Anders M. Dale^69^, Nina Dalkner^50^, Friederike S. David^3^, Franziska Degenhardt^3,70^, Srdjan Djurovic^71,72^, Amanda L. Dobbyn^1,2^, Athanassios Douzenis^46^, Torbjørn Elvsåshagen^18,73,74^, Valentina Escott-Price^75^, I. Nicol Ferrier^76^, Alessia Fiorentino^40^, Tatiana M. Foroud^77^, Liz Forty^75^, Josef Frank^78^, Oleksandr Frei^6,18^, Nelson B. Freimer^23,79^, Louise Frisén^80^, Katrin Gade^36,81^, Julie Garnham^82^, Joel Gelernter^83,84,85^, Marianne Giørtz Pedersen^14,38,39^, Ian R. Gizer^86^, Scott D. Gordon^87^, Katherine Gordon-Smith^88^, Tiffany A. Greenwood^89^, Jakob Grove^12,13,14,90^, José Guzman-Parra^91^, Kyooseob Ha^92^, Magnus Haraldsson^93^, Martin Hautzinger^94^, Urs Heilbronner^36^, Dennis Hellgren^20^, Stefan Herms^3,95,96^, Per Hoffmann^3,95,96^, Peter A. Holmans^75^, Laura Huckins^1,2^, Stéphane Jamain^97,98^, Jessica S. Johnson^1,2^, Janos L. Kalman^36,37,99^, Yoichiro Kamatani^100,101^, James L. Kennedy^102,103,104,105^, Sarah Kittel-Schneider^57,106^, James A. Knowles^107,108^, Manolis Kogevinas^109^, Maria Koromina^110^, Thorsten M. Kranz^57^, Henry R. Kranzler^111,112^, Michiaki Kubo^113^, Ralph Kupka^114,115,116^, Steven A. Kushner^117^, Catharina Lavebratt^118,119^, Jacob Lawrence^120^, Markus Leber^121^, Heon-Jeong Lee^122^, Phil H. Lee^123^, Shawn E. Levy^124^, Catrin Lewis^75^, Calwing Liao^125,126^, Susanne Lucae^62^, Martin Lundberg^118,119^, Donald J. MacIntyre^127^, Sigurdur H. Magnusson^32^, Wolfgang Maier^128^, Adam Maihofer^89^, Dolores Malaspina^1,2^, Eirini Maratou^129^, Lina Martinsson^80^, Manuel Mattheisen^12,13,14,106,130^, Steven A. McCarroll^25,26^, Nathaniel W. McGregor^131^, Peter McGuffin^9^, James D. McKay^132^, Helena Medeiros^108^, Sarah E. Medland^87^, Vincent Millischer^118,119^, Grant W. Montgomery^11^, Jennifer L. Moran^25,133^, Derek W. Morris^134^, Thomas W. Mühleisen^4,95^, Niamh O’Brien^40^, Claire O’Donovan^82^, Loes M. Olde Loohuis^23,79^, Lilijana Oruc^135^, Sergi Papiol^36,37^, Antonio F. Pardiñas^75^, Amy Perry^88^, Andrea Pfennig^49^, Evgenia Porichi^46^, James B. Potash^136^, Digby Quested^137,138^, Towfique Raj^1,29,30,31^, Mark H. Rapaport^139^, J. Raymond DePaulo^136^, Eline J. Regeer^140^, John P. Rice^141^, Fabio Rivas^91^, Margarita Rivera^142,143^, Julian Roth^106^, Panos Roussos^1,2,29^, Douglas M. Ruderfer^144^, Cristina Sánchez-Mora^53,54,56,145^, Eva C. Schulte^36,37^, Fanny Senner^36,37^, Sally Sharp^40^, Paul D. Shilling^89^, Engilbert Sigurdsson^93,146^, Lea Sirignano^78^, Claire Slaney^82^, Olav B. Smeland^6,7^, Daniel J. Smith^147^, Janet L. Sobell^148^, Christine Søholm Hansen^14,48^, Maria Soler Artigas^53,54,56,145^, Anne T. Spijker^149^, Dan J. Stein^150^, John S. Strauss^102^, Beata Świątkowska^151^, Chikashi Terao^101^, Thorgeir E. Thorgeirsson^32^, Claudio Toma^152,153,154^, Paul Tooney^59^, Evangelia-Eirini Tsermpini^110^, Marquis P. Vawter^155^, Helmut Vedder^156^, James T. R. Walters^75^, Stephanie H. Witt^78^, Simon Xi^157^, Wei Xu^158^, Jessica Mei Kay Yang^75^, Allan H. Young^159,160^, Hannah Young^1^, Peter P. Zandi^136^, Hang Zhou^83,84^, Lea Zillich^78^, HUNT All-In Psychiatry*, Rolf Adolfsson^161^, Ingrid Agartz^51,130,162^, Martin Alda^82,163^, Lars Alfredsson^164^, Gulja Babadjanova^165^, Lena Backlund^118,119^, Bernhard T. Baune^166,167,168^, Frank Bellivier^169,170^, Susanne Bengesser^50^, Wade H. Berrettini^171^, Douglas H. R. Blackwood^61^, Michael Boehnke^172^, Anders D. Børglum^14,173,174^, Gerome Breen^9,10^, Vaughan J. Carr^175^, Stanley Catts^176^, Aiden Corvin^177^, Nicholas Craddock^75^, Udo Dannlowski^166^, Dimitris Dikeos^178^, Tõnu Esko^26,27,179,180^, Bruno Etain^169,170^, Panagiotis Ferentinos^9,46^, Mark Frye^64^, Janice M. Fullerton^152,153^, Micha Gawlik^106^, Elliot S. Gershon^42,181^, Fernando S. Goes^136^, Melissa J. Green^152,175^, Maria Grigoroiu-Serbanescu^182^, Joanna Hauser^68^, Frans Henskens^59^, Jan Hillert^80^, Kyung Sue Hong^47^, David M. Hougaard^14,48^, Christina M. Hultman^20^, Kristian Hveem^19,183^, Nakao Iwata^24^, Assen V. Jablensky^184^, Ian Jones^75^, Lisa A. Jones^88^, René S. Kahn^2,52^, John R. Kelsoe^89^, George Kirov^75^, Mikael Landén^20,185^, Marion Leboyer^97,98,186^, Cathryn M. Lewis^9,10,187^, Qingqin S. Li^188^, Jolanta Lissowska^189^, Christine Lochner^190^, Carmel Loughland^59^, Nicholas G. Martin^87,191^, Carol A. Mathews^192^, Fermin Mayoral^91^, Susan L. McElroy^193^, Andrew M. McIntosh^127,194^, Francis J. McMahon^195^, Ingrid Melle^6,196^, Patricia Michie^59^, Lili Milani^27^, Philip B. Mitchell^175^, Gunnar Morken^21,197^, Ole Mors^14,198^, Preben Bo Mortensen^12,14,38,39^, Bryan Mowry^176^, Bertram Müller-Myhsok^62,199,200^, Richard M. Myers^124^, Benjamin M. Neale^25,45,179^, Caroline M. Nievergelt^89,201^, Merete Nordentoft^14,202^, Markus M. Nöthen^3^, Michael C. O’Donovan^75^, Ketil J. Oedegaard^203,204^, Tomas Olsson^205^, Michael J. Owen^75^, Sara A. Paciga^206^, Chris Pantelis^207^, Carlos Pato^108^, Michele T. Pato^108^, George P. Patrinos^110,208,209^, Roy H. Perlis^210,211^, Danielle Posthuma^212,213^, Josep Antoni Ramos-Quiroga^53,54,55,56^, Andreas Reif^57^, Eva Z. Reininghaus^50^, Marta Ribasés^53,54,56,145^, Marcella Rietschel^78^, Stephan Ripke^25,28,45^, Guy A. Rouleau^126,214^, Takeo Saito^24^, Ulrich Schall^59^, Martin Schalling^118,119^, Peter R. Schofield^152,153^, Thomas G. Schulze^36,78,81,136,215^, Laura J. Scott^172^, Rodney J. Scott^59^, Alessandro Serretti^216^, Cynthia Shannon Weickert^152,175,217^, Jordan W. Smoller^25,133,218^, Hreinn Stefansson^32^, Kari Stefansson^32,219^, Eystein Stordal^220,221^, Fabian Streit^78^, Patrick F. Sullivan^20,222,223^, Gustavo Turecki^224^, Arne E. Vaaler^225^, Eduard Vieta^226^, John B. Vincent^102^, Irwin D. Waldman^227^, Thomas W. Weickert^152,175,217^, Thomas Werge^14,228,229,230^, Naomi R. Wray^11,231^, John-Anker Zwart^18,19,33^, Joanna M. Biernacka^8,64^, John I. Nurnberger^232^, Sven Cichon^3,4,95,96^, Howard J. Edenberg^77,233^, Eli A. Stahl^1,2,179,236^, Andrew McQuillin^40,236^, Arianna Di Florio^75,223,236^, Roel A. Ophoff^23,79,117,234,236^ and Ole A. Andreassen^6,7,236†^

**Affiliations**

1 Department of Genetics and Genomic Sciences, Icahn School of Medicine at Mount Sinai, New York, NY, USA. 2 Department of Psychiatry, Icahn School of Medicine at Mount Sinai, New York, NY, USA. 3 Institute of Human Genetics, University of Bonn, School of Medicine and University Hospital Bonn, Bonn, Germany. 4 Institute of Neuroscience and Medicine (INM-1), Research Centre Jülich, Jülich, Germany. 5 Centre for Human Genetics, University of Marburg, Marburg, Germany. 6 Division of Mental Health and Addiction, Oslo University Hospital, Oslo, Norway. 7 NORMENT, University of Oslo, Oslo, Norway. 8 Department of Health Sciences Research, Mayo Clinic, Rochester, MN, USA. 9 Social, Genetic and Developmental Psychiatry Centre, King’s College London, London, UK. 10 NIHR Maudsley BRC, King’s College London, London, UK. 11 Institute for Molecular Bioscience, The University of Queensland, Brisbane, Queensland, Australia. 12 iSEQ, Center for Integrative Sequencing, Aarhus University, Aarhus, Denmark. 13 Department of Biomedicine – Human Genetics, Aarhus University, Aarhus, Denmark. 14 iPSYCH, The Lundbeck Foundation Initiative for Integrative Psychiatric Research, Aarhus, Denmark. 15 Department of Psychiatry and Behavioral Sciences, SUNY Downstate Health Sciences University, Brooklyn, NY, USA. 16 VA NY Harbor Healthcare System, Brooklyn, NY, USA. 17 Research and Communication Unit for Musculoskeletal Health, Division of Clinical Neuroscience, Oslo University Hospital, Oslo, Norway. 18 Institute of Clinical Medicine, University of Oslo, Oslo, Norway. 19 K. G. Jebsen Center for Genetic Epidemiology, Department of Public Health and Nursing, Faculty of Medicine and Health Sciences, Norwegian University of Science and Technology, Trondheim, Norway. 20 Department of Medical Epidemiology and Biostatistics, Karolinska Institutet, Stockholm, Sweden. 21 Department of Mental Health, Faculty of Medicine and Health Sciences, Norwegian University of Science and Technology (NTNU), Trondheim, Norway. 22 Department of Østmarka, Division of Mental Health Care, St Olavs Hospital, Trondheim University Hospital, Trondheim, Norway. 23 Department of Psychiatry and Biobehavioral Science, Semel Institute, David Geffen School of Medicine, University of California, Los Angeles, Los Angeles, CA, USA. 24 Department of Psychiatry, School of Medicine, Fujita Health University, Toyoake, Japan. 25 Stanley Center for Psychiatric Research, Broad Institute, Cambridge, MA, USA. 26 Department of Genetics, Harvard Medical School, Boston, MA, USA. 27 Estonian Genome Center, Institute of Genomics, University of Tartu, Tartu, Estonia. 28 Department of Psychiatry and Psychotherapy, Charité -Universitätsmedizin, Berlin, Germany. 29 Department of Neuroscience, Icahn School of Medicine at Mount Sinai, New York, NY, USA. 30 Ronald M. Loeb Center for Alzheimer’s Disease, Icahn School of Medicine at Mount Sinai, New York, NY, USA. 31 Estelle and Daniel Maggin Department of Neurology, Icahn School of Medicine at Mount Sinai, New York, NY, USA. 32 deCODE Genetics/Amgen, Reykjavik, Iceland. 33 Department of Research, Innovation and Education, Division of Clinical Neuroscience, Oslo University Hospital, Oslo, Norway. 34 Samsung Advanced Institute for Health Sciences and Technology (SAIHST), Samsung Medical Center, Sungkyunkwan University, Seoul, South Korea. 35 Russian Academy of Medical Sciences, Mental Health Research Center, Moscow, Russian Federation. 36 Institute of Psychiatric Phenomics and Genomics (IPPG), University Hospital, LMU Munich, Munich, Germany. 37 Department of Psychiatry and Psychotherapy, University Hospital, LMU Munich, Munich, Germany. 38 National Centre for Register-Based Research, Aarhus University, Aarhus, Denmark. 39 Centre for Integrated Register-Based Research, Aarhus University, Aarhus, Denmark. 40 Division of Psychiatry, University College London, London, UK. 41 Department of Neuroscience, Istituto Di Ricerche Farmacologiche Mario Negri IRCCS, Milan, Italy. 42 Department of Psychiatry and Behavioral Neuroscience, University of Chicago, Chicago, IL, USA. 43 Northwestern University, Chicago, IL, USA. 44 Psychiatry, Berkshire Healthcare NHS Foundation Trust, Bracknell, UK. 45 Analytic and Translational Genetics Unit, Massachusetts General Hospital, Boston, MA, USA. 46 2nd Department of Psychiatry, Attikon General Hospital, National and Kapodistrian University of Athens, Athens, Greece. 47 Department of Psychiatry, Samsung Medical Center, School of Medicine, Sungkyunkwan University, Seoul, South Korea. 48 Center for Neonatal Screening, Department for Congenital Disorders, Statens Serum Institut, Copenhagen, Denmark. 49 Department of Psychiatry and Psychotherapy, University Hospital Carl Gustav Carus, Technische Universität Dresden, Dresden, Germany. 50 Department of Psychiatry and Psychotherapeutic Medicine, Medical University of Graz, Graz, Austria. 51 Department of Psychiatric Research, Diakonhjemmet Hospital, Oslo, Norway. 52 Psychiatry, Brain Center UMC Utrecht, Utrecht, the Netherlands. 53 Instituto de Salud Carlos III, Biomedical Network Research Centre on Mental Health (CIBERSAM), Madrid, Spain. 54 Department of Psychiatry, Hospital Universitari Vall d’Hebron, Barcelona, Spain. 55 Department of Psychiatry and Forensic Medicine, Universitat Autònoma de Barcelona, Barcelona, Spain. 56 Psychiatric Genetics Unit, Group of Psychiatry Mental Health and Addictions, Vall d’Hebron Research Institut (VHIR), Universitat Autònoma de Barcelona, Barcelona, Spain. 57 Department of Psychiatry, Psychosomatic Medicine and Psychotherapy, University Hospital Frankfurt, Frankfurt am Main, Germany. 58 Psychiatry, University of California San Francisco, San Francisco, CA, USA. 59 University of Newcastle, Newcastle, New South Wales, Australia. 60 Mood Disorders Program, Department of Psychiatry, McGill University Health Center, Montreal, Quebec, Canada. 61 Division of Psychiatry, University of Edinburgh, Edinburgh, UK. 62 Department of Translational Research in Psychiatry, Max Planck Institute of Psychiatry, Munich, Germany. 63 Department of Psychiatry, Universidad Autonoma de Nuevo Leon, Monterrey, Mexico. 64 Department of Psychiatry and Psychology, Mayo Clinic, Rochester, MN, USA. 65 Department of Laboratory Medicine and Pathology, Mayo Clinic, Rochester, MN, USA. 66 Centre for Psychiatry, Queen Mary University of London, London, UK. 67 UCL Genetics Institute, University College London, London, UK. 68 Department of Psychiatry, Laboratory of Psychiatric Genetics, Poznan University of Medical Sciences, Poznan, Poland. 69 Center for Multimodal Imaging and Genetics, Departments of Neurosciences, Radiology, and Psychiatry, University of California, San Diego, CA, USA. 70 Department of Child and Adolescent Psychiatry, Psychosomatics and Psychotherapy, University Hospital Essen, University of Duisburg-Essen, Duisburg, Germany. 71 Department of Medical Genetics, Oslo University Hospital, Oslo, Norway. 72 NORMENT, Department of Clinical Science, University of Bergen, Bergen, Norway. 73 Department of Neurology, Oslo University Hospital, Oslo, Norway. 74 NORMENT, KG Jebsen Centre for Psychosis Research, Oslo University Hospital, Oslo, Norway. 75 Medical Research Council Centre for Neuropsychiatric Genetics and Genomics, Division of Psychological Medicine and Clinical Neurosciences, Cardiff University, Cardiff, UK. 76 Academic Psychiatry, Newcastle University, Newcastle upon Tyne, UK. 77 Department of Medical and Molecular Genetics, Indiana University, Indianapolis, IN, USA. 78 Department of Genetic Epidemiology in Psychiatry, Central Institute of Mental Health, Medical Faculty Mannheim, Heidelberg University, Mannheim, Germany. 79 Center for Neurobehavioral Genetics, Semel Institute for Neuroscience and Human Behavior, Los Angeles, CA, USA. 80 Department of Clinical Neuroscience, Karolinska Institutet, Stockholm, Sweden. 81 Department of Psychiatry and Psychotherapy, University Medical Center Göttingen, Göttingen, Germany. 82 Department of Psychiatry, Dalhousie University, Halifax, Nova Scotia, Canada. 83 Department of Psychiatry, Yale School of Medicine, New Haven, CT, USA. 84 Veterans Affairs Connecticut Healthcare System, West Haven, CT, USA. 85 Departments of Genetics and Neuroscience, Yale University School of Medicine, New Haven, CT, USA. 86 Department of Psychological Sciences, University of Missouri, Columbia, MO, USA. 87 Genetics and Computational Biology, QIMR Berghofer Medical Research Institute, Brisbane, Queensland, Australia. 88 Psychological Medicine, University of Worcester, Worcester, UK. 89 Department of Psychiatry, University of California San Diego, La Jolla, CA, USA. 90 Bioinformatics Research Centre, Aarhus University, Aarhus, Denmark. 91 Mental Health Department, University Regional Hospital, Biomedicine Institute (IBIMA), Málaga, Spain. 92 Department of Psychiatry, Seoul National University College of Medicine, Seoul, South Korea. 93 Landspitali University Hospital, Reykjavik, Iceland. 94 Department of Psychology, Eberhard Karls Universität Tübingen, Tübingen, Germany. 95 Department of Biomedicine, University of Basel, Basel, Switzerland. 96 Institute of Medical Genetics and Pathology, University Hospital Basel, Basel, Switzerland. 97 Neuropsychiatrie Translationnelle, Inserm U955, Créteil, France. 98 Faculté de Santé, Université Paris Est, Créteil, France. 99 International Max Planck Research School for Translational Psychiatry (IMPRS-TP), Munich, Germany. 100 Laboratory of Complex Trait Genomics, Department of Computational Biology and Medical Sciences, Graduate School of Frontier Sciences, The University of Tokyo, Tokyo, Japan. 101 Laboratory for Statistical and Translational Genetics, RIKEN Center for Integrative Medical Sciences, Yokohama, Japan. 102 Campbell Family Mental Health Research Institute, Centre for Addiction and Mental Health, Toronto, Ontario, Canada. 103 Neurogenetics Section, Centre for Addiction and Mental Health, Toronto, Ontario, Canada. 104 Department of Psychiatry, University of Toronto, Toronto, Ontario, Canada. 105 Institute of Medical Sciences, University of Toronto, Toronto, Ontario, Canada. 106 Department of Psychiatry, Psychosomatics and Psychotherapy, Center of Mental Health, University Hospital Würzburg, Würzburg, Germany. 107 Cell Biology, SUNY Downstate Medical Center College of Medicine, Brooklyn, NY, USA. 108 Institute for Genomic Health, SUNY Downstate Medical Center College of Medicine, Brooklyn, NY, USA. 109 ISGlobal, Barcelona, Spain. 110 Laboratory of Pharmacogenomics and Individualized Therapy, Department of Pharmacy, School of Health Sciences, University of Patras, Patras, Greece. 111 Mental Illness Research, Education and Clinical Center, Crescenz VAMC, Philadelphia, PA, USA. 112 Center for Studies of Addiction, University of Pennsylvania Perelman School of Medicine, Philadelphia, PA, USA. 113 RIKEN Center for Integrative Medical Sciences, Yokohama, Japan. 114 Psychiatry, Altrecht, Utrecht, the Netherlands. 115 Psychiatry, GGZ inGeest, Amsterdam, the Netherlands. 116 Psychiatry, VU Medisch Centrum, Amsterdam, the Netherlands. 117 Department of Psychiatry, Erasmus MC, University Medical Center Rotterdam, Rotterdam, the Netherlands. 118 Department of Molecular Medicine and Surgery, Karolinska Institutet, Stockholm, Sweden. 119 Center for Molecular Medicine, Karolinska University Hospital, Stockholm, Sweden. 120 Psychiatry, North East London NHS Foundation Trust, Ilford, UK. 121 Clinic for Psychiatry and Psychotherapy, University Hospital Cologne, Cologne, Germany. 122 Department of Psychiatry, Korea University College of Medicine, Seoul, South Korea. 123 Psychiatric and Neurodevelopmental Genetics Unit, Center for Genomic Medicine, Massachusetts General Hospital and Harvard Medical School, Boston, MA, USA. 124 HudsonAlpha Institute for Biotechnology, Huntsville, AL, USA. 125 Department of Human Genetics, McGill University, Montréal, Quebec, Canada. 126 Montreal Neurological Institute and Hospital, McGill University, Montréal, Quebec, Canada. 127 Division of Psychiatry, Centre for Clinical Brain Sciences, The University of Edinburgh, Edinburgh, UK. 128 Department of Psychiatry and Psychotherapy, University of Bonn, Bonn, Germany. 129 Clinical Biochemistry Laboratory, Attikon General Hospital, Medical School, National and Kapodistrian University of Athens, Athens, Greece. 130 Department of Clinical Neuroscience, Centre for Psychiatry Research, Karolinska Institutet, Stockholm, Sweden. 131 Systems Genetics Working Group, Department of Genetics, Stellenbosch University, Stellenbosch, South Africa. 132 Genetic Cancer Susceptibility Group, International Agency for Research on Cancer, Lyon, France. 133 Department of Psychiatry, Massachusetts General Hospital, Boston, MA, USA. 134 Centre for Neuroimaging and Cognitive Genomics (NICOG), National University of Ireland Galway, Galway, Ireland. 135 Medical Faculty, School of Science and Technology, University Sarajevo, Sarajevo, Bosnia and Herzegovina. 136 Department of Psychiatry and Behavioral Sciences, Johns Hopkins University School of Medicine, Baltimore, MD, USA. 137 Oxford Health NHS Foundation Trust, Warneford Hospital, Oxford, UK. 138 Department of Psychiatry, University of Oxford, Warneford Hospital, Oxford, UK. 139 Department of Psychiatry and Behavioral Sciences, Emory University School of Medicine, Atlanta, GA, USA. 140 Outpatient Clinic for Bipolar Disorder, Altrecht, Utrecht, the Netherlands. 141 Department of Psychiatry, Washington University in Saint Louis, Saint Louis, MO, USA. 142 Department of Biochemistry and Molecular Biology II, Faculty of Pharmacy, University of Granada, Granada, Spain. 143 Institute of Neurosciences, Biomedical Research Center (CIBM), University of Granada, Granada, Spain. 144 Medicine, Psychiatry, Biomedical Informatics, Vanderbilt University Medical Center, Nashville, TN, USA. 145 Department of Genetics, Microbiology and Statistics, Faculty of Biology, Universitat de Barcelona, Barcelona, Spain. 146 Faculty of Medicine, Department of Psychiatry, School of Health Sciences, University of Iceland, Reykjavik, Iceland. 147 Institute of Health and Wellbeing, University of Glasgow, Glasgow, UK. 148 Psychiatry and the Behavioral Sciences, University of Southern California, Los Angeles, CA, USA. 149 Mood Disorders, PsyQ, Rotterdam, the Netherlands. 150 SAMRC Unit on Risk and Resilience in Mental Disorders, Department of Psychiatry and Neuroscience Institute, University of Cape Town, Cape Town, South Africa. 151 Department of Environmental Epidemiology, Nofer Institute of Occupational Medicine, Lodz, Poland. 152 Neuroscience Research Australia, Sydney, New South Wales, Australia. 153 School of Medical Sciences, University of New South Wales, Sydney, New South Wales, Australia. 154 Centro de Biología Molecular Severo Ochoa, Universidad Autónoma de Madrid and CSIC, Madrid, Spain. 155 Department of Psychiatry and Human Behavior, School of Medicine, University of California, Irvine, Irvine, CA, USA. 156 Psychiatry, Psychiatrisches Zentrum Nordbaden, Wiesloch, Germany. 157 Computational Sciences Center of Emphasis, Pfizer Global Research and Development, Cambridge, MA, USA. 158 Dalla Lana School of Public Health, University of Toronto, Toronto, Ontario, Canada. 159 Department of Psychological Medicine, Institute of Psychiatry, Psychology and Neuroscience, King’s College London, London, UK. 160 South London and Maudsley NHS Foundation Trust, Bethlem Royal Hospital, Beckenham, UK. 161 Department of Clinical Sciences, Psychiatry, Umeå University Medical Faculty, Umeå, Sweden. 162 NORMENT, KG Jebsen Centre for Psychosis Research, Division of Mental Health and Addiction, Institute of Clinical Medicine and Diakonhjemmet Hospital, University of Oslo, Oslo, Norway. 163 National Institute of Mental Health, Klecany, Czech Republic. 164 Institute of Environmental Medicine, Karolinska Institutet, Stockholm, Sweden. 165 Institute of Pulmonology, Russian State Medical University, Moscow, Russian Federation. 166 Department of Psychiatry, University of Münster, Münster, Germany. 167 Department of Psychiatry, Melbourne Medical School, The University of Melbourne, Melbourne, Victoria, Australia. 168 The Florey Institute of Neuroscience and Mental Health, The University of Melbourne, Parkville, Victoria, Australia. 169 Université de Paris, INSERM, Optimisation Thérapeutique en Neuropsychopharmacologie, UMRS 1144, Paris, France. 170 APHP Nord, DMU Neurosciences, Département de Psychiatrie et de Médecine Addictologique, GHU Saint Louis-Lariboisière-Fernand Widal, Paris, France. 171 Psychiatry, University of Pennsylvania, Philadelphia, PA, USA. 172 Center for Statistical Genetics and Department of Biostatistics, University of Michigan, Ann Arbor, MI, USA. 173 Department of Biomedicine and the iSEQ Center, Aarhus University, Aarhus, Denmark. 174 Center for Genomics and Personalized Medicine, CGPM, Aarhus, Denmark. 175 School of Psychiatry, University of New South Wales, Sydney, New South Wales, Australia. 176 University of Queensland, Brisbane, Queensland, Australia. 177 Neuropsychiatric Genetics Research Group, Department of Psychiatry and Trinity Translational Medicine Institute, Trinity College Dublin, Dublin, Ireland. 178 1st Department of Psychiatry, Eginition Hospital, National and Kapodistrian University of Athens, Athens, Greece. 179 Medical and Population Genetics, Broad Institute, Cambridge, MA, USA. 180 Division of Endocrinology, Children’s Hospital Boston, Boston, MA, USA. 181 Department of Human Genetics, University of Chicago, Chicago, IL, USA. 182 Biometric Psychiatric Genetics Research Unit, Alexandru Obregia Clinical Psychiatric Hospital, Bucharest, Romania. 183 HUNT Research Center, Department of Public Health and Nursing, Faculty of Medicine and Health Sciences, Norwegian University of Science and Technology, Trondheim, Norway. 184 University of Western Australia, Nedlands, Western Australia, Australia. 185 Institute of Neuroscience and Physiology, University of Gothenburg, Gothenburg, Sweden. 186 Department of Psychiatry and Addiction Medicine, Assistance Publique - Hôpitaux de Paris, Paris, France. 187 Department of Medical and Molecular Genetics, King’s College London, London, UK. 188 Neuroscience Therapeutic Area, Janssen Research and Development, LLC, Titusville, NJ, USA. 189 Cancer Epidemiology and Prevention, M. Sklodowska-Curie National Research Institute of Oncology, Warsaw, Poland. 190 SA MRC Unit on Risk and Resilience in Mental Disorders, Department of Psychiatry, Stellenbosch University, Stellenbosch, South Africa. 191 School of Psychology, The University of Queensland, Brisbane, Queensland, Australia. 192 Department of Psychiatry and Genetics Institute, University of Florida, Gainesville, FL, USA. 193 Research Institute, Lindner Center of HOPE, Mason, OH, USA. 194 Centre for Cognitive Ageing and Cognitive Epidemiology, University of Edinburgh, Edinburgh, UK. 195 Human Genetics Branch, Intramural Research Program, National Institute of Mental Health, Bethesda, MD, USA. 196 Division of Mental Health and Addiction, University of Oslo, Institute of Clinical Medicine, Oslo, Norway. 197 Psychiatry, St Olavs University Hospital, Trondheim, Norway. 198 Psychosis Research Unit, Aarhus University Hospital - Psychiatry, Risskov, Denmark. 199 Munich Cluster for Systems Neurology (SyNergy), Munich, Germany. 200 University of Liverpool, Liverpool, UK. 201 Research/Psychiatry, Veterans Affairs San Diego Healthcare System, San Diego, CA, USA. 202 Mental Health Services in the Capital Region of Denmark, Mental Health Center Copenhagen, University of Copenhagen, Copenhagen, Denmark. 203 Division of Psychiatry, Haukeland Universitetssjukehus, Bergen, Norway. 204 Faculty of Medicine and Dentistry, University of Bergen, Bergen, Norway. 205 Department of Clinical Neuroscience and Center for Molecular Medicine, Karolinska Institutet at Karolinska University Hospital, Solna, Sweden. 206 Human Genetics and Computational Biomedicine, Pfizer Global Research and Development, Groton, CT, USA. 207 University of Melbourne, Melbourne, Victoria, Australia. 208 Department of Pathology, College of Medicine and Health Sciences, United Arab Emirates University, Al-Ain, United Arab Emirates. 209 Zayed Center of Health Sciences, United Arab Emirates University, Al-Ain, United Arab Emirates. 210 Psychiatry, Harvard Medical School, Boston, MA, USA. 211 Division of Clinical Research, Massachusetts General Hospital, Boston, MA, USA. 212 Department of Complex Trait Genetics, Center for Neurogenomics and Cognitive Research, Amsterdam Neuroscience, Vrije Universiteit Amsterdam, Amsterdam, the Netherlands. 213 Department of Clinical Genetics, Amsterdam Neuroscience, Vrije Universiteit Medical Center, Amsterdam, the Netherlands. 214 Department of Neurology and Neurosurgery, Faculty of Medicine, McGill University, Montreal, Quebec, Canada. 215 Department of Psychiatry and Behavioral Sciences, SUNY Upstate Medical University, Syracuse, NY, USA. 216 Department of Biomedical and NeuroMotor Sciences, University of Bologna, Bologna, Italy. 217 Department of Neuroscience, SUNY Upstate Medical University, Syracuse, NY, USA. 218 Psychiatric and Neurodevelopmental Genetics Unit (PNGU), Massachusetts General Hospital, Boston, MA, USA. 219 Faculty of Medicine, University of Iceland, Reykjavik, Iceland. 220 Department of Psychiatry, Hospital Namsos, Namsos, Norway. 221 Department of Neuroscience, Norges Teknisk Naturvitenskapelige Universitet Fakultet for Naturvitenskap og Teknologi, Trondheim, Norway. 222 Department of Genetics, University of North Carolina at Chapel Hill, Chapel Hill, NC, USA. 223 Department of Psychiatry, University of North Carolina at Chapel Hill, Chapel Hill, NC, USA. 224 Department of Psychiatry, McGill University, Montreal, Quebec, Canada. 225 Department of Psychiatry, Sankt Olavs Hospital Universitetssykehuset i Trondheim, Trondheim, Norway. 226 Clinical Institute of Neuroscience, Hospital Clinic, University of Barcelona, IDIBAPS, CIBERSAM, Barcelona, Spain. 227 Department of Psychology, Emory University, Atlanta, GA, USA. 228 Institute of Biological Psychiatry, Mental Health Services, Copenhagen University Hospital, Copenhagen, Denmark. 229 Department of Clinical Medicine, University of Copenhagen, Copenhagen, Denmark. 230 Center for GeoGenetics, GLOBE Institute, University of Copenhagen, Copenhagen, Denmark. 231 Queensland Brain Institute, The University of Queensland, Brisbane, Queensland, Australia. 232 Psychiatry, Indiana University School of Medicine, Indianapolis, IN, USA. 233 Biochemistry and Molecular Biology, Indiana University School of Medicine, Indianapolis, IN, USA. 234 Department of Human Genetics, David Geffen School of Medicine, University of California Los Angeles, Los Angeles, CA, USA. 235 These authors contributed equally: Niamh Mullins, Andreas J. Forstner. 236 These authors jointly supervised this work: Eli A. Stahl, Andrew McQuillin, Arianna Di Florio, Roel A. Ophoff, Ole A. Andreassen. * A list of members and their affiliations appears in the Supplementary Information of Mullins et al., Nat Genet, 2021; 53(6):817-829. Kevin S. O’Connell, Brandon Coombes, Jonathan R. I. Coleman and Zhen Qiao contributed equally to this work and should be regarded as joint second authors.

†

#### Major Depressive Disorder Working Group of the Psychiatric Genomics Consortium <u>(2023)</u>

Mark J Adams 1 *

Fabian Streit 2 *

Swapnil Awasthi 3 *

Brett N Adey 4

Karmel W Choi 5, 6

V Kartik Chundru 7

Jonathan RI Coleman 4, 8

Jerome C Foo 2

Olga Giannakopoulou 9

Alisha S M Hall 2, 10

Jens Hjerling-Leffler 11

David M Howard 4

Christopher Hübel 4, 12, 13

Alex S F Kwong 1, 14

Bochao Danae Lin 15

Xiangrui Meng 9

Guiyan Ni 16

Oliver Pain 17

Gita A Pathak 18, 19

Eva C Schulte 20, 21, 22, 23

Jackson G Thorp 24

Alicia Walker 16

Shuyang Yao 25

Jian Zeng 16

Johan Zvrskovec 4, 8

Dag Aarsland 26

Ky’Era V Actkins 27

Mazda Adli 3, 28

Esben Agerbo 12, 29, 30

Mareike Aichholzer 31

Tracy M Air 32

Allison Aiello 33

Thomas D Als 30, 34, 35

Evelyn Andersson 36

Till F M Andlauer 37, 38

Volker Arolt 39

Helga Ask 40, 41

Sunita Badola 42

Clive Ballard 43

Karina Banasik 44

Nicholas J Bass 9

Aartjan T F Beekman 45

Sintia Belangero 46

Elisabeth B Binder 38, 47

Ottar Bjerkeset 48, 49

Gyda Bjornsdottir 50

Julia Boberg 36

Sigrid Børte 51, 52, 53

Emma Bränn 54

Alice Braun 55

Thorsten Brodersen 56

Søren Brunak 44

Mie T Bruun 57

Pichit Buspavanich 58, 59

Jonas Bybjerg-Grauholm 60,61

Enda M Byrne 62

Archie Campbell 63, 64

Megan L. Campbell 65

Enrique Castelao 66

Jorge Cervilla 67, 68

Boris Chaumette 69

Chia-Yen Chen 70

Zhengming Chen 71, 72

Sven Cichon 73, 74, 75, 76

Lucía Colodro-Conde 24

Anne Corbett 43

Elizabeth C Corfield 40, 77

Baptiste Couvy-Duchesne 78

Nick Craddock 79, 80

Udo Dannlowski 39

Gail Davies 81

EJC de Geus 82

Ian J Deary 81

Franziska Degenhardt 76, 83

Abbas Dehghan 84, 85

J Raymond DePaulo 86

Michael Deuschle 87

Maria Didriksen 88

Khoa Manh Dinh 89

Nese Direk 90

Srdjan Djurovic 91, 92

Anna R Docherty 93, 94, 95

Katharina Domschke 96

Joseph Dowsett 88

Ole Kristian Drange 49, 97, 98, 99

Erin C Dunn 6, 100

Gudmundur Einarsson 50

Thalia C Eley 4

Samar S M Elsheikh 101

Jan Engelmann 102

Michael E Benros 60, 103, 104

Christian Erikstrup 89

Valentina Escott-Price 80

Chiara Fabbri 4, 105

Yu Fang 106

Sarah Finer 107

Josef Frank 2

Robert C Free 108

He Gao 109

Michael Gill 110

Maria Gilles 87

Fernando S Goes 86

Scott Douglas Gordon 24

Jakob Grove 30, 34, 35, 111

Daniel F Gudbjartsson 50, 112

Blanca Gutierrez 67, 68

Tim Hahn 39

Lynsey S Hall 80

Thomas F Hansen 44, 60,113

Magnus Haraldsson 114

Catherina A Hartman 115

Alexandra Havdahl 40

Caroline Hayward 116

Stefanie Heilmann-Heimbach 76

Stefan Herms 74, 76

Ian B Hickie 117

Henrik Hjalgrim 118

Per Hoffmann 74, 76

Georg Homuth 119

Carsten Horn 120

Jouke-Jan Hottenga 82

David M Hougaard 60, 61

Iiris Hovatta 121

Qin Qin Huang 7

Floris Huider 82

Karen A Hunt 122

Marcus Ising 123

Erkki Isometsä 124

Rick Jansen 45

Yunxuan Jiang 125

Ian Jones 80

Lisa A Jones 126

Lina Jonsson 127

Robert Karlsson 25

Siegfried Kasper 128

Kenneth S Kendler 129

Ronald C Kessler 130

Stefan Kloiber 101, 123, 131, 132

James A Knowles 133

Nastassja Koen 65

Julia Kraft 55

Henry R Kranzler 134, 135

Kristi Krebs 136

Theodora Kunovac Kallak 137

Zoltán Kutalik 138, 139, 140

Elisa Lahtela 141

Margit Hørup Larsen 88

Eric J Lenze 142

Daniel F Levey 143, 144

Melissa Lewins 1

Glyn Lewis 9

Liming Li 145, 146

Kuang Lin 71

Penelope A Lind 24

Donald J MacIntyre 1, 147, 148

Dean F MacKinnon 86

Hermine HM Maes 149, 150

Wolfgang Maier 151

Victoria S Marshe 101, 152

Hamdi Mbarek 82

Peter McGuffin 4

Sarah E Medland 24

Susanne Meinert 39, 153

Susan Mikkelsen 89

Christina Mikkelsen 88, 154

Yuri Milaneschi 45

Iona Y Millwood 71, 72

Brittany L Mitchell 24

Esther Molina 67, 155

Francis M Mondimore 86

Preben Bo Mortensen 12, 29, 30

Benoit H Mulsant 101, 131

Joonas Naamanka 121

Jake M Najman 156

Matthias Nauck 157, 158

Igor Nenadić 159

Kasper R Nielsen 160

Ilja M Nolte 161

Merete Nordentoft 60, 103, 104

Markus M Nöthen 76

Mette Nyegaard 30, 162, 163, 164

Michael C O’Donovan 80

Asmundur Oddsson 50

Catherine M Olsen 165, 166

Hogni Oskarsson 167

Sisse Rye Ostrowski 88, 168

Vanessa K Ota 46

Michael J Owen 80

Richard Packer 169

Teemu Palviainen 141

Pedro M Pan 170

Carlos N Pato 171

Michele T Pato 171

Nancy L Pedersen 25

Ole Birger Pedersen 172

Roseann E Peterson 129, 173

Wouter J Peyrot 45

James B Potash 86

Martin Preisig 66

Jorge A Quiroz 174

Charles F Reynolds III 175

John P Rice 142

Giovanni A Salum 176

Robert A Schoevers 177, 178

Andrew Schork 30, 179, 180

Thomas G Schulze 2, 21, 86, 181, 182

Tabea S Send 87

Jianxin Shi 183

Engilbert Sigurdsson 114

Kritika Singh 27

Grant C B Sinnamon 184

Lea Sirignano 2

Olav B Smeland 185, 186

Daniel J Smith 187

Erik Sørensen 88

Sundararajan Srinivasan 188

Hreinn Stefansson 50

Kari Stefansson 50, 189

Dan J. Stein 190

Frederike Stein 191

André Tadic 102, 192

Henning Teismann 193

Alexander Teumer 194

Anita Thapar 80, 195

Pippa A Thomson 64

Lise Wegner Thørner 88

Apostolia Topaloudi 196

Ioanna Tzoulaki 84, 85, 197

Monica Uddin 198

André G Uitterlinden 199

Henrik Ullum 88, 200, 201

Daniel Umbricht 202

Robert J Ursano 203

Sandra Van der Auwera 204

David A van Heel 122

Albert M van Hemert 205

Abirami Veluchamy 188

Alexander Viktorin 25

Henry Völzke 194

Agaz Wani 198

G Bragi Walters 50

Robin G Walters 71, 72

Sylvia Wassertheil-Smoller 206

Myrna M Weissman 207, 208

Jürgen Wellmann 193

David C Whiteman 165

Derek Wildman 198

Gonneke Willemsen 82

Alexander T Williams 169

Bendik S Winsvold 51, 52, 209

Stephanie H Witt 2

Ying Xiong 25

Lea Zillich 2

John-Anker Zwart 51, 52, 53

23andMe Research Team 125

Estonian Biobank Research Team 136

HUNT All-In Psychiatry 210

China Kadoorie Biobank Collaborative Group 211

Genes & Health Research Team 212

Ole A Andreassen 185, 186, 213

Bernhard T Baune 214, 215, 216

Klaus Berger 193

Dorret I Boomsma 82

Anders D Børglum 30, 34, 35

Gerome Breen 4, 8

Na Cai 217, 218, 219

Hilary Coon 94

William E Copeland 220

Byron Creese 43

Lea K Davis 27

Eske M Derks 24

Enrico Domenici 221

Paul Elliott 84, 85, 197, 222

Andreas J Forstner 73, 76

Micha Gawlik 223

Joel Gelernter 19, 143, 224

Hans J Grabe 204

Steven P Hamilton 225

Kristian Hveem 226, 227, 228

Catherine John 169, 229

Jaakko Kaprio 141

Tilo Kircher 159

Marie-Odile Krebs 230

Karoline Kuchenbaecker 9, 71

Mikael Landén 25, 127

Kelli Lehto 136

Douglas F Levinson 231

Qingqin S Li 232

Klaus Lieb 102

Yi Lu 25

Susanne Lucae 123

Jurjen J Luykx 15, 233

Patrik K Magnusson 25

Nicholas G Martin 24

Hilary C Martin 7

Andrew McQuillin 9

Christel M Middeldorp 62, 234

Lili Milani 136

Ole Mors 30, 235

Daniel J Müller 101, 131, 132, 236

Bertram Müller-Myhsok 38, 237, 238

Albertine J Oldehinkel 115

Sara A Paciga 239

Colin NA Palmer 188

Peristera Paschou 196

Brenda WJH Penninx 45

Roy H Perlis 5, 6, 240

Giorgio Pistis 66

Renato Polimanti 18, 19

Patrick F Sullivan 25, 253

Martin Tesli 40

Thorgeir E Thorgeirsson 50

Henning Tiemeier 254, 255

Nicholas J Timpson 14

Rudolf Uher 256

Jens R Wendland 42

David J Porteous 64

Danielle Posthuma 241, 242

Ted Reichborn-Kjennerud 40

Andreas Reif 31

Frances Rice 80, 243

Roland Ricken 3

Marcella Rietschel 2

Margarita Rivera 67, 244

Christian Rück 245

Catherine Schaefer 246

Srijan Sen 106, 247

Alessandro Serretti 105

Alkistis Skalkidou 137

Jordan W Smoller 5, 248, 249

Frederike Stein 191

Murray B Stein 250, 251, 252

Thomas Werge 60, 179, 201, 257, 258

Naomi R Wray 16, 259 **

Stephan Ripke 3, 248 **

Cathryn M Lewis 4, 260 **

Andrew M McIntosh 1, 261 **

* Joint Lead Authors

** Joint Last Authors

**Affiliations**

1, Division of Psychiatry, University of Edinburgh, Edinburgh, UK

2, Department of Genetic Epidemiology in Psychiatry, Central Institute of Mental Health, Medical Faculty Mannheim, Heidelberg University, Mannheim, BW, DE

3, Department of Psychiatry and Psychotherapy, Charité – Universitätsmedizin Berlin, Berlin, BE, DE

4, Social, Genetic and Developmental Psychiatry Centre, King’s College London, London, UK

5, Department of Psychiatry, Massachusetts General Hospital, Boston, MA, US

6, Department of Psychiatry, Harvard Medical School, Boston, MA, US

7, Human Genetics, Wellcome Sanger Institute, Hinxton, UK

8, NIHR Maudsley Biomedical Research Centre, King’s College London, London, UK

9, Division of Psychiatry, University College London, London, UK

10, Department of Clinical Medicine, Aarhus University, Aarhus, DK

11, Department of Medical Biochemistry and Biophysics, Karolinska Institutet, Stockholm, SE

12, National Centre for Register-based Research, Aarhus University, Aarhus, DK

13, Department of Pediatric Neurology, Charité – Universitätsmedizin Berlin, Berlin, BE, DE

14, MRC Integrative Epidemiology Unit, University of Bristol, Bristol, UK

15, Department of Psychiatry and Neuropsychology, School for Mental Health and Neuroscience, Maastricht University Medical Centre, Maastricht, NL

16, Institute for Molecular Bioscience, University of Queensland, Brisbane, QLD, AU

17, Maurice Wohl Clinical Neuroscience Institute, Department of Basic and Clinical Neuroscience, King’s College London, London, UK

18, Veterans Affairs Connecticut Healthcare System, West Haven, CT, US

19, Department of Psychiatry, Yale University School of Medicine, New Haven, CT, US

20, Department of Psychiatry, University of Munich, Munich, BY, DE

21, Institute of Psychiatric Phenomics and Genomics, University of Munich, Munich, BY, DE

22, Department of Psychiatry and Psychotherapy, University Hospital Bonn, Medical Faculty, University of Bonn, Bonn, DE

23, Institute of Human Genetics, University Hospital Bonn, Medical Faculty, University of Bonn, Bonn, DE

24, Mental Health and Neuroscience, QIMR Berghofer Medical Research Institute, Brisbane, QLD, AU

25, Department of Medical Epidemiology and Biostatistics, Karolinska Institutet, Stockholm, SE

26, Old Age Psychiatry, King’s College London, London, UK

27, Department of Medicine, Division of Genetic Medicine, Vanderbilt University Medical Center, Nashville, TN, US

28, Department of Psychiatry and Psychotherapy, Fliedner Klinik Berlin, Berlin, BE, DE

29, Centre for Integrated Register-based Research, Aarhus University, Aarhus, DK

30, iPSYCH, The Lundbeck Foundation Initiative for Integrative Psychiatric Research, Aarhus, DK

31, Department of Psychiatry, Psychosomatic Medicine and Psychotherapy, Goethe University Frankfurt - University Hospital, Frankfurt am Main, DE

32, Discipline of Psychiatry, University of Adelaide, Adelaide, SA, AU

33, Department of Epidemiology, Columbia University Mailman School of Public Health, New York, NY, US

34, Department of Biomedicine and Centre for Integrative Sequencing, iSEQ, Aarhus University, Aarhus, DK

35, Center for Genomics and Personalized Medicine, Aarhus University, Aarhus, DK

36, Department of Clinical Neuroscience, Karolinska Institutet,, SE

37, Department of Neurology, Klinikum rechts der Isar, Technical University of Munich, Munich, BY, DE

38, Department of Translational Research in Psychiatry, Max Planck Institute of Psychiatry, Munich, BY, DE

39, Institute for Translational Psychiatry, University of Münster, Münster, NRW, DE

40, Department of Mental Disorders, Norwegian Institute of Public Health, Oslo, NO

41, PROMENTA Research Center, Department of Psychology, University of Oslo, Oslo, NO

42, Research and Development, Takeda Pharmaceutical Company Limited, Cambridge, MA, US

43, Faculty of Health and Life Sciences, University of Exeter, Exeter, UK

44, Novo Nordisk Center for Protein Research, Department of Health Sciences, University of Copenhagen, Copenhagen, DK

45, Department of Psychiatry, Amsterdam Public Health and Amsterdam Neuroscience, Amsterdam UMC, Vrije Universiteit Amsterdam, Amsterdam, NL

46, Morphology and Genetics, Universidade Federal de Sao Paulo, Sao Paulo, SP, BR

47, Department of Psychiatry and Behavioral Sciences, Emory University School of Medicine, Atlanta, GA, US

48, Faculty of Nursing and Health Sciences, NORD University, Levanger, NO

49, Department of Mental Health, Faculty of Medicine and Health Sciences, Norwegian University of Science and Technology (NTNU), Trondheim, TRD, NO

50, deCODE Genetics / Amgen, Reykjavik, IS

51, K. G. Jebsen Center for Genetic Epidemiology, Department of Public Health and Nursing, Faculty of Medicine and Health Sciences, Norwegian University of Science and Technology (NTNU), Trondheim, TRD, NO

52, Department of Research and Innovation, Division of Clinical Neuroscience, Oslo University Hospital, Oslo, NO

53, Institute of Clinical Medicine, Faculty of Medicine, University of Oslo, Oslo, NO

54, Institute of Environmental Medicine, Unit of Integrative Epidemiology, Karolinska Institutet, Stockholm, SE

55, Department of Psychiatry and Psychotherapy, Charité – Universitätsmedizin Berlin, Berlin, DE

56, Department of Clinical Immunology, Roskilde University/Næstved Hospital, Roskilde, DK

57, Department of Clinical Immunology, Odense University Hospital, Odense, DK

58, Department of Psychiatry, Psychotherapy and Psychosomatics, Brandenburg Medical School Theodor Fontane, Neuruppin, BB, DE

59, Department of Psychiatry and Psychotherapy, Gender Research in Medicine, Institute of Sexology and Sexual Medicine, Charité – Universitätsmedizin Berlin, Berlin, BE, DE

60, iPSYCH, The Lundbeck Foundation Initiative for Integrative Psychiatric Research, Copenhagen, DK

61, Center for Neonatal Screening, Department for Congenital Disorders, Statens Serum Institut, Copenhagen, DK

62, Child Health Research Centre, University of Queensland, Brisbane, QLD, AU

63, Centre for Medical Informatics, Usher Institute, University of Edinburgh, Edinburgh, UK

64, Centre for Genomic & Experimental Medicine, Institute for Genetics and Cancer, University of Edinburgh, Edinburgh, UK

65, Department of Psychiatry and Mental Health, University of Cape Town, Cape Town, SA

66, Department of Psychiatry, Lausanne University Hospital and University of Lausanne, Prilly, VD, CH

67, Instituto de Investigación Biosanitaria ibs.GRANADA, Granada, ES

68, Department of Psychiatry, Faculty of Medicine and Institute of Neurosciences, Biomedical Research Centre (CIBM), University of Granada, Granada, ES

69, Université de Paris Cité, INSERM U1266, Institute of Psychiatry and Neuroscience of Paris, GHU Paris Psychiatry and Neuroscience, Paris, FR

70, Translational Biology, Biogen, Cambridge, MA, US

71, Nuffield Department of Population Health, University of Oxford, Oxford, UK

72, MRC Population Health Research Unit, University of Oxford, Oxford, UK

73, Institute of Neuroscience and Medicine (INM-1), Research Center Juelich, Juelich, DE

74, Human Genomics Research Group, Department of Biomedicine, University of Basel, Basel, CH

75, Institute of Medical Genetics and Pathology, University Hospital Basel, University of Basel, Basel, CH

76, Institute of Human Genetics, University of Bonn, School of Medicine & University Hospital Bonn, Bonn, DE

77, Nic Waals Institute, Lovisenberg Diakonale Hospital, Oslo, NO

78, Centre for Advanced Imaging, University of Queensland, Saint Lucia, QLD, AU

79, Psychological Medicine, Cardiff University, Cardiff, WLS, UK

80, Centre for Neuropsychiatric Genetics and Genomics, Cardiff University, Cardiff, WLS, UK

81, The Lothian Birth Cohorts, University of Edinburgh, Edinburgh, UK

82, Department of Biological Psychology & Amsterdam Public Health Research Institute, Vrije Universiteit Amsterdam, Amsterdam, NL

83, Department of Child and Adolescent Psychiatry, Psychosomatics and Psychotherapy, University Hospital Essen, Unversity of Duisburg-Essen, Duisburg, DE

84, MRC Centre for Environment and Health, School of Public Health, Imperial College London, London, UK

85, Imperial College Dementia Research Institute, Imperial College London, London, UK

86, Department of Psychiatry and Behavioral Sciences, Johns Hopkins University School of Medicine, Baltimore, MD, US

87, Department of Psychiatry and Psychotherapy, Research Group Stress Related Disorders, Central Institute of Mental Health, Medical Faculty Mannheim, Heidelberg University, Mannheim, BW, DE

88, Department of Clinical Immunology, Copenhagen University Hospital, Rigshospitalet, Copenhagen, CPH, DK

89, Department of Clinical Immunology, Aarhus University Hospital, Aarhus, DK

90, Department of Psychiatry, Istanbul University, Istanbul, TR

91, Department of Medical Genetics, Oslo University Hospital, Oslo, OSL, NO

92, NORMENT, Department of Clinical Science, University of Bergen, Bergen, NO

93, Virginia Institute for Psychiatric & Behavioral Genetics, Virginia Commonwealth University, Richmond, VA, US

94, Psychiatry Department / Huntsman Mental Health Institute, University of Utah School of Medicine, Salt Lake City, UT, US

95, Center for Genomic Research, University of Utah School of Medicine, Salt Lake City, UT, US

96, Department of Psychiatry and Psychotherapy, Medical Center, University of Freiburg, Faculty of Medicine, University of Freiburg, Freiburg, DE

97, Division of Mental Health Care, St. Olavs Hospital, Trondheim University Hospital, Trondheim, TRD, NO

98, Department of Psychiatry, Sørlandet Hospital, Kristiansand, AG, NO

99, University of Oslo, NORMENT Centre, Institute of Clinical Medicine, Oslo, OSL, NO

100, Center for Genomic Medicine, Massachusetts General Hospital, Boston, MA, US

101, Centre for Addiction and Mental Health, Toronto, ON, CA

102, Department of Psychiatry and Psychotherapy, University Medical Center of the Johannes Gutenberg University Mainz, Mainz, DE

103, Mental Health Center Copenhagen, Mental Health Services Capital Region of Denmark, Copenhagen, DK

104, Faculty of Health Science, Department of Clinical Medicine, University of Copenhagen, Copenhagen, DK

105, Department of Biomedical and Neuromotor Sciences, University of Bologna, Bologna, IT

106, Michigan Neuroscience Institute, University of Michigan, Ann Arbor, MI, US

107, Wolfson Institute of Population Health, Queen Mary University of London, London, UK

108, School of Computing and Mathematical Sciences, University of Leicester, Leicester, UK

109, Department of Epidemiology and Biostatistics, Imperial College London, London, UK

110, Discipline of Psychiatry, School of Medicine, Trinity College Dublin, Dublin, IE

111, Bioinformatics Research Centre, Aarhus University, Aarhus, DK

112, School of Engineering, University of Iceland, Reykjavik, IS

113, Danish Headache Centre, Department of Neurology, Rigshospitalet, Glostrup, DK

114, Faculty of Medicine, Department of Psychiatry, University of Iceland, Reykjavik, IS

115, Department of Psychiatry, University of Groningen, University Medical Center Groningen, Groningen, NL

116, MRC Human Genetics Unit, Institute for Genetics and Cancer, University of Edinburgh, Edinburgh, UK

117, Brain and Mind Centre, University of Sydney, Sydney, NSW, AU

118, Department of Epidemiology Research, Statens Serum Institut, Copenhagen, DK

119, Interfaculty Institute for Genetics and Functional Genomics, Department of Functional Genomics, University Medicine Greifswald, Greifswald, MV, DE

120, Roche Pharmaceutical Research and Early Development, Pharmaceutical Sciences, Roche Innovation Center Basel, F. Hoffmann-La Roche Ltd, Basel, CH

121, SleepWell Research Program and Department of Psychology and Logopedics, University of Helsinki, Helsinki, FI

122, Blizard Institute, Barts and the London School of Medicine and Dentistry, Queen Mary University of London, London, UK

123, Max Planck Institute of Psychiatry, Munich, BY, DE

124, Department of Psychiatry, University of Helsinki, Helsinki, FI

125, 23andMe Research Team, 23andMe, Inc., Sunnyvale, CA, US

126, Department of Psychological Medicine, University of Worcester, Worcester, UK

127, Institution of Neuroscience and Physiology, University of Gothenburg, Gothenburg, SE

128, Department of Psychiatry and Psychotherapy, Medical University of Vienna, Vienna, AT

129, Department of Psychiatry, Virginia Commonwealth University, Richmond, VA, US

130, Health Care Policy, Harvard Medical School, Boston, MA, US

131, Department of Psychiatry, University of Toronto, Toronto, ON, CA

132, Department of Pharmacology & Toxicology, University of Toronto, Toronto, ON, CA

133, Department of Genetics, Rutgers University, Piscataway, NJ, US

134, Department of Psychiatry, Perelman School of Medicine, University of Pennsylvania, Philadelphia, PA, US

135, Mental Illness Research, Education and Clinical Center, Crescenz VA Medical Center, Philadelphia, PA, US

136, Estonian Genome Centre, Institute of Genomics, University of Tartu, Tartu, EE

137, Department of Women’s and Children’s Health, Uppsala University, Uppsala, SE

138, Department of Epidemiology and Health Systems, Center for Primary Care and Public Health, Lausanne, VD, CH

139, Swiss Institute of Bioinformatics, Lausanne, VD, CH

140, Department of Computational Biology, University of Lausanne, Lausanne, VD, CH

141, Institute for Molecular Medicine Finland - FIMM, University of Helsinki, Helsinki, FI

142, Department of Psychiatry, Washington University School of Medicine in St. Louis, St. Louis, MO, US

143, Psychiatry, Veterans Affairs Connecticut Healthcare System, West Haven, CT, US

144, Department of Psychiatry, Yale University, New Haven, CT, US

145, Department of Epidemiology and Biostatistics, School of Public Health, Peking University, Beijing, CN

146, Peking University Center for Public Health and Epidemic Preparedness & Response, Peking University, Beijing, CN

147, Mental Health, NHS

24, Glasgow, UK

148, Royal Edinburgh Hospital, NHS Lothian, Edinburgh, UK

149, Department of Human and Molecular Genetics, Virginia Commonwealth University, Richmond, VA, USA

150, Virginia Institute for Psychiatric and Behavioral Genetics, Virginia Commonwealth University, Richmond, VA, USA

151, Department of Psychiatry and Psychotherapy, University of Bonn, Bonn, DE

152, Center for Translational and Computational Neuroimmunology, Columbia University Medical Center, New York, NY, US

153, Institute for Translational Neuroscience, University of Münster, Münster, NRW, DE

154, Novo Nordisk Foundation Center for Basic Metabolic Research, Faculty of Health Science, Copenhagen University, Copenhagen, DK

155, Department of Nursing, Faculty of Health Sciences and Institute of Neurosciences, Biomedical Research Centre (CIBM), University of Granada, Granada, ES

156, School of Public Health, University of Queensland, Brisbane, QLD, AU

157, DZHK (German Centre for Cardiovascular Research), Partner Site Greifswald, Greifswald, MV, DE

158, Institute of Clinical Chemistry and Laboratory Medicine, University Medicine Greifswald, Greifswald, MV, DE

159, Department of Psychiatry, University of Marburg, Marburg, DE

160, Department of Clinical Immunology, Aalborg University Hospital, Aalborg, DK

161, Department of Epidemiology, University of Groningen, University Medical Center Groningen, Groningen, NL

162, Department of Health, Science and Technology, Aalborg University, Aalborg, DK

163, Centre for Integrative Sequencing, iSEQ, Aarhus University, Aarhus, DK

164, Department of Biomedicine-Human Genetics, Aarhus University, Aarhus, DK

165, Population Health, QIMR Berghofer Medical Research Institute, Brisbane, QLD, AU

166, The Fraser Institute, Faculty of Medicine, University of Queensland, Brisbane, QLD, AU

167, Humus, Reykjavik, IS

168, Department of Clinical Medicine, University of Copenhagen, Copenhagen, CPH, DK

169, Department of Population Health Sciences, University of Leicester, Leicester, UK

170, Department of Psychiatry, Universidade Federal de Sao Paulo, Sao Paulo, SP, BR

171, Department of Psychiatry, Rutgers University, Piscataway, NJ, US

172, Department of Clinical Immunology, Zealand University Hospital, Køge, DK

173, Department of Psychiatry and Behavioral Sciences, SUNY Downstate Health Sciences University, Brooklyn, NY, US

174, NMD Pharma, Lexington, MA, US

175, Psychiatry, University of Pittsburgh Medical Centre, Pittsburgh, PA, US

176, Psychiatry, Universidade Federal do Rio Grande do Sul, Porto Alegre, BR

177, Department of Psychiatry, University Medical Center Groningen, Groningen, NL

178, Research School of Behavioural and Cognitive Neurosciences (BCN), University of Groningen, Groningen, NL

179, Institute of Biological Psychiatry, Mental Health Center Sct. Hans, Mental Health Services Capital Region of Denmark, Copenhagen, DK

180, Neurogenomics Division, The Translational Genomics Research Institute (TGEN), Phoenix, AZ, US

181, Human Genetics Branch, NIMH Division of Intramural Research Programs, Bethesda, MD, US

182, Department of Psychiatry and Psychotherapy, University Medical Center Göttingen, Goettingen, NI, DE

183, Division of Cancer Epidemiology and Genetics, National Cancer Institute, Bethesda, MD, US

184, School of Medicine and Dentistry, James Cook University, Townsville, QLD, AU

185, Division of Mental Health and Addiction, Oslo University Hospital, Oslo, OSL, NO

186, NORMENT, Institute of Clinical Medicine, University of Oslo, Oslo, OSL, NO

187, Institute of Health and Wellbeing, University of Glasgow, Glasgow, UK

188, Division of Population Health and Genomics, Ninewells Hospital and School of Medicine, University of Dundee, Dundee, UK

189, Faculty of Medicine, University of Iceland, Reykjavik, IS

190, SAMRC Unit on Risk & Resilience in Mental Disorders, Department of Psychiatry and Mental Health, University of Cape Town, Cape Town, SA

191, Department of Psychiatry and Psychotherapy, University of Marburg, Marburg, HE, DE

192, Department of Psychiatry, Psychotherapy and Psychosomatics, Dr. Fontheim Mentale Gesundheit, Liebenburg, DE

193, Institute of Epidemiology and Social Medicine, University of Münster, Münster, NRW, DE

194, Institute for Community Medicine, University Medicine Greifswald, Greifswald, MV, DE

195, Wolfson Centre for Young People’s Mental Health, Division of Psychological Medicine and Clinical Neurosciences, Cardiff University, Cardiff, WLS, UK

196, Department of Biological Sciences, Purdue University, West Lafayette, IN, US

197, Imperial College BHF Centre for Research Excellence, Imperial College London, London, UK

198, Genomics Program, University of South Florida College of Public Health, Tampa, FL, US

199, Department of Internal Medicine, Erasmus University Medical Center Rotterdam, Rotterdam, NL

200, Management Section, Statens Serum Institut, Copenhagen, DK

201, Department of Clinical Medicine, University of Copenhagen, Copenhagen, DK

202, Xperimed LLC, Basel, CH

203, Psychiatry, USUHS, Bethesda, US

204, Department of Psychiatry and Psychotherapy, University Medicine Greifswald, Greifswald, MV, DE

205, Department of Psychiatry, Leiden University Medical Center, Leiden, NL

206, Department of Epidemiology and Population Health, Albert Einstein College of Medicine, Bronx, NY, US

207, Department of Psychiatry, Columbia University College of Physicians and Surgeons, New York, NY, US

208, Division of Epidemiology, New York State Psychiatric Institute, New York, NY, US

209, Department of Neurology, Oslo University Hospital, Oslo, NO

210, HUNT All-In Psychiatry

211, China Kadoorie Biobank Collaborative Group 212, Genes & Health Research Team

213, KG Jebsen Centre for Neurodevelopmental Research, University of Oslo, Oslo, OSL, NO

214, Department of Psychiatry, University of Melbourne, Melbourne, VIC, AU

215, Florey Institute of Neuroscience and Mental Health, University of Melbourne, Melbourne, VIC, AU

216, Department of Psychiatry, University of Münster, Münster, NRW, DE

217, Computational Health Centre, Helmholtz Zentrum München, Neuherberg, DE

218, School of Medicine, Technical University of Munich, Munich, BY, DE

219, Helmholtz Pioneer Campus, Helmholtz Zentrum München, Neuherberg, DE

220, Department of Psychiatry, University of Vermont, Burlington, VT, US

221, Department of Cellular, Computational and Integrative Biology, Università degli Studi di Trento, Trento, IT

222, Imperial College Biomedical Research Centre, Imperial College London, London, UK

223, Department of Psychiatry, Psychosomatics and Psychotherapy, Julius-Maximilians-Universität Würzburg, Würzburg, DE

224, Department of Genetics, Department of Neuroscience, Yale University School of Medicine, New Haven, CT, US

225, Psychiatry, Kaiser Permanente Northern California, San Francisco, CA, US

226, K. G. Jebsen Center for Genetic Epidemiology, Department of Public Health and Nursing, Faculty of Medicine and Health Sciences, Norwegian University of Science and Technology (NTNU), Trondheim, NO

227, HUNT Research Center, Department of Public Health and Nursing, Faculty of Medicine and Health Sciences, Norwegian University of Science and Technology (NTNU), Trondheim, NO

228, Department of Research, Innovation and Education, St. Olavs Hospital, Trondheim University Hospital, Trondheim, NO

229, NIHR Leicester Biomedical Research Centre, Glenfield Hospital, Leicester, UK

230, Pathophysiology of Psychiatric Diseases, INSERM, Univ Paris Cité, GHU Paris, Paris, FR

231, Department of Psychiatry & Behavioral Sciences, Stanford University, Stanford, CA, US

232, Neuroscience Therapeutic Area, Janssen Research and Development, LLC, Titusville, NJ, US

233, Second Opinion Outpatient Clinic, GGNet Mental Health, Warnsveld, NL

234, Child and Youth Mental Health Service, Children’s Health Queensland Hospital and Health Service, Brisbane, QLD, AU

235, Psychosis Research Unit, Aarhus University Hospital-Psychiatry, Aarhus, DK

236, Department of Psychiatry, Psychosomatics and Psychotherapy, University Hospital of Würzburg, Würzburg, DE

237, Munich Cluster for Systems Neurology (SyNergy), Munich, BY, DE

238, University of Liverpool, Liverpool, UK

239, Human Genetics and Computational Biomedicine, Pfizer Global Research and Development, Groton, CT, US

240, Centre for Quantitative Health, Massachusetts General Hospital, Boston, MA, US

241, Child and Adolescent Psychiatry, Amsterdam UMC, Vrije Universiteit Amsterdam, Amsterdam, NL

242, Complex Trait Genetics, Vrije Universiteit Amsterdam, Amsterdam, NL

243, Wolfson Centre for Young People’s Mental Health, Division of Psychological Medicine and Clinical Neurosciences, Cardiff University, Cardiff, UK

244, Department of Biochemistry and Molecular Biology II, Faculty of Pharmacy and Institute of Neurosciences, Biomedical Research Centre (CIBM), University of Granada, Granada, ES

245, Department of Clinical Neuroscience, Karolinska Institutet, Stockholm, SE

246, Division of Research, Kaiser Permanente Northern California, Oakland, CA, US

247, Department of Psychiatry, University of Michigan, Ann Arbor, MI, US

248, Stanley Center for Psychiatric Research, Broad Institute of MIT and Harvard, Cambridge, MA, US

249, Psychiatric and Neurodevelopmental Genetics Unit, Massachusetts General Hospital, Boston, MA, US

250, Psychiatry, UCSD School of Medicine, La Jolla, CA, US

251, Public Health, UCSD School of Public Health, La Jolla, CA, US

252, Psychiatry, Veterans Affairs San Diego Healthcare System, San Diego, CA, US

253, Departments of Genetics and Psychiatry, University of North Carolina at Chapel Hill, Chapel Hill, NC, US

254, Child and Adolescent Psychiatry, Erasmus University Medical Center Rotterdam, Rotterdam, NL

255, Social and Behavioral Science, Harvard T.H. Chan School of Public Health, Boston, MA, US

256, Psychiatry, Dalhousie University, Halifax, NS, CA

257, Institute of Biological Psychiatry, Mental Health Center Sct. Hans, Copenhagen University Hospital, Mental Health Services, Copenhagen, DK

258, GLOBE Institute, Lundbeck Foundation Centre for Geogenetics, University of Copenhagen, Copenhagen, DK

259, Queensland Brain Institute, University of Queensland, Brisbane, QLD, AU

260, Department of Medical & Molecular Genetics, King’s College London, London, UK

261, Institute for Genomics and Cancer, University of Edinburgh, Edinburgh, UK

#### Integrative Psychiatric Research (iPSYCH) Study Consortium

| Group | PI Full Name |
| --- | --- |
| iPSYCH | Anders D. Børglum<sup>1,2,3</sup> |
| iPSYCH | David M. Hougaard<sup>4</sup> |
| iPSYCH | Merete Nordentoft<sup>5</sup> |
| iPSYCH | Ole Mors Mors<sup>6</sup> |
| iPSYCH | Preben Bo Mortensen<sup>7,8,9</sup> |
| iPSYCH | Thomas Werge<sup>10,11</sup> |
| Aarhus University | Ditte Demontis<sup>12</sup> |
| Aarhus University | Jakob Grove<sup>2,12,13,14</sup> |
| Aarhus University | Anna Starnawska<sup>13</sup> |
| Aarhus University | Thomas Damm Als<sup>13</sup> |
| Institute of Biological Psychiatry (IBP) | Alfonso Buil<sup>10,11</sup> |
| Institute of Biological Psychiatry (IBP) | Anders Rosengren<sup>10</sup> |
| Institute of Biological Psychiatry (IBP) | Dorte Helenius<sup>10</sup> |
| Institute of Biological Psychiatry (IBP) | Andrés Ingason<sup>10,11</sup> |
| Institute of Biological Psychiatry (IBP) | Jesper Gådin<sup>10</sup> |
| Institute of Biological Psychiatry (IBP) | Richard Zetterberg<sup>10</sup> |
| Institute of Biological Psychiatry (IBP) | Vivek Appadurai<sup>10</sup> |
| National Centre for Register-based Research (NCRR) | Bjarni J. Vilhjálmsson<sup>14,15</sup> |
| National Centre for Register-based Research (NCRR) | Carsten Bøcker Pedersen<sup>15</sup> |
| National Centre for Register-based Research (NCRR) | Jakob Christensen<sup>15</sup> |
| National Centre for Register-based Research (NCRR) | Liselotte V. Petersen<sup>15</sup> |
| National Centre for Register-based Research (NCRR) | Marianne Giørtz Pedersen<sup>15</sup> |
| Statens Serum Institut (SSI) | Jonas Bybjerg-Grauholm<sup>4</sup> |
| Statens Serum Institut (SSI) | Marie Bækvad-Hansen<sup>4</sup> |

1, Department of Biomedicine, Aarhus University, Aarhus, Denmark

2, The Lundbeck Foundation Initiative for Integrative Psychiatric Research, iPSYCH, Denmark

3, Center for Genomics and Personalized Medicine, Aarhus, Denmark

4, Department for Congenital Disorders, Statens Serum Institute, Copenhagen, Denmark

5, Mental Health Centre Copenhagen, Capital Region of Denmark, Copenhagen University Hospital, Copenhagen, Denmark

6, Psychosis Research Unit, Aarhus University Hospital-Psychiatry, Denmark

7, NCRR - National Centre for Register-Based Research, Business and Social Sciences, Aarhus University, Aarhus V, Denmark

8, Centre for Integrated Register-based Research, CIRRAU, Aarhus University, Aarhus, Denmark

9, Centre for Integrative Sequencing, Department of Biomedicine and iSEQ, Aarhus University, Aarhus, Denmark;

10, Institute of Biological Psychiatry, Mental Health Center - Sct Hans, Copenhagen University Hospital, Copenhagen, Denmark

11, Section for Geogenetics, GLOBE Institute, Faculty of Health and Medical Sciences, Copenhagen University

12, Center for Genomics and Personalized Medicine, Aarhus, Denmark

13, Department of Biomedicine, Aarhus University, Aarhus, Denmark

14, BiRC - Bioinformatics Research Centre, Aarhus University, Aarhus, Denmark

15, National Centre for Register-based Research, BSS, Aarhus University

