## Supplementary Figures and Tables for "Identifying genetic differences between bipolar disorder and major depression through multiple GWAS": SF5.forest_plots.BDvsMDD.pdf

| | | | | | | | |
|---|---|---|---|---|---|---|---|
| **rs10933** | | **C/T** | | **3:52719816** | | | |
| −++−+++++−+++ | | | | het_P: | 0.795 | het_I: | 0.0 |
| | **ngt** | **info** | **p_value** | **f_ca(n)** | **f_co(n)** | **ln(OR)** | **STDerr** |
| dan_bvd_grp1−usa1−2_eur | 0 | 0.98 | 0.213 | 0.553(2636) | 0.570(1019) | −0.0668 | 0.0536 |
| dan_bvd_grp1−usa1_eur | 0 | 0.89 | 0.0934 | 0.545(1604) | 0.567(1242) | −0.0967 | 0.0576 |
| dan_bvd_grp10−uk1r_eur | 0 | 0.96 | 0.00244 | 0.520(2516) | 0.553(2045) | −0.132 | 0.0435 |
| dan_bvd_grp11−ger2_eur | 0 | 0.93 | 0.25 | 0.528(234) | 0.558(442) | −0.138 | 0.12 |
| dan_bvd_grp12−swes_eur | 0 | 1.00 | 0.104 | 0.552(1273) | 0.573(1544) | −0.0886 | 0.0544 |
| dan_bvd_grp13−usa6_eur | 0 | 0.91 | 0.58 | 0.572(141) | 0.539(101) | 0.11 | 0.2 |
| dan_bvd_grp2−scot_eur | 0 | 0.85 | 0.536 | 0.545(280) | 0.559(368) | −0.0762 | 0.123 |
| dan_bvd_grp3−aus1_eur | 0 | 0.94 | 0.0512 | 0.506(328) | 0.543(1362) | −0.177 | 0.0907 |
| dan_bvd_grp4_ger1_eur | 0 | 0.93 | 0.0833 | 0.558(1338) | 0.578(2307) | −0.0888 | 0.0513 |
| dan_bvd_grp5−neth_eur | 0 | 0.94 | 0.921 | 0.573(737) | 0.570(247) | 0.0108 | 0.109 |
| dan_bvd_grp6−usa2_eur | 0 | 0.98 | 0.00067 | 0.538(3426) | 0.576(1496) | −0.153 | 0.045 |
| dan_bvd_grp7−usa3_eur | 0 | 0.96 | 0.813 | 0.550(375) | 0.556(281) | −0.0275 | 0.116 |
| dan_bvd_grp8−usa4_eur | 0 | 0.99 | 0.759 | 0.561(644) | 0.552(466) | 0.0269 | 0.0878 |
| **dan_** | **0** | **0.95** | **7.78e−08** | **0.544(15532)** | **0.565(12920)** | **−0.0988** | **0.0184** |

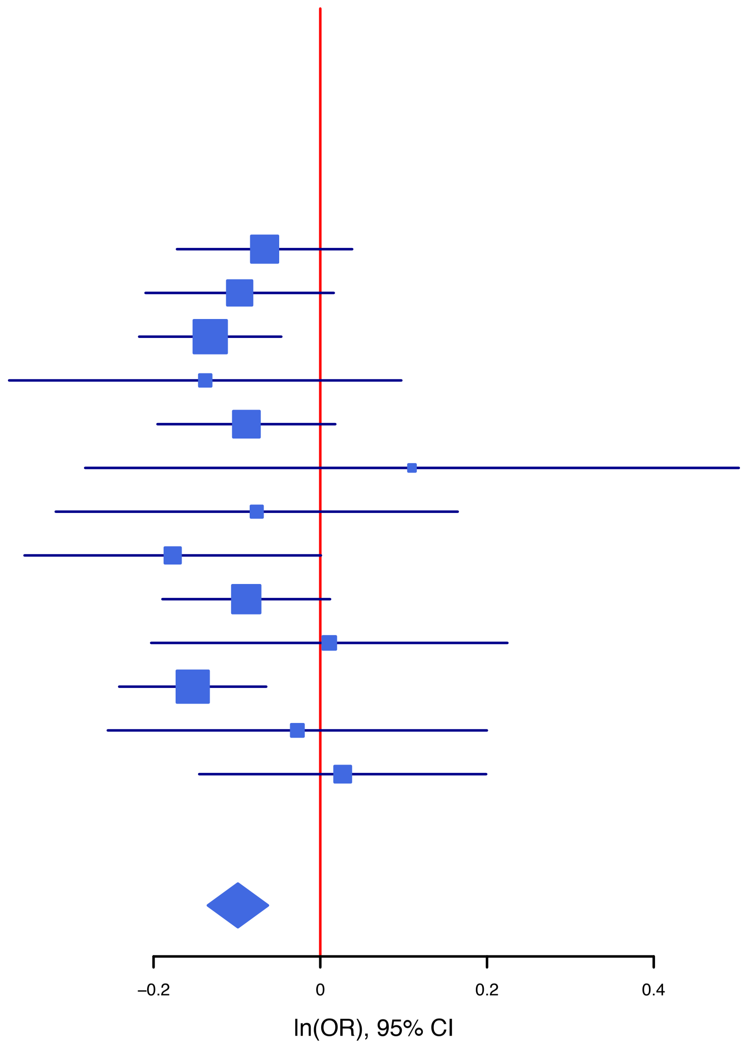

**rs11887238** **C/A** **2:58143267**

+----+-------- het_P: 0.341 het_I: 10.5

| | ngt | info | p_value | f_ca(n) | f_co(n) | ln(OR) | STDerr |
|---|---|---|---|---|---|---|---|
| dan_bvd_grp1−usa1−2_eur | 0 | 0.98 | 0.0556 | 0.964(2636) | 0.956(1019) | 0.256 | 0.134 |
| dan_bvd_grp1−usa1_eur | 0 | 1.03 | 0.0897 | 0.960(1604) | 0.949(1242) | 0.216 | 0.127 |
| dan_bvd_grp10−uk1r_eur | 0 | 0.97 | 0.0283 | 0.959(2516) | 0.949(2045) | 0.225 | 0.103 |
| dan_bvd_grp11−ger2_eur | 0 | 1.11 | 0.958 | 0.965(234) | 0.964(442) | 0.0155 | 0.294 |
| dan_bvd_grp12−swes_eur | 0 | 1.03 | 0.484 | 0.952(1273) | 0.947(1544) | 0.0854 | 0.122 |
| dan_bvd_grp13−usa6_eur | 0 | 0.97 | 0.294 | 0.989(141) | 0.975(101) | 0.799 | 0.762 |
| dan_bvd_grp2−scot_eur | 0 | 1.01 | 0.9 | 0.951(280) | 0.948(368) | 0.0328 | 0.26 |
| dan_bvd_grp3−aus1_eur | 0 | 0.97 | 0.603 | 0.957(328) | 0.953(1362) | 0.113 | 0.217 |
| dan_bvd_grp4_ger1_eur | 0 | 1.02 | 0.00408 | 0.975(1338) | 0.963(2307) | 0.421 | 0.147 |
| dan_bvd_grp5−neth_eur | 0 | 0.97 | 0.091 | 0.961(737) | 0.943(247) | 0.409 | 0.242 |
| dan_bvd_grp6−usa2_eur | 0 | 0.98 | 7.63e−05 | 0.964(3426) | 0.948(1496) | 0.421 | 0.106 |
| dan_bvd_grp7−usa3_eur | 0 | 1.00 | 0.811 | 0.960(375) | 0.963(281) | −0.0704 | 0.295 |
| dan_bvd_grp8−usa4_eur | 0 | 0.99 | 0.428 | 0.953(644) | 0.959(466) | −0.17 | 0.214 |
| **dan_** | **0** | **1.00** | **1.77e−07** | **0.962(15532)** | **0.953(12920)** | **0.23** | **0.0441** |

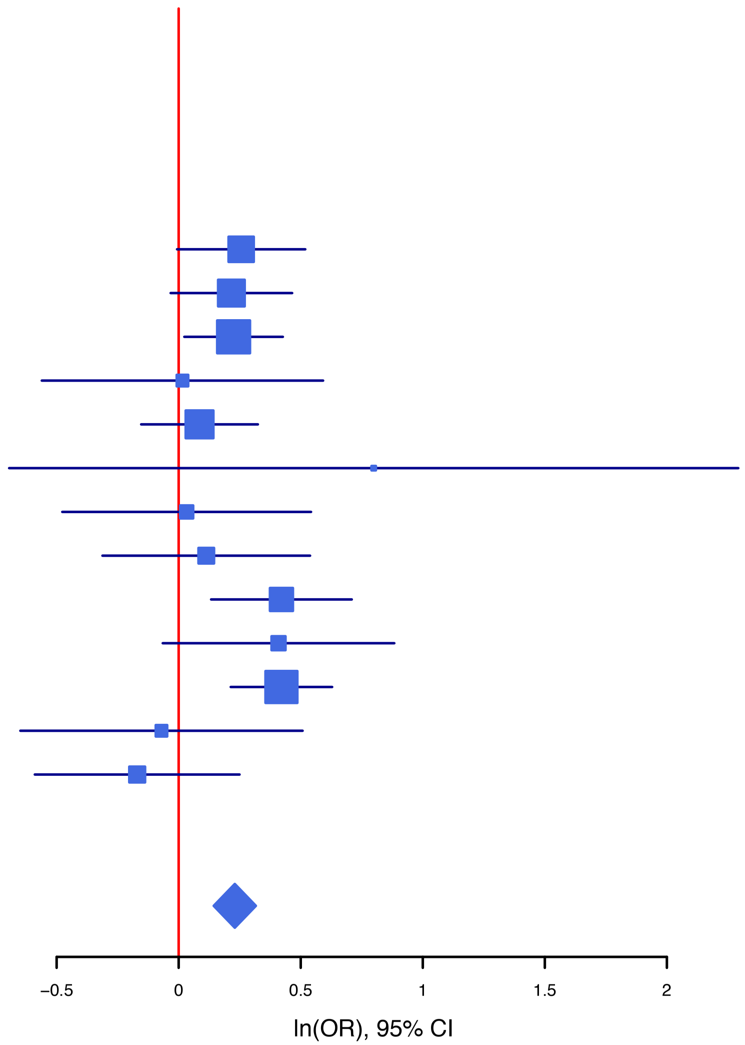

| rs13314421 | | A/G | | 3:36962388 | | | |
|---|---|---|---|---|---|---|---|
|  | ------+------- |  |  | het_P: | 0.574 | het_I: | 0.0 |
| | **ngt** | **info** | **p_value** | **f_ca(n)** | **f_co(n)** | **ln(OR)** | **STDerr** |
| dan_bvd_grp1–usa1–2_eur | 0 | 1.04 | 0.131 | 0.513(2636) | 0.530(1019) | −0.0779 | 0.0515 |
| dan_bvd_grp1–usa1_eur | 0 | 0.95 | 0.00724 | 0.509(1604) | 0.540(1242) | −0.149 | 0.0555 |
| dan_bvd_grp10–uk1r_eur | 0 | 0.96 | 0.165 | 0.518(2516) | 0.529(2045) | −0.0601 | 0.0433 |
| dan_bvd_grp11–ger2_eur | 0 | 0.98 | 0.0744 | 0.478(234) | 0.528(442) | −0.209 | 0.117 |
| dan_bvd_grp12–swes_eur | 0 | 0.99 | 0.24 | 0.529(1273) | 0.546(1544) | −0.064 | 0.0545 |
| dan_bvd_grp13–usa6_eur | 0 | 1.08 | 0.655 | 0.512(141) | 0.528(101) | −0.0803 | 0.18 |
| dan_bvd_grp2–scot_eur | 0 | 0.97 | 0.897 | 0.533(280) | 0.536(368) | −0.0148 | 0.115 |
| dan_bvd_grp3–aus1_eur | 0 | 0.98 | 0.0326 | 0.492(328) | 0.542(1362) | −0.19 | 0.0888 |
| dan_bvd_grp4_ger1_eur | 0 | 0.96 | 0.00339 | 0.497(1338) | 0.531(2307) | −0.147 | 0.0501 |
| dan_bvd_grp5–neth_eur | 0 | 0.97 | 0.181 | 0.492(737) | 0.529(247) | −0.143 | 0.107 |
| dan_bvd_grp6–usa2_eur | 0 | 0.98 | 0.0265 | 0.516(3426) | 0.539(1496) | −0.0988 | 0.0445 |
| dan_bvd_grp7–usa3_eur | 0 | 0.95 | 0.182 | 0.548(375) | 0.515(281) | 0.157 | 0.117 |
| dan_bvd_grp8–usa4_eur | 0 | 1.00 | 0.237 | 0.518(644) | 0.545(466) | −0.103 | 0.0871 |
| **dan_** | **0** | **0.98** | **8.14e−08** | **0.514(15532)** | **0.535(12920)** | **−0.0968** | **0.018** |

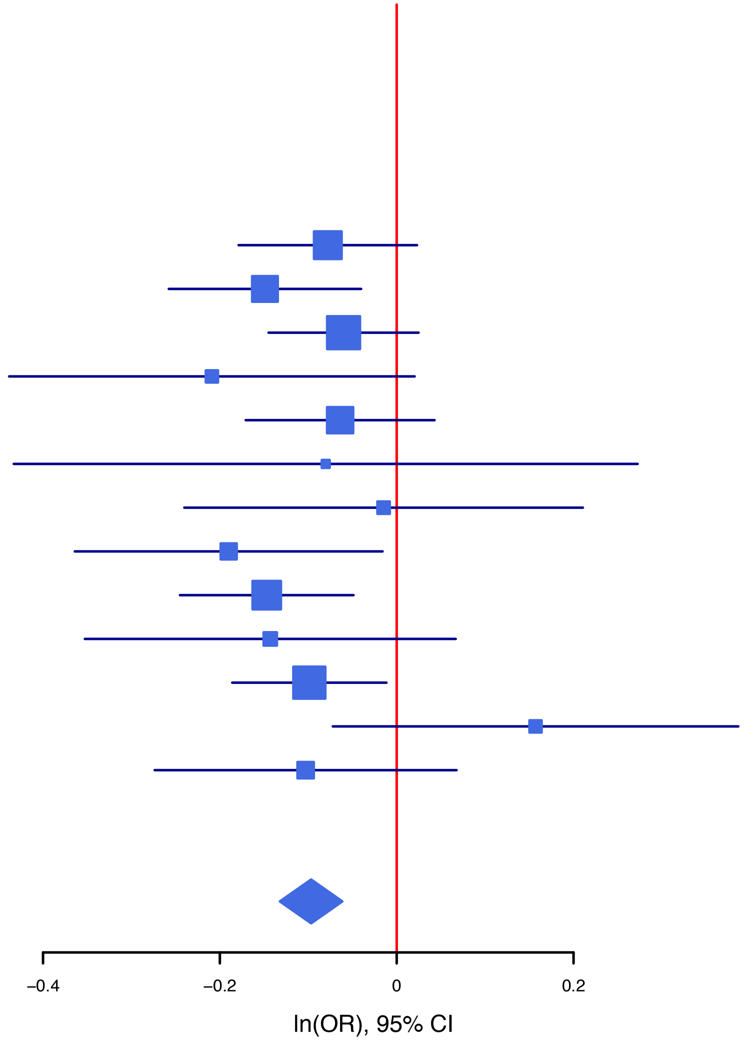

**rs1406857** **G/A** **20:37362432**

-----+-------- het_P: 0.772 het_I: 0.0

| | ngt | info | p_value | f_ca(n) | f_co(n) | ln(OR) | STDerr |
|---|---|---|---|---|---|---|---|
| dan_bvd_grp1–usa1–2_eur | 0 | 0.98 | 0.0704 | 0.681(2636) | 0.660(1019) | 0.101 | 0.056 |
| dan_bvd_grp1–usa1_eur | 0 | 0.94 | 0.538 | 0.687(1604) | 0.680(1242) | 0.0368 | 0.0598 |
| dan_bvd_grp10–uk1r_eur | 0 | 0.95 | 0.00094 | 0.699(2516) | 0.667(2045) | 0.154 | 0.0467 |
| dan_bvd_grp11–ger2_eur | 1 | 1.05 | 0.183 | 0.712(234) | 0.676(442) | 0.164 | 0.123 |
| dan_bvd_grp12–swes_eur | 1 | 1.00 | 0.463 | 0.726(1273) | 0.720(1544) | 0.0443 | 0.0604 |
| dan_bvd_grp13–usa6_eur | 0 | 1.01 | 0.446 | 0.663(141) | 0.630(101) | 0.152 | 0.199 |
| dan_bvd_grp2–scot_eur | 0 | 0.98 | 0.448 | 0.683(280) | 0.669(368) | 0.093 | 0.123 |
| dan_bvd_grp3–aus1_eur | 0 | 0.95 | 0.905 | 0.691(328) | 0.689(1362) | 0.0116 | 0.0969 |
| dan_bvd_grp4_ger1_eur | 0 | 0.93 | 0.332 | 0.692(1338) | 0.681(2307) | 0.0531 | 0.0547 |
| dan_bvd_grp5–neth_eur | 0 | 0.94 | 0.206 | 0.671(737) | 0.642(247) | 0.143 | 0.113 |
| dan_bvd_grp6–usa2_eur | 0 | 0.94 | 0.000854 | 0.688(3426) | 0.654(1496) | 0.161 | 0.0482 |
| dan_bvd_grp7–usa3_eur | 0 | 0.91 | 0.752 | 0.677(375) | 0.686(281) | –0.0406 | 0.129 |
| dan_bvd_grp8–usa4_eur | 0 | 0.99 | 0.263 | 0.683(644) | 0.663(466) | 0.104 | 0.0931 |
| **dan_** | **2** | **0.96** | **5.87e–07** | **0.691(15532)** | **0.677(12920)** | **0.0977** | **0.0196** |

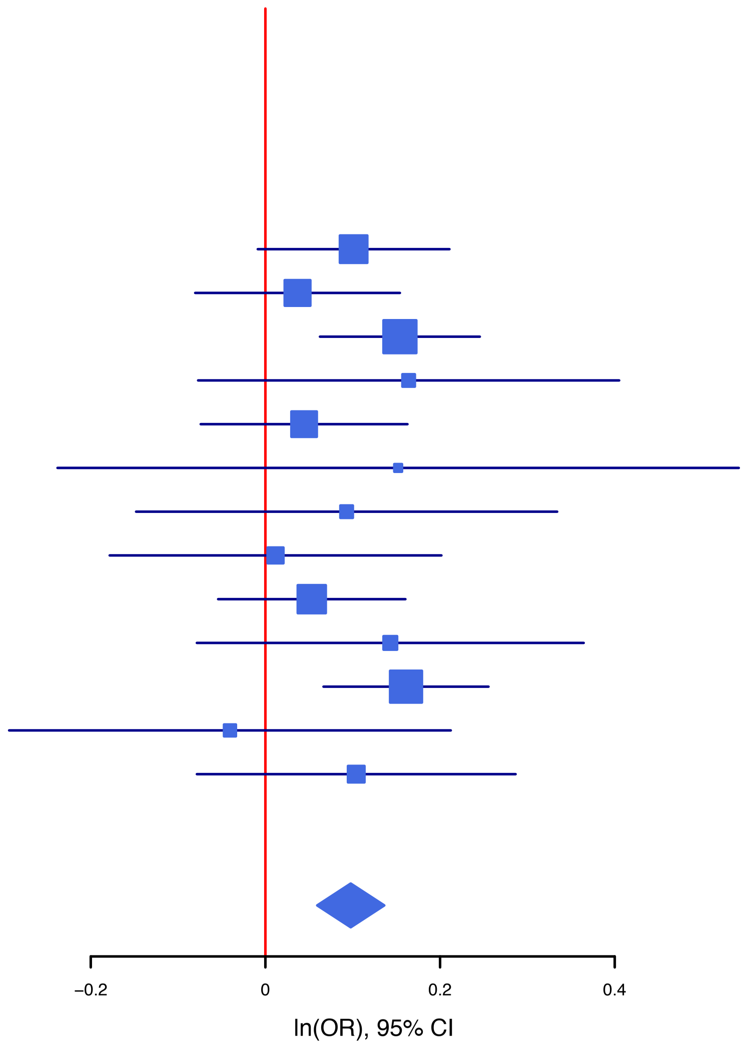

**rs324017** **A/C** **12:57487814**

--?-------+--? het_P: 0.529 het_I: 0.0

| | ngt | info | p_value | f_ca(n) | f_co(n) | ln(OR) | STDerr |
|---|---|---|---|---|---|---|---|
| dan_bvd_grp1–usa1–2_eur | 0 | 1.00 | 0.00638 | 0.278(2636) | 0.310(1019) | –0.156 | 0.0572 |
| dan_bvd_grp1–usa1_eur | 0 | 0.73 | 0.751 | 0.288(1604) | 0.286(1242) | 0.0222 | 0.0698 |
| dan_bvd_grp10–uk1r_eur | 0 | 0.97 | 0.936 | 0.290(2516) | 0.292(2045) | –0.00381 | 0.0472 |
| dan_bvd_grp11–ger2_eur | 0 | 0.95 | 0.0748 | 0.258(234) | 0.300(442) | –0.238 | 0.134 |
| dan_bvd_grp12–swes_eur | 0 | 0.99 | 0.0713 | 0.288(1273) | 0.311(1544) | –0.107 | 0.0594 |
| dan_bvd_grp13–usa6_eur | 0 | 0.97 | 0.431 | 0.246(141) | 0.274(101) | –0.171 | 0.217 |
| dan_bvd_grp2–scot_eur | 0 | 0.76 | 0.066 | 0.264(280) | 0.305(368) | –0.267 | 0.145 |
| dan_bvd_grp3–aus1_eur | 0 | 0.99 | 0.103 | 0.270(328) | 0.302(1362) | –0.16 | 0.0984 |
| dan_bvd_grp4_ger1_eur | 0 | 0.99 | 0.0299 | 0.277(1338) | 0.300(2307) | –0.119 | 0.0546 |
| dan_bvd_grp5–neth_eur | 0 | 1.01 | 0.826 | 0.297(737) | 0.295(247) | 0.0251 | 0.114 |
| dan_bvd_grp6–usa2_eur | 0 | 0.96 | 0.004 | 0.276(3426) | 0.303(1496) | –0.142 | 0.0493 |
| dan_bvd_grp7–usa3_eur | 0 | 0.99 | 0.402 | 0.274(375) | 0.286(281) | –0.107 | 0.127 |
| dan_bvd_grp8–usa4_eur | 0 | 0.97 | 0.1 | 0.274(644) | 0.306(466) | –0.159 | 0.0968 |
| **dan_** | **0** | **0.98** | **7.17e–07** | **0.280(15532)** | **0.301(12920)** | **–0.105** | **0.0213** |

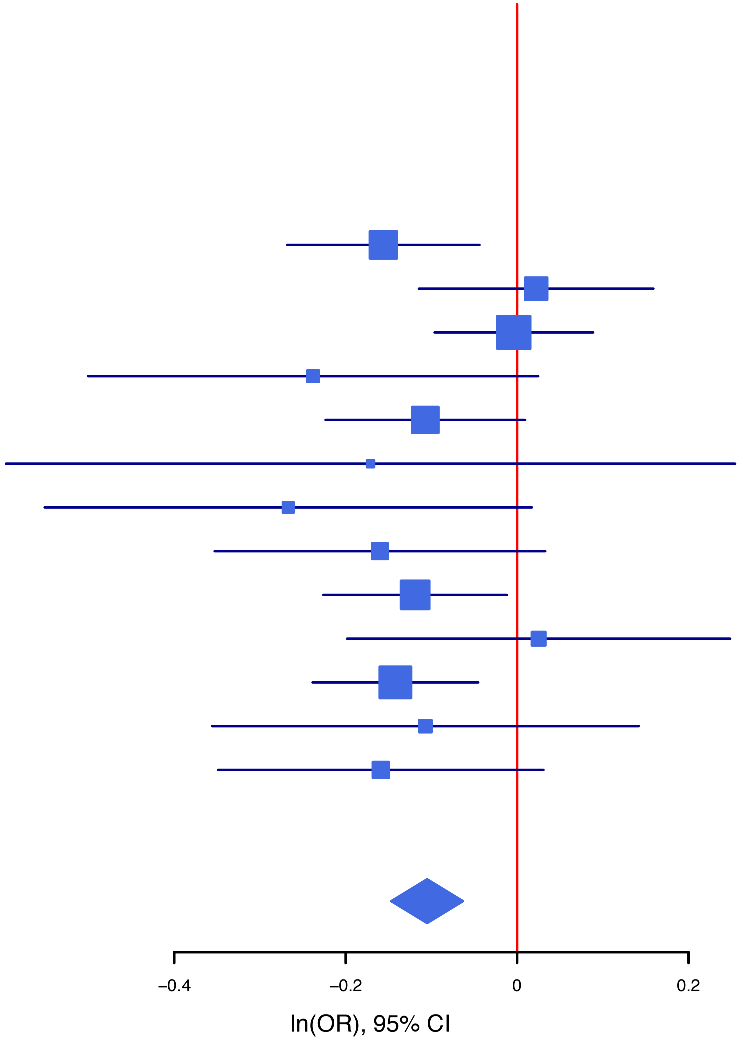

**rs6517397** **A/G** **21:38352125**

het_P: 0.369 het_I: 7.7

| | ngt | info | p_value | f_ca(n) | f_co(n) | ln(OR) | STDerr |
|---|---|---|---|---|---|---|---|
| dan_bvd_grp1−usa1−2_eur | 0 | 0.96 | 0.424 | 0.572(2636) | 0.582(1019) | −0.0434 | 0.0543 |
| dan_bvd_grp1−usa1_eur | 0 | 0.83 | 0.49 | 0.576(1604) | 0.585(1242) | −0.0416 | 0.0602 |
| dan_bvd_grp10−uk1r_eur | 0 | 1.01 | 0.000925 | 0.580(2516) | 0.613(2045) | −0.142 | 0.0429 |
| dan_bvd_grp11−ger2_eur | 0 | 1.02 | 0.715 | 0.541(234) | 0.548(442) | −0.042 | 0.115 |
| dan_bvd_grp12−swes_eur | 0 | 1.00 | 0.286 | 0.544(1273) | 0.557(1544) | −0.0581 | 0.0545 |
| dan_bvd_grp13−usa6_eur | 0 | 0.92 | 0.702 | 0.539(141) | 0.559(101) | −0.0748 | 0.195 |
| dan_bvd_grp2−scot_eur | 0 | 0.85 | 0.323 | 0.590(280) | 0.613(368) | −0.124 | 0.125 |
| dan_bvd_grp3−aus1_eur | 0 | 0.99 | 0.0152 | 0.551(328) | 0.603(1362) | −0.215 | 0.0885 |
| dan_bvd_grp4_ger1_eur | 0 | 1.00 | 0.396 | 0.564(1338) | 0.571(2307) | −0.0419 | 0.0493 |
| dan_bvd_grp5−neth_eur | 0 | 1.01 | 0.00736 | 0.543(737) | 0.612(247) | −0.286 | 0.107 |
| dan_bvd_grp6−usa2_eur | 0 | 0.99 | 0.0387 | 0.577(3426) | 0.598(1496) | −0.093 | 0.045 |
| dan_bvd_grp7−usa3_eur | 0 | 1.00 | 0.0197 | 0.562(375) | 0.619(281) | −0.273 | 0.117 |
| dan_bvd_grp8−usa4_eur | 0 | 0.98 | 0.894 | 0.570(644) | 0.575(466) | −0.0118 | 0.0886 |
| **dan_** | **0** | **0.98** | **5.67e−07** | **0.569(15532)** | **0.587(12920)** | **−0.0916** | **0.0183** |

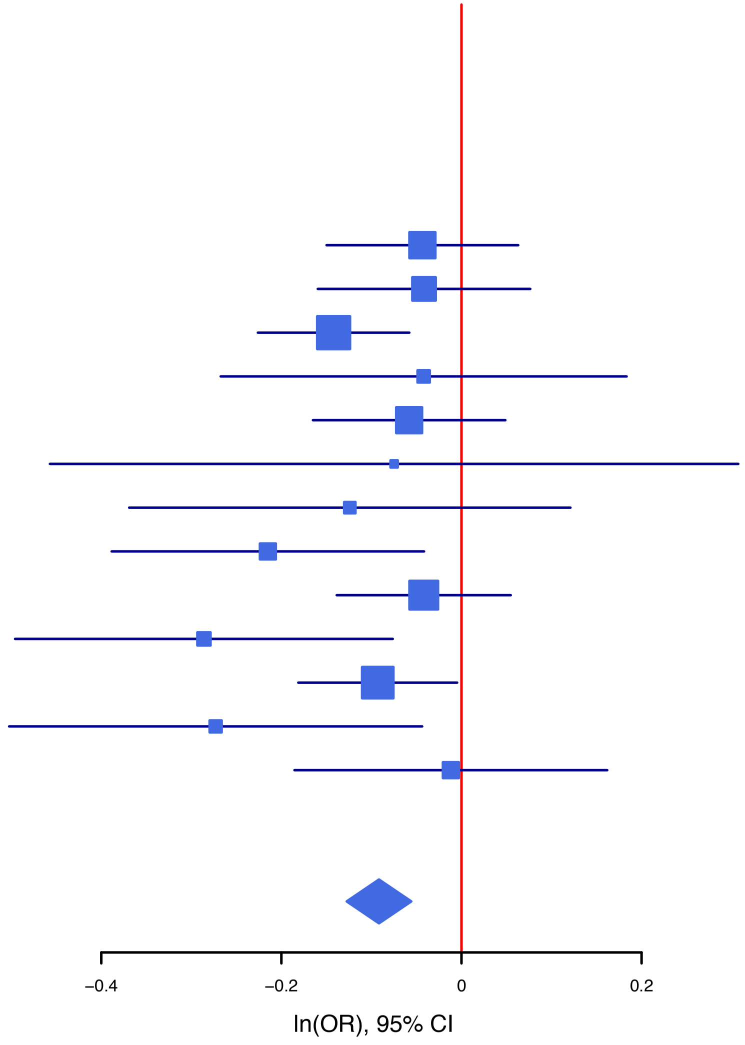

**rs67428741** **A/G** **12:77125630**

+++++++++++++ het_P: 0.579 het_I: 0.0

| | ngt | info | p_value | f_ca(n) | f_co(n) | ln(OR) | STDerr |
|---|---|---|---|---|---|---|---|
| dan_bvd_grp1−usa1−2_eur | 0 | 0.91 | 0.0356 | 0.783(2636) | 0.762(1019) | 0.137 | 0.0652 |
| dan_bvd_grp1−usa1_eur | 0 | 0.85 | 0.723 | 0.781(1604) | 0.777(1242) | 0.0249 | 0.0703 |
| dan_bvd_grp10−uk1r_eur | 0 | 0.96 | 0.0544 | 0.783(2516) | 0.767(2045) | 0.0996 | 0.0518 |
| dan_bvd_grp11−ger2_eur | 0 | 0.91 | 0.76 | 0.784(234) | 0.778(442) | 0.0448 | 0.146 |
| dan_bvd_grp12−swes_eur | 0 | 0.94 | 0.196 | 0.791(1273) | 0.776(1544) | 0.0875 | 0.0677 |
| dan_bvd_grp13−usa6_eur | 0 | 1.02 | 0.545 | 0.797(141) | 0.778(101) | 0.138 | 0.228 |
| dan_bvd_grp2−scot_eur | 0 | 0.94 | 0.948 | 0.783(280) | 0.780(368) | 0.00926 | 0.141 |
| dan_bvd_grp3−aus1_eur | 0 | 0.96 | 0.00754 | 0.827(328) | 0.782(1362) | 0.312 | 0.117 |
| dan_bvd_grp4_ger1_eur | 0 | 0.93 | 0.00297 | 0.805(1338) | 0.778(2307) | 0.188 | 0.0633 |
| dan_bvd_grp5−neth_eur | 0 | 0.97 | 0.0134 | 0.795(737) | 0.745(247) | 0.309 | 0.125 |
| dan_bvd_grp6−usa2_eur | 0 | 0.92 | 0.0605 | 0.778(3426) | 0.762(1496) | 0.102 | 0.0542 |
| dan_bvd_grp7−usa3_eur | 0 | 0.94 | 0.995 | 0.764(375) | 0.765(281) | 0.0009 | 0.138 |
| dan_bvd_grp8−usa4_eur | 0 | 0.93 | 0.593 | 0.782(644) | 0.774(466) | 0.0576 | 0.108 |
| **dan_** | **0** | **0.93** | **3.68e−07** | **0.785(15532)** | **0.772(12920)** | **0.113** | **0.0223** |

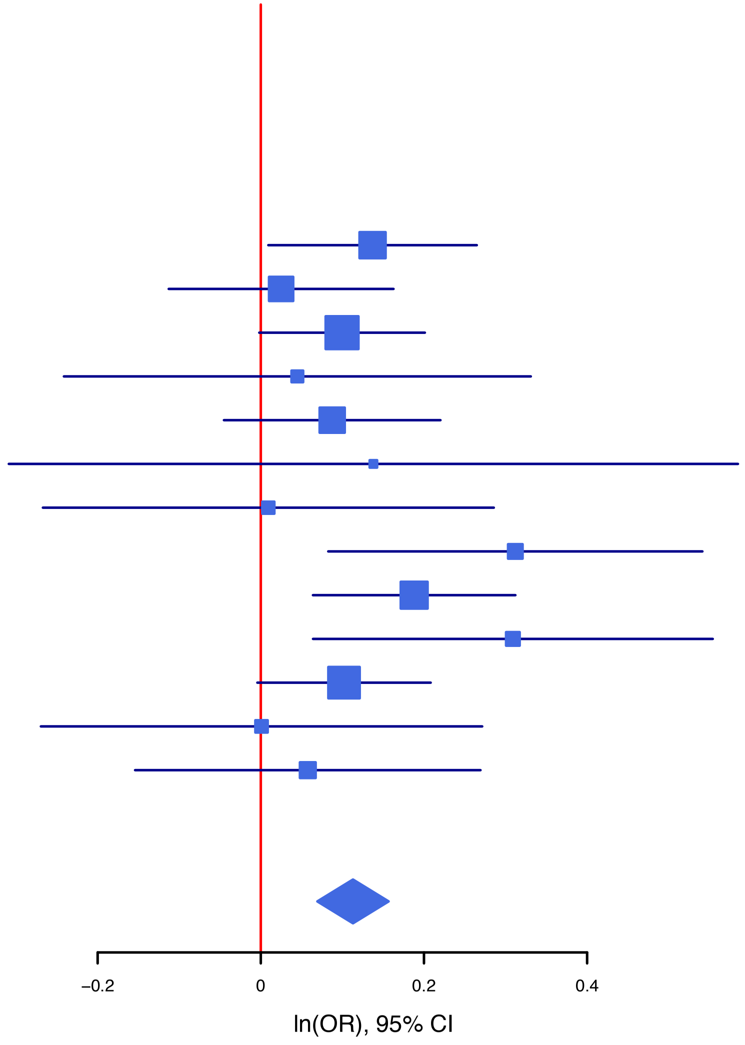

**rs75184229** **C/G** **11:85517549**

+++−+++++−+++ het_P: 0.78 het_I: 0.0

| | ngt | info | p_value | f_ca(n) | f_co(n) | ln(OR) | STDerr |
|---|---|---|---|---|---|---|---|
| dan_bvd_grp1−usa1−2_eur | 0 | 0.97 | 0.00057 | 0.966(2636) | 0.947(1019) | 0.44 | 0.128 |
| dan_bvd_grp1−usa1_eur | 0 | 0.97 | 0.349 | 0.963(1604) | 0.960(1242) | 0.134 | 0.143 |
| dan_bvd_grp10−uk1r_eur | 0 | 0.97 | 0.0584 | 0.971(2516) | 0.964(2045) | 0.23 | 0.121 |
| dan_bvd_grp11−ger2_eur | 0 | 1.04 | 0.0672 | 0.968(234) | 0.945(442) | 0.547 | 0.299 |
| dan_bvd_grp12−swes_eur | 0 | 0.96 | 0.0365 | 0.973(1273) | 0.964(1544) | 0.336 | 0.161 |
| dan_bvd_grp13−usa6_eur | 0 | 0.97 | 0.906 | 0.924(141) | 0.935(101) | −0.0452 | 0.383 |
| dan_bvd_grp2−scot_eur | 0 | 0.98 | 0.276 | 0.967(280) | 0.954(368) | 0.326 | 0.299 |
| dan_bvd_grp3−aus1_eur | 0 | 0.99 | 0.517 | 0.966(328) | 0.960(1362) | 0.154 | 0.237 |
| dan_bvd_grp4_ger1_eur | 0 | 0.99 | 0.471 | 0.962(1338) | 0.959(2307) | 0.0914 | 0.127 |
| dan_bvd_grp5−neth_eur | 0 | 0.95 | 0.903 | 0.966(737) | 0.967(247) | −0.036 | 0.297 |
| dan_bvd_grp6−usa2_eur | 0 | 0.95 | 0.00509 | 0.967(3426) | 0.956(1496) | 0.322 | 0.115 |
| dan_bvd_grp7−usa3_eur | 0 | 0.99 | 0.598 | 0.976(375) | 0.966(281) | 0.182 | 0.345 |
| dan_bvd_grp8−usa4_eur | 0 | 0.95 | 0.439 | 0.961(644) | 0.953(466) | 0.17 | 0.22 |
| **dan_** | **0** | **0.97** | **2.17e−07** | **0.967(15532)** | **0.959(12920)** | **0.247** | **0.0476** |

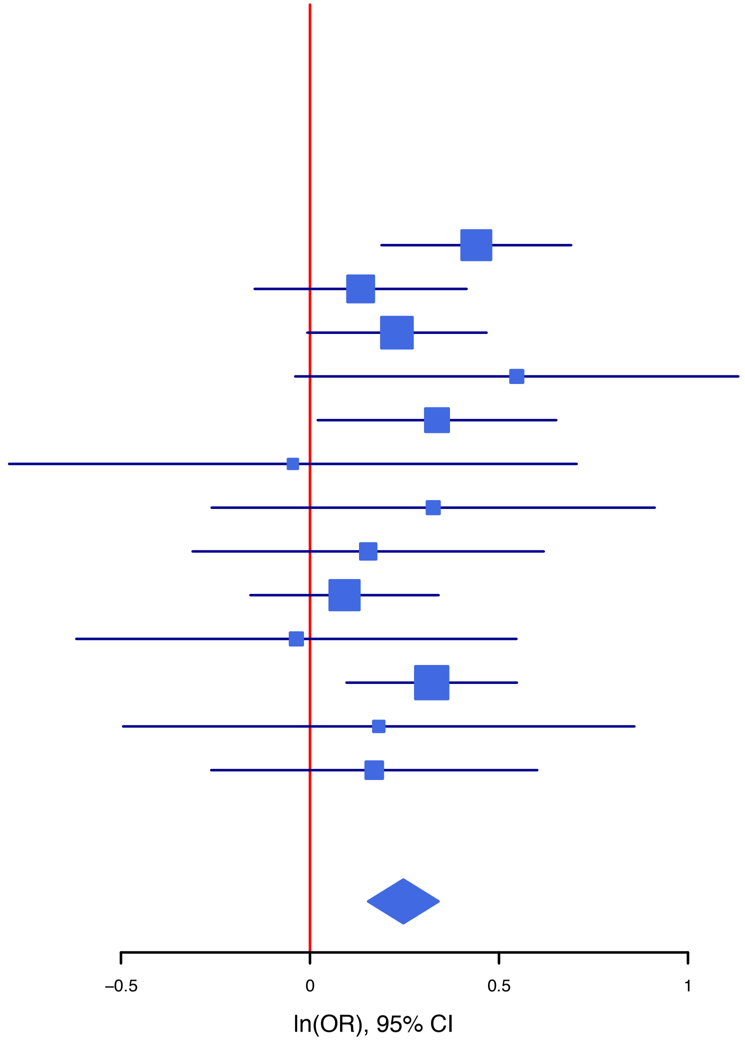
