## Supplementary figures and images for "Identifying genetic differences between bipolar disorder and major depression through multiple GWAS"

### SF1a.BDvsMDD_PCA.pdf

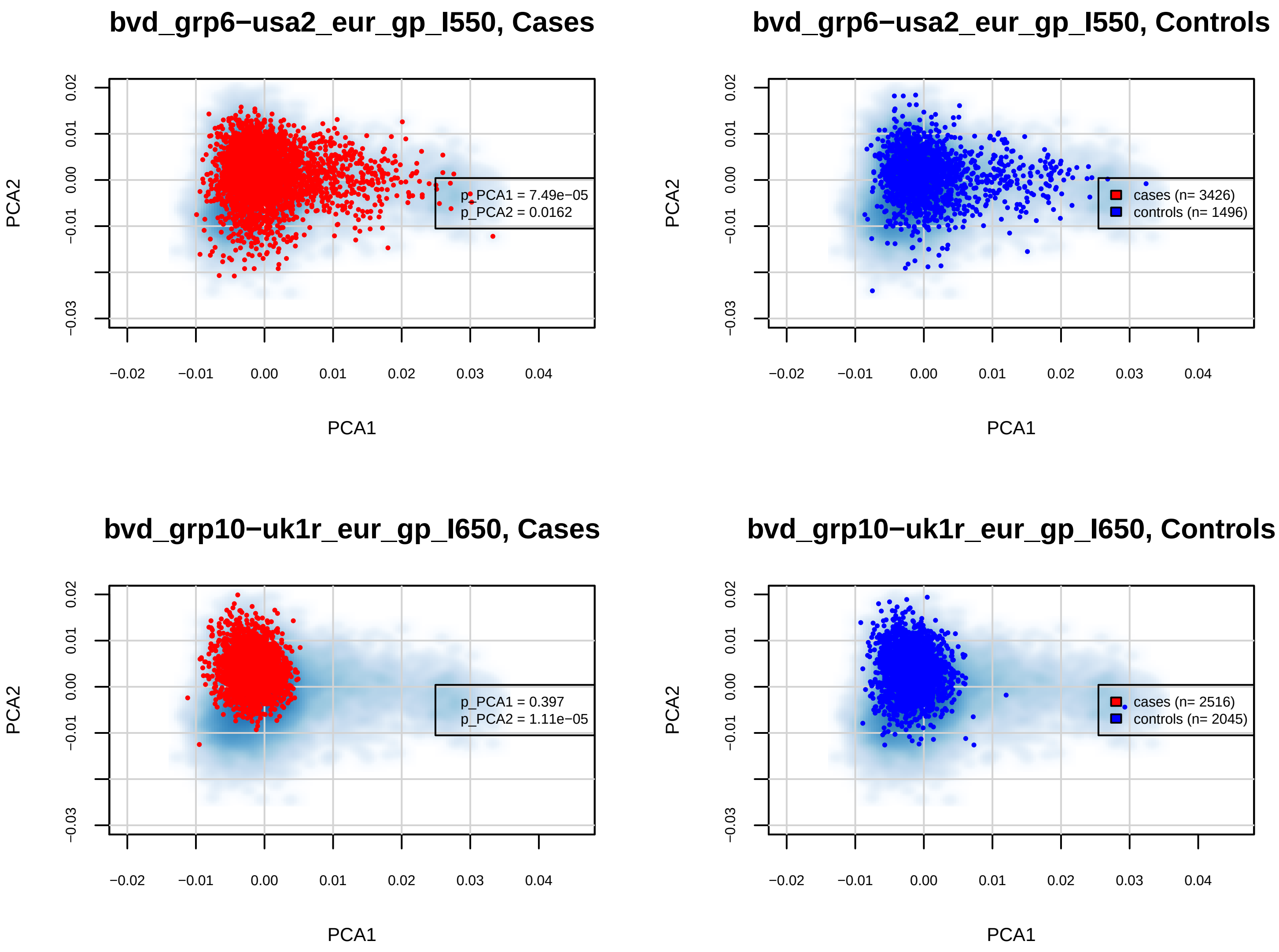

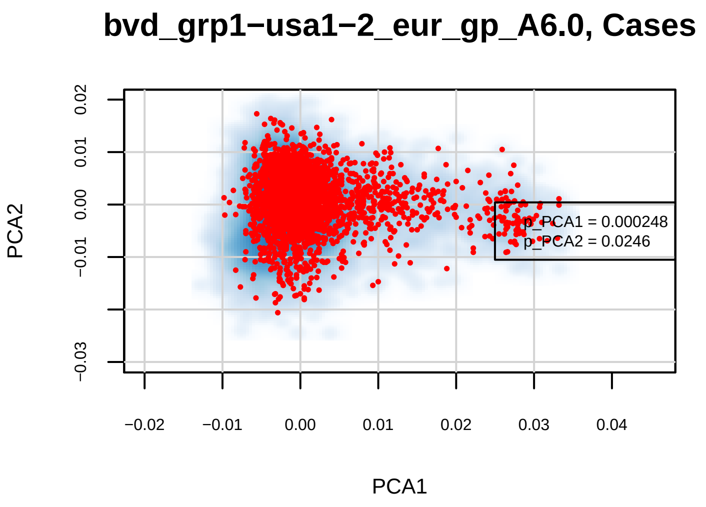

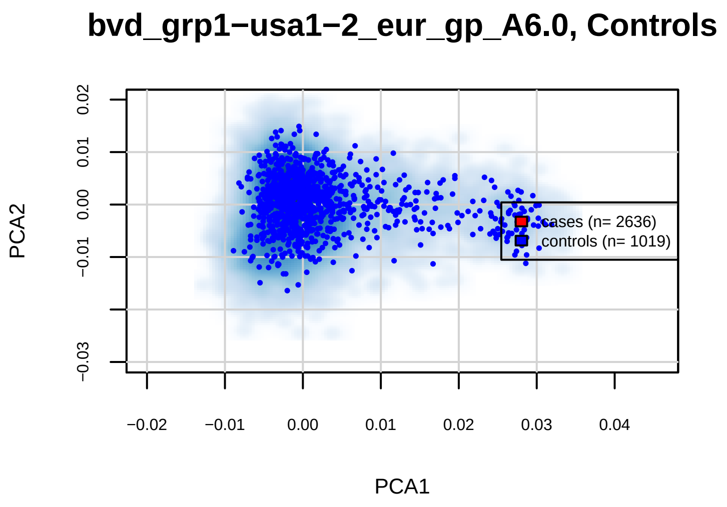

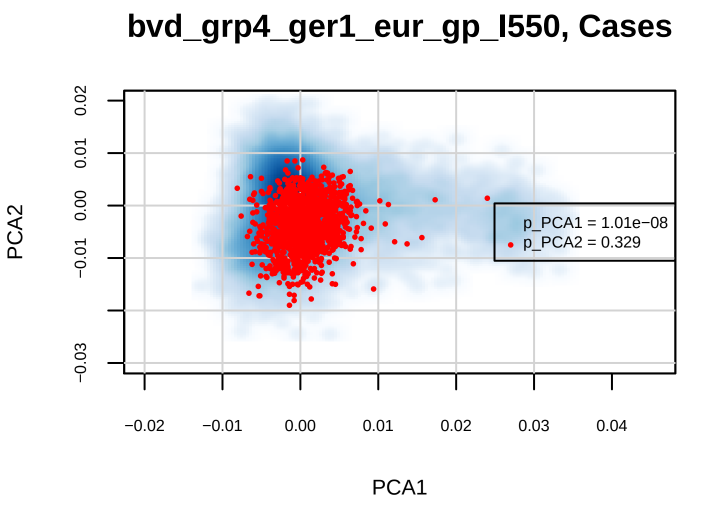

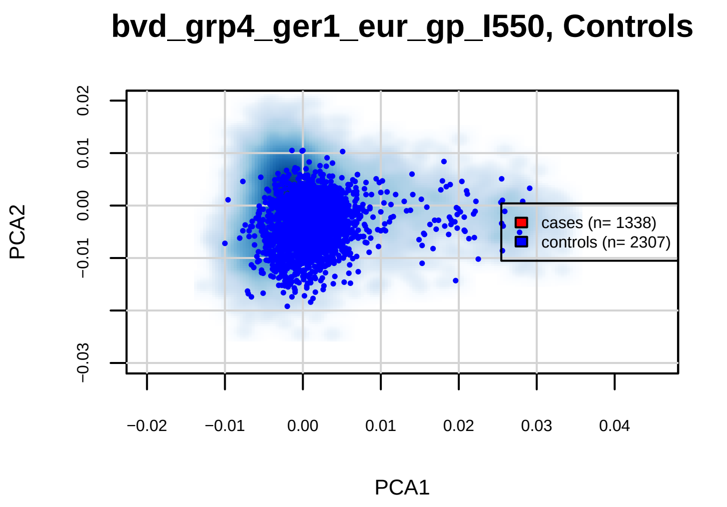

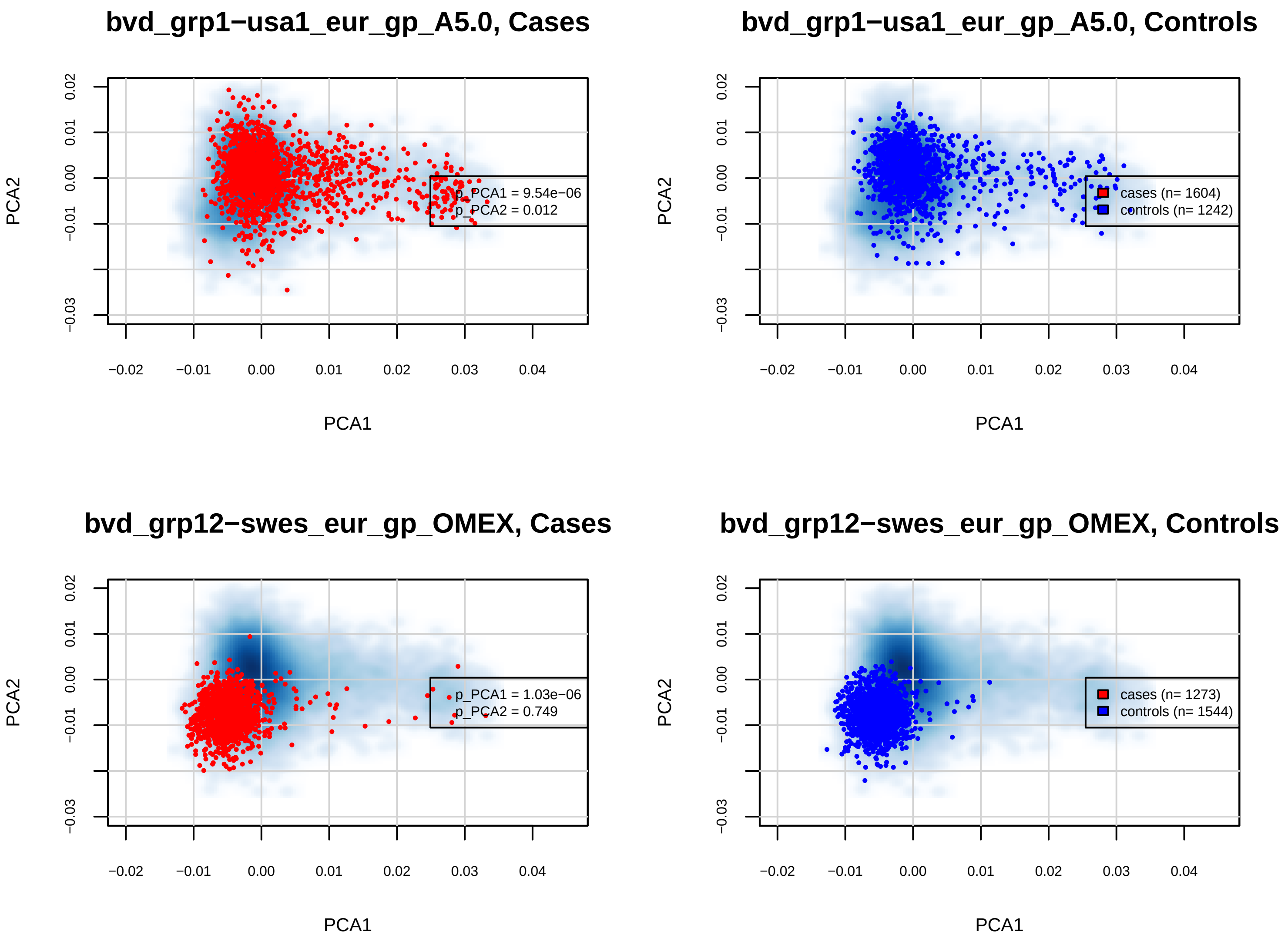

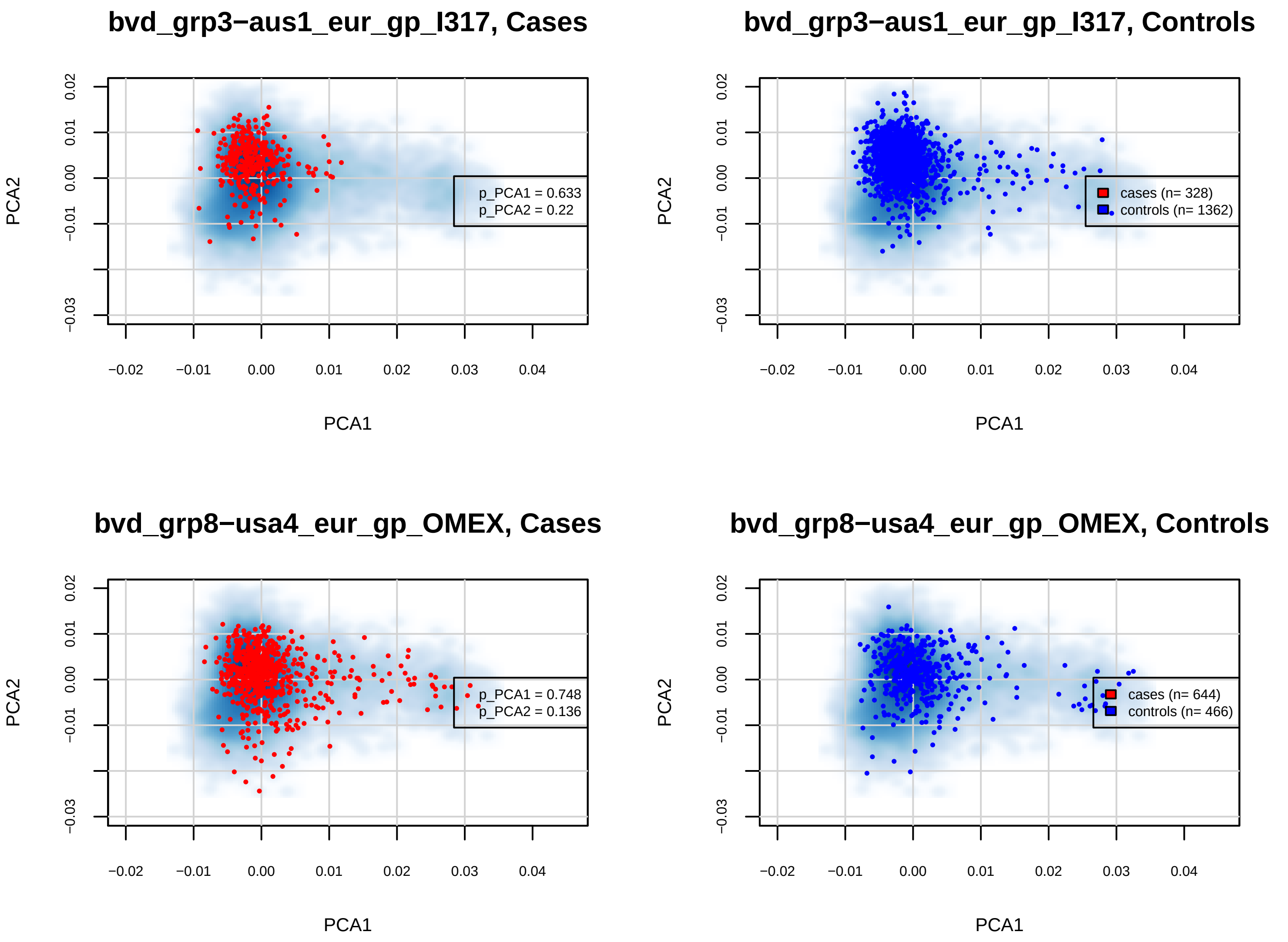

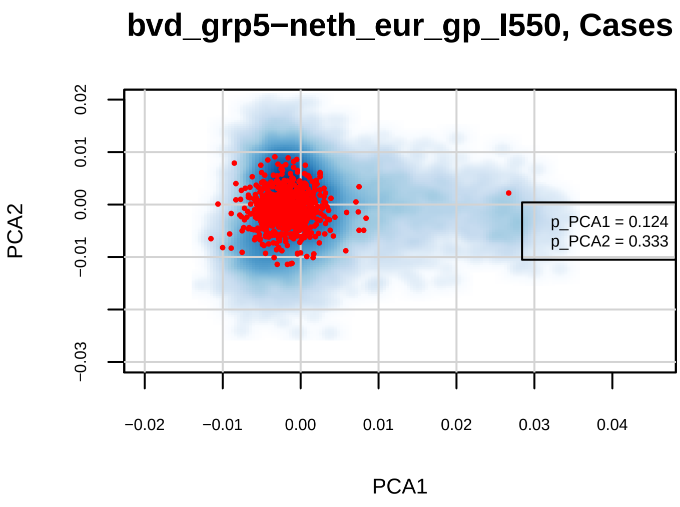

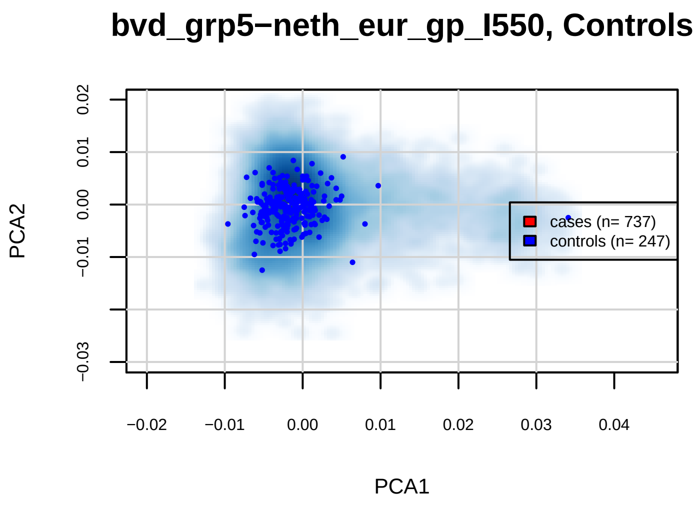

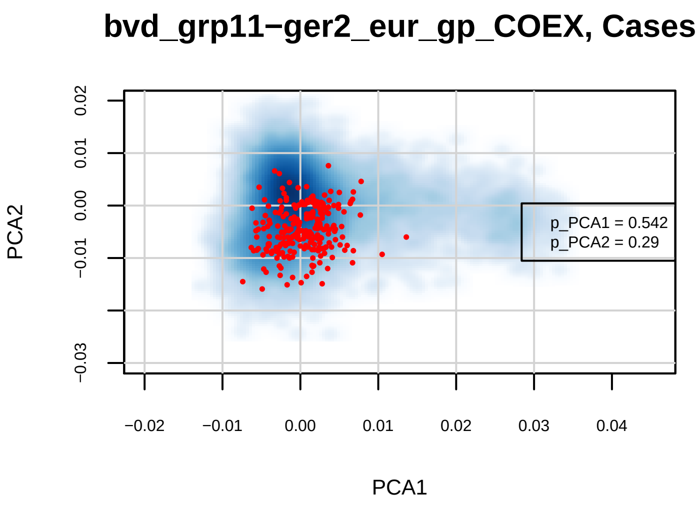

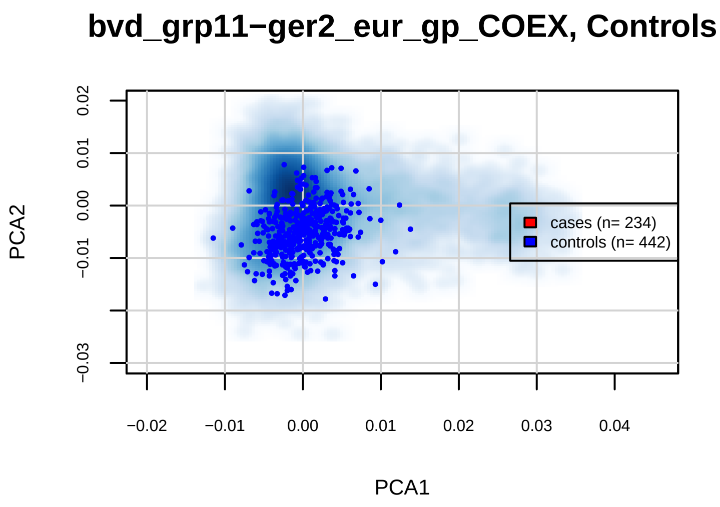

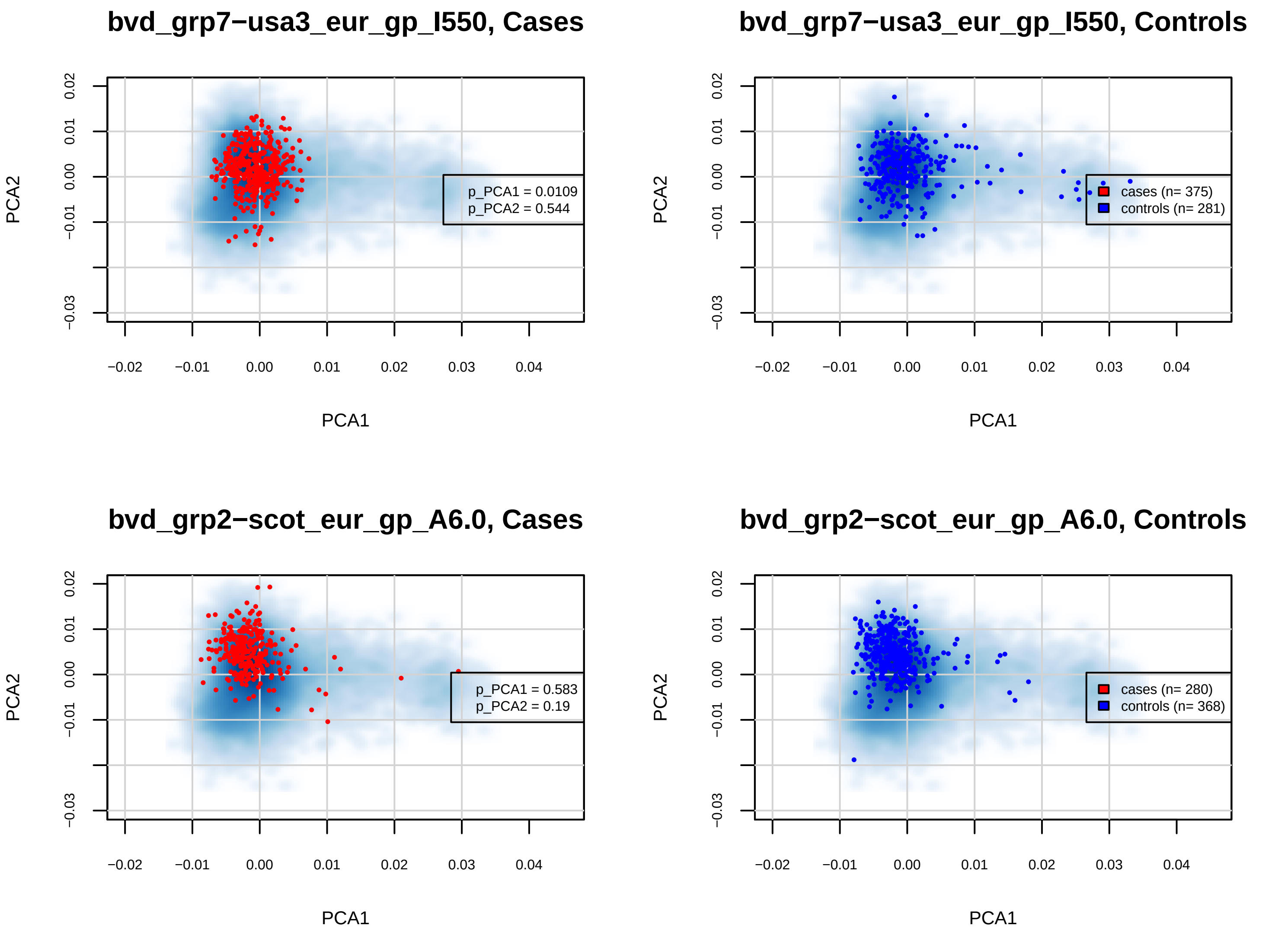

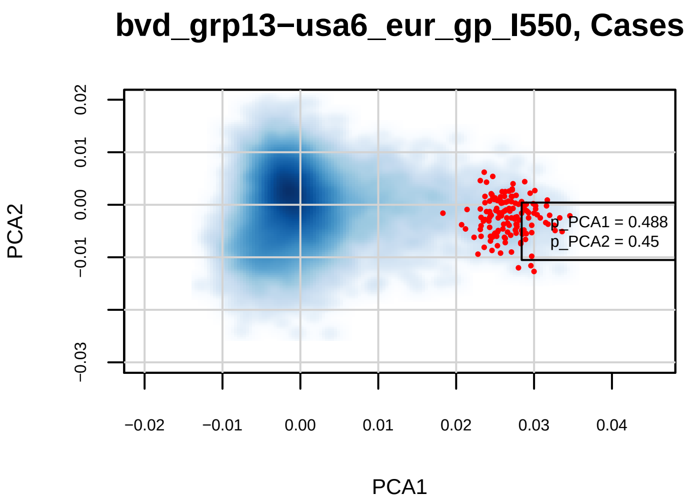

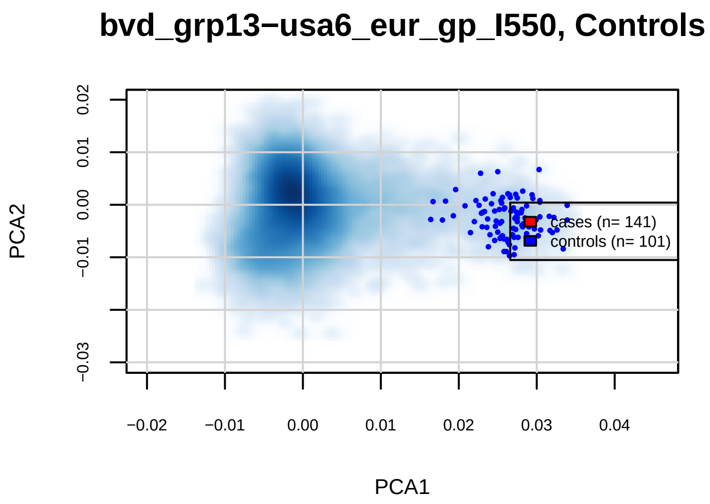

### SF1b.BDvsMDDvsCon_PCA.pdf

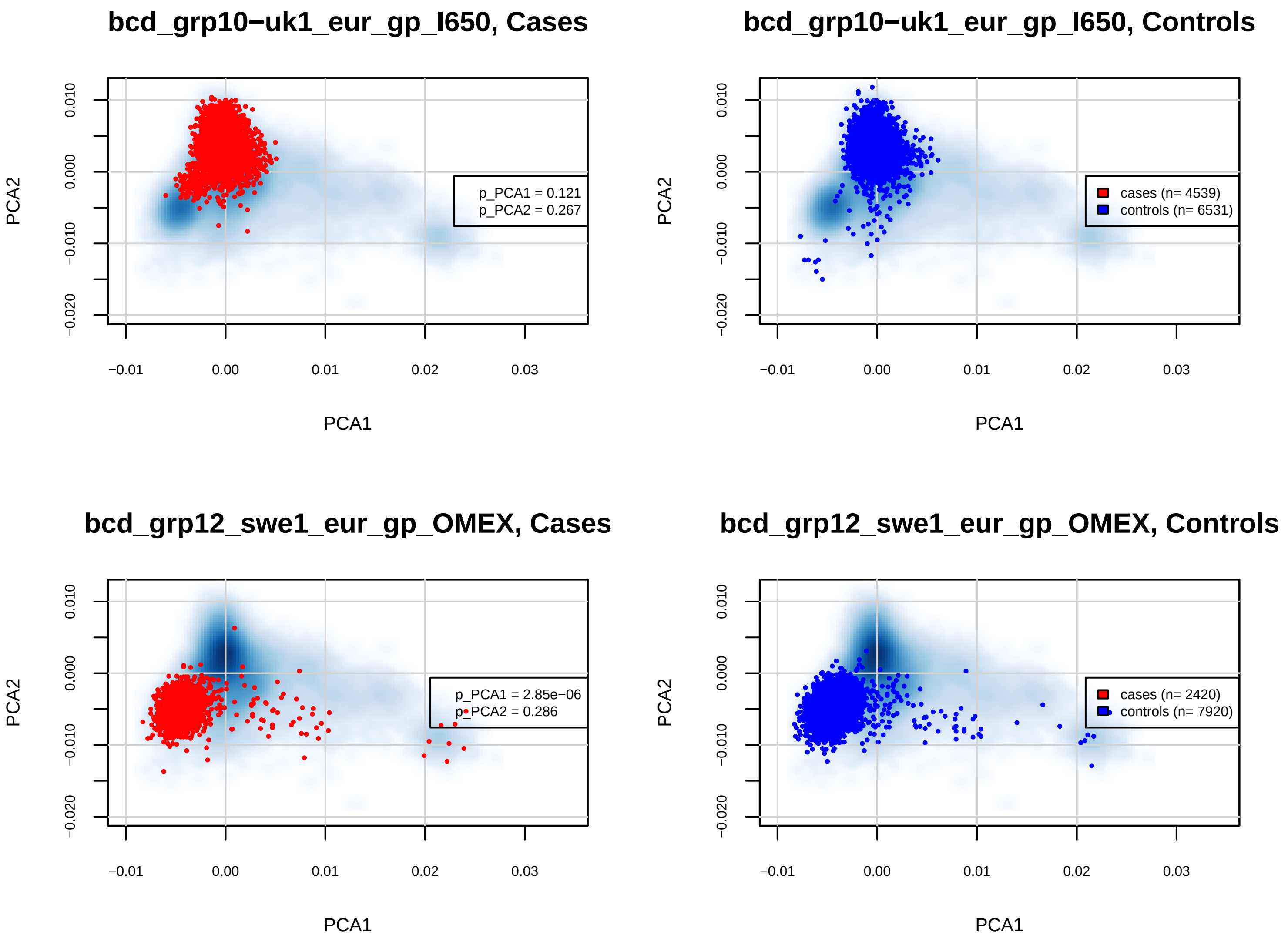

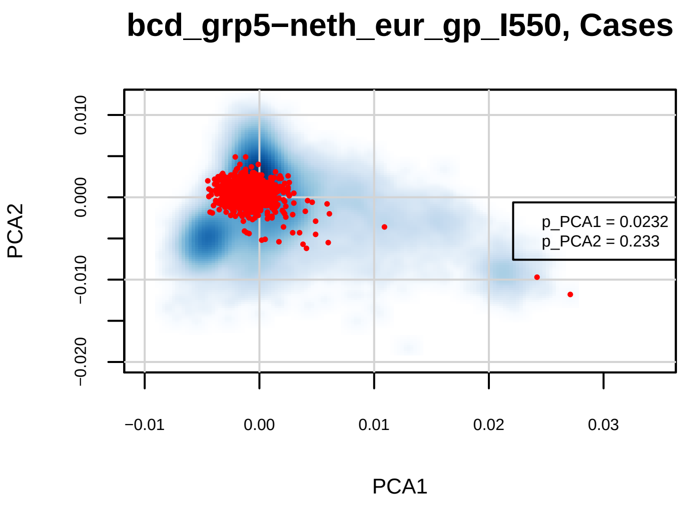

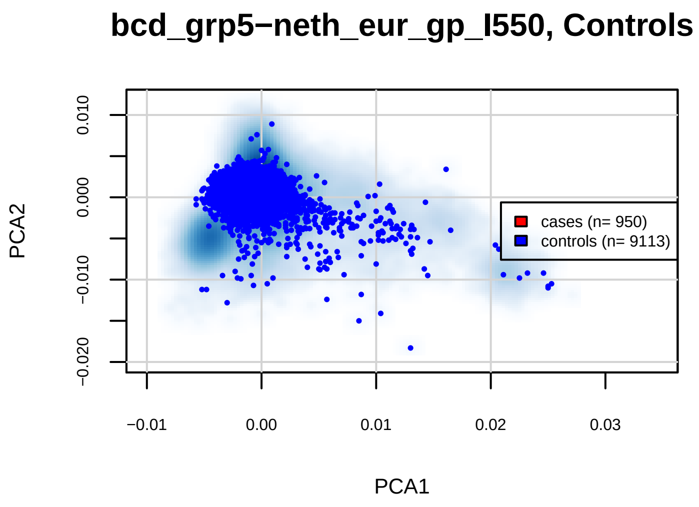

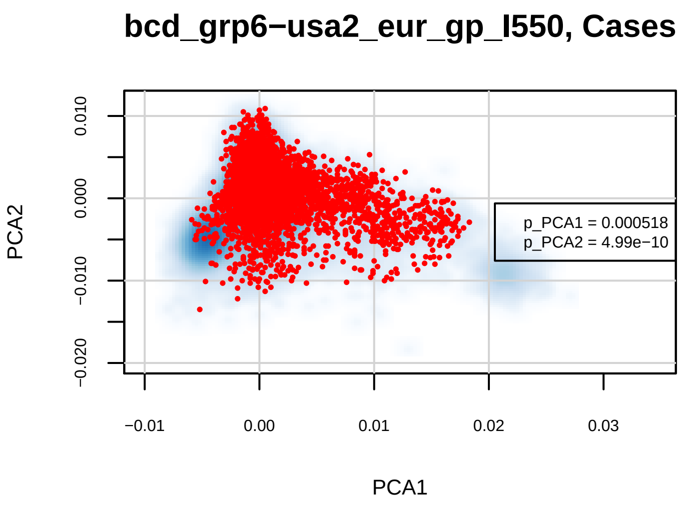

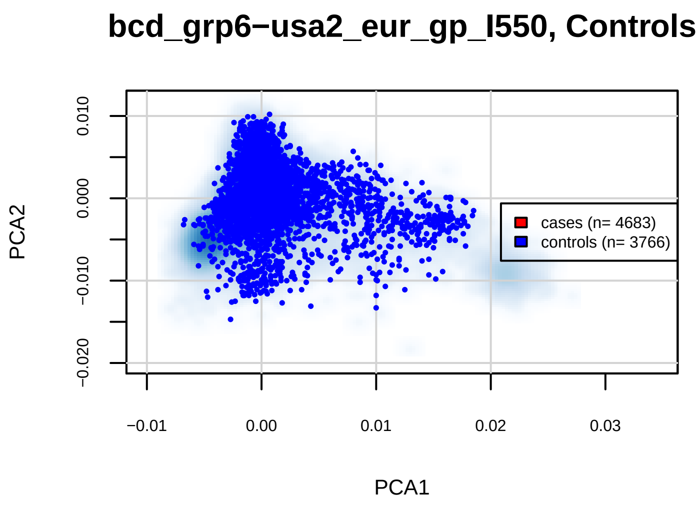

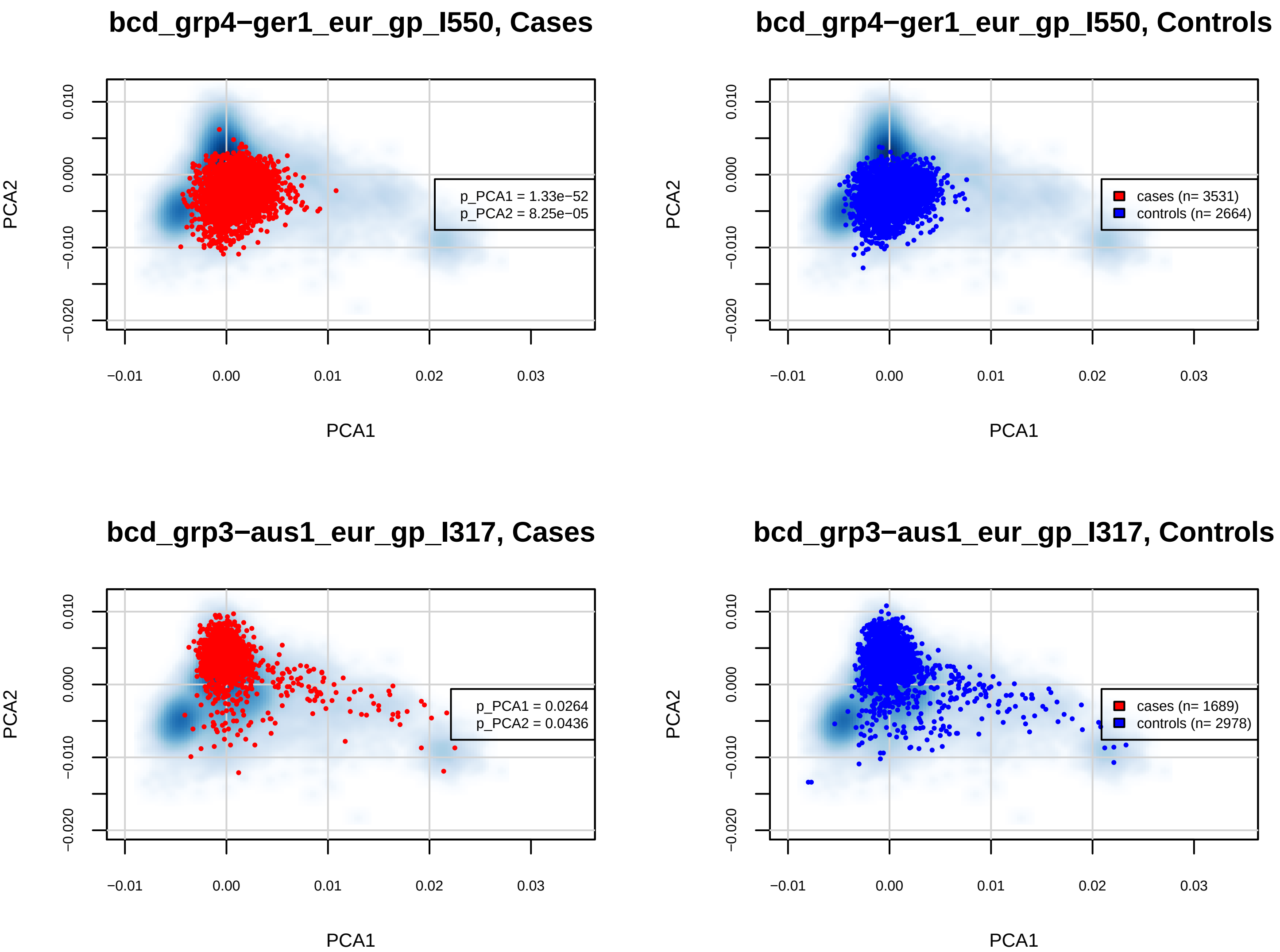

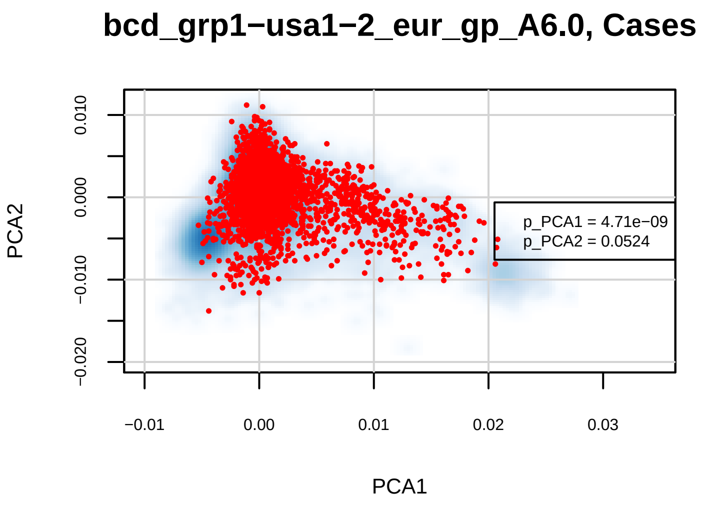

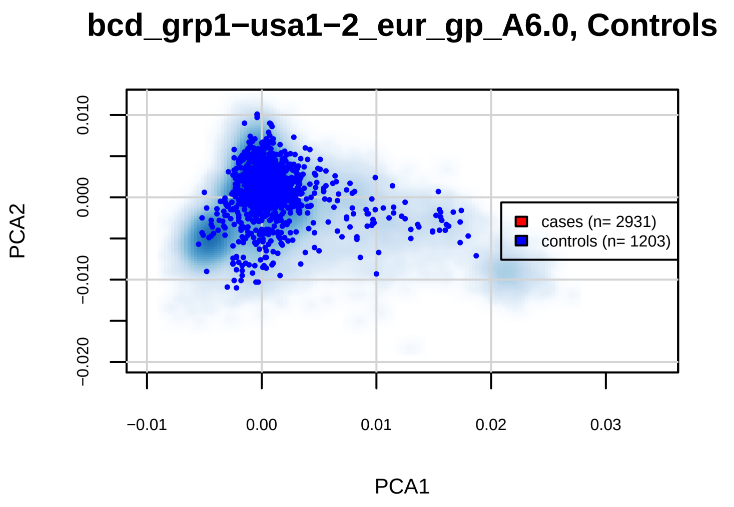

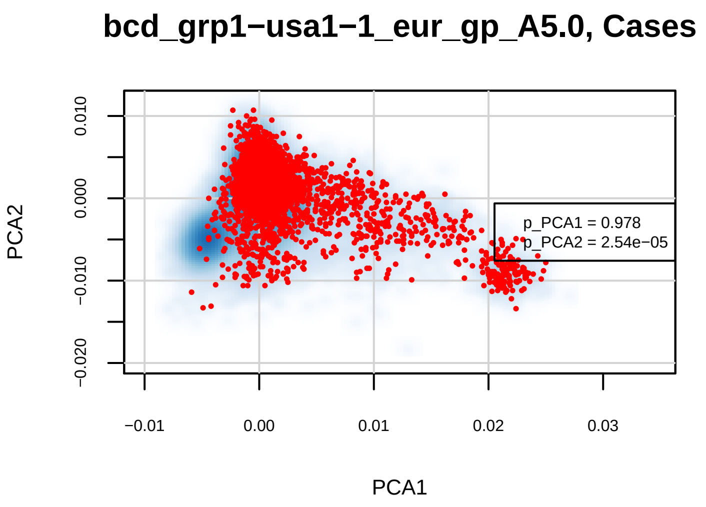

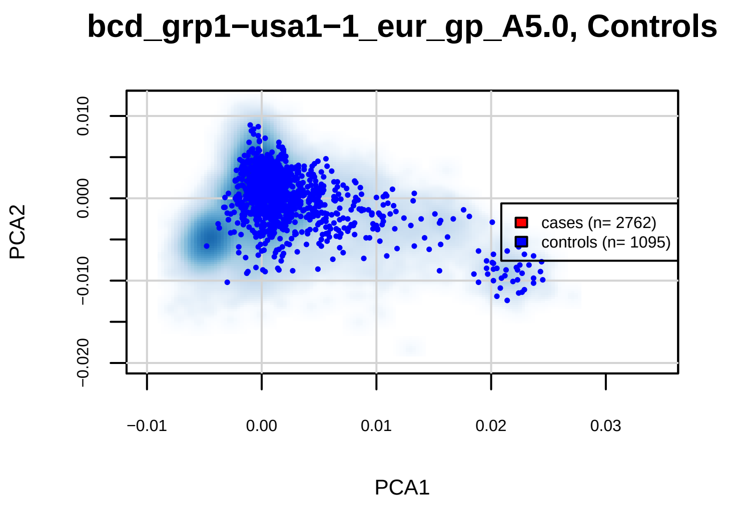

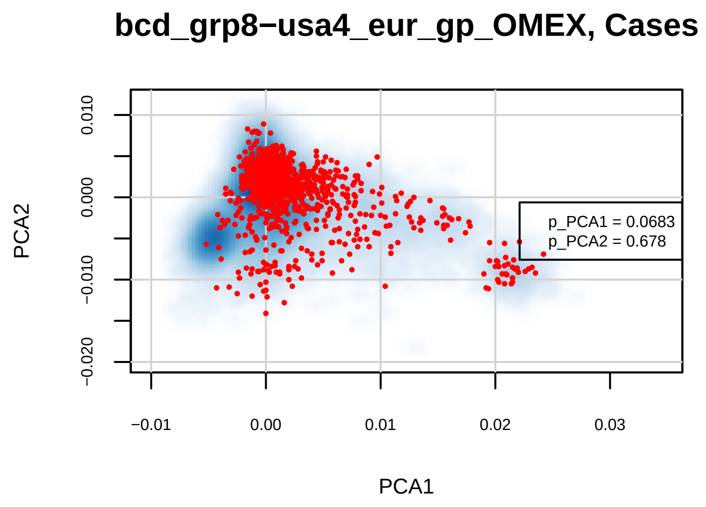

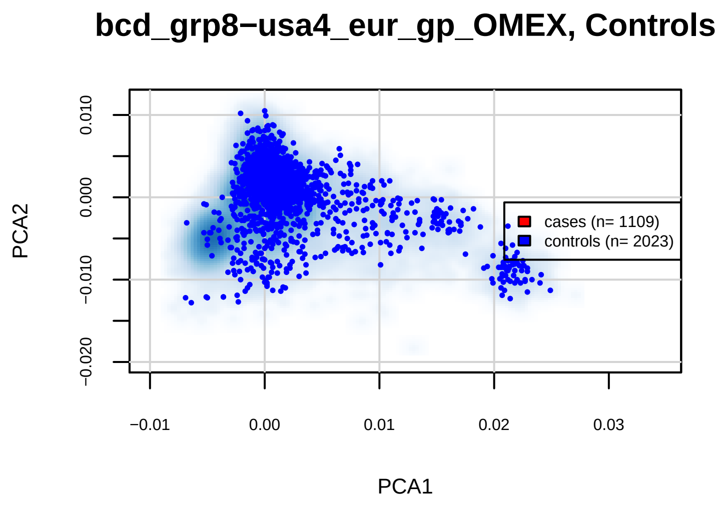

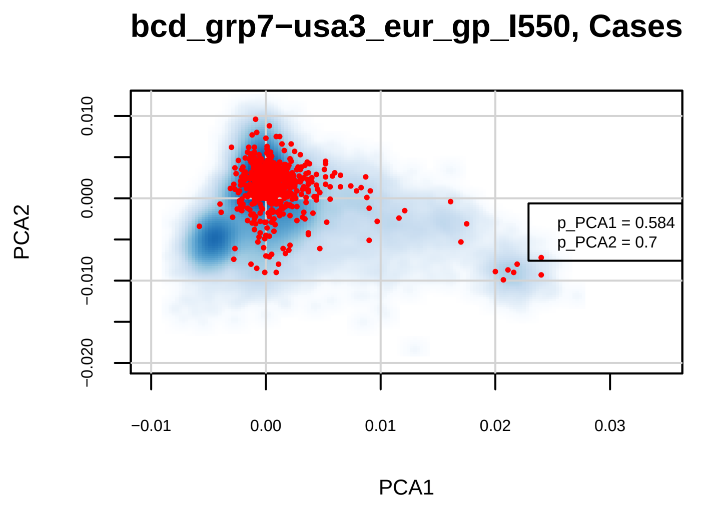

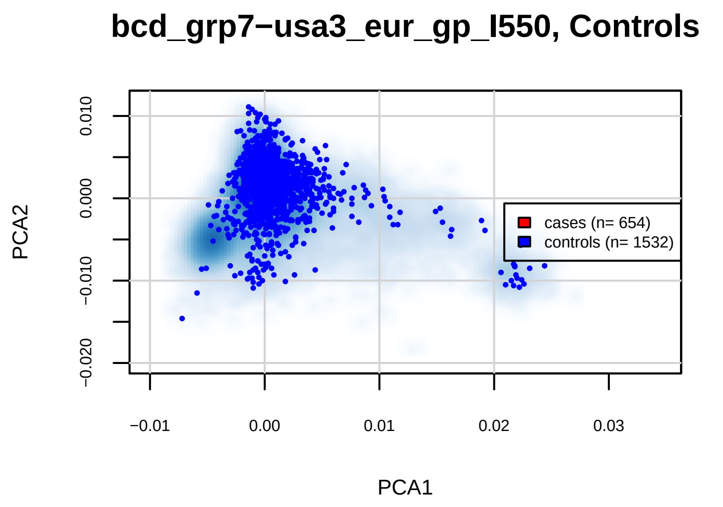

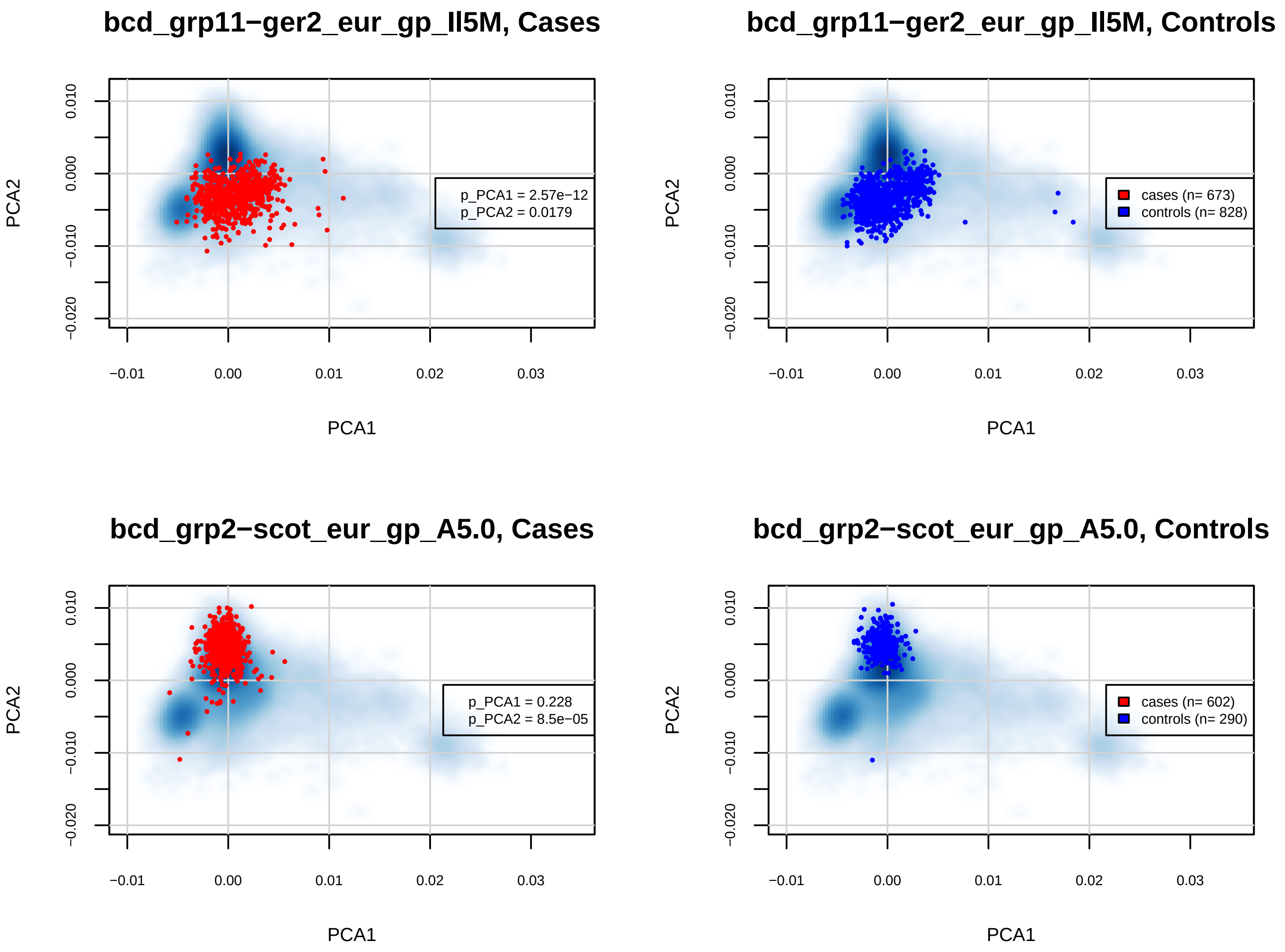

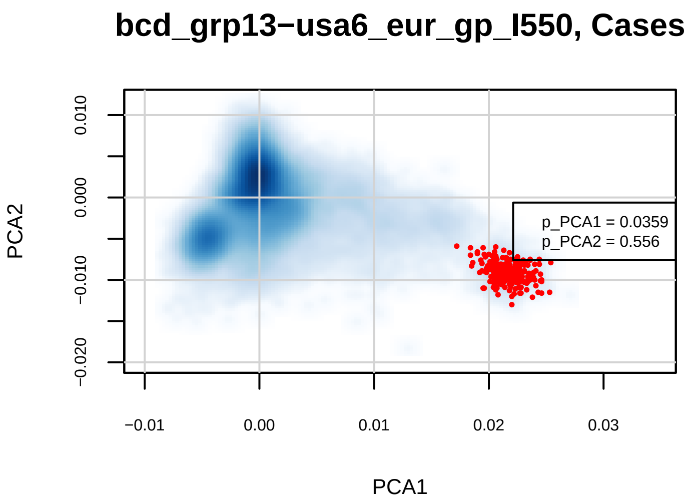

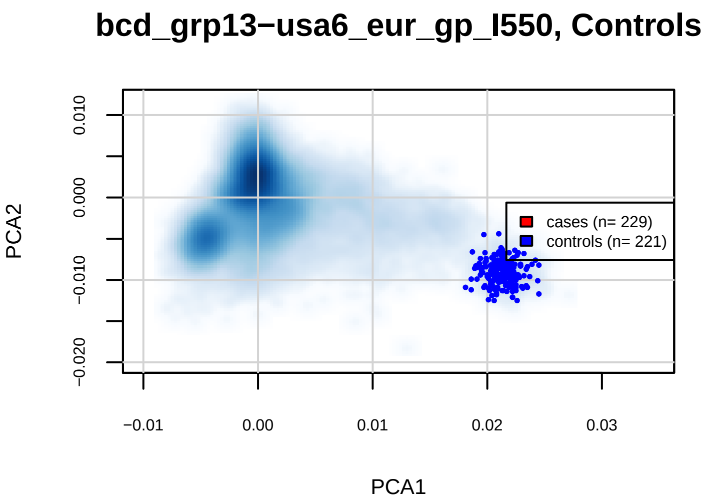

### SF2a.manhattan.v2.BDvsMDD.pdf

#### daner_BIP−MDD_13_run2.gz.p3_GWA

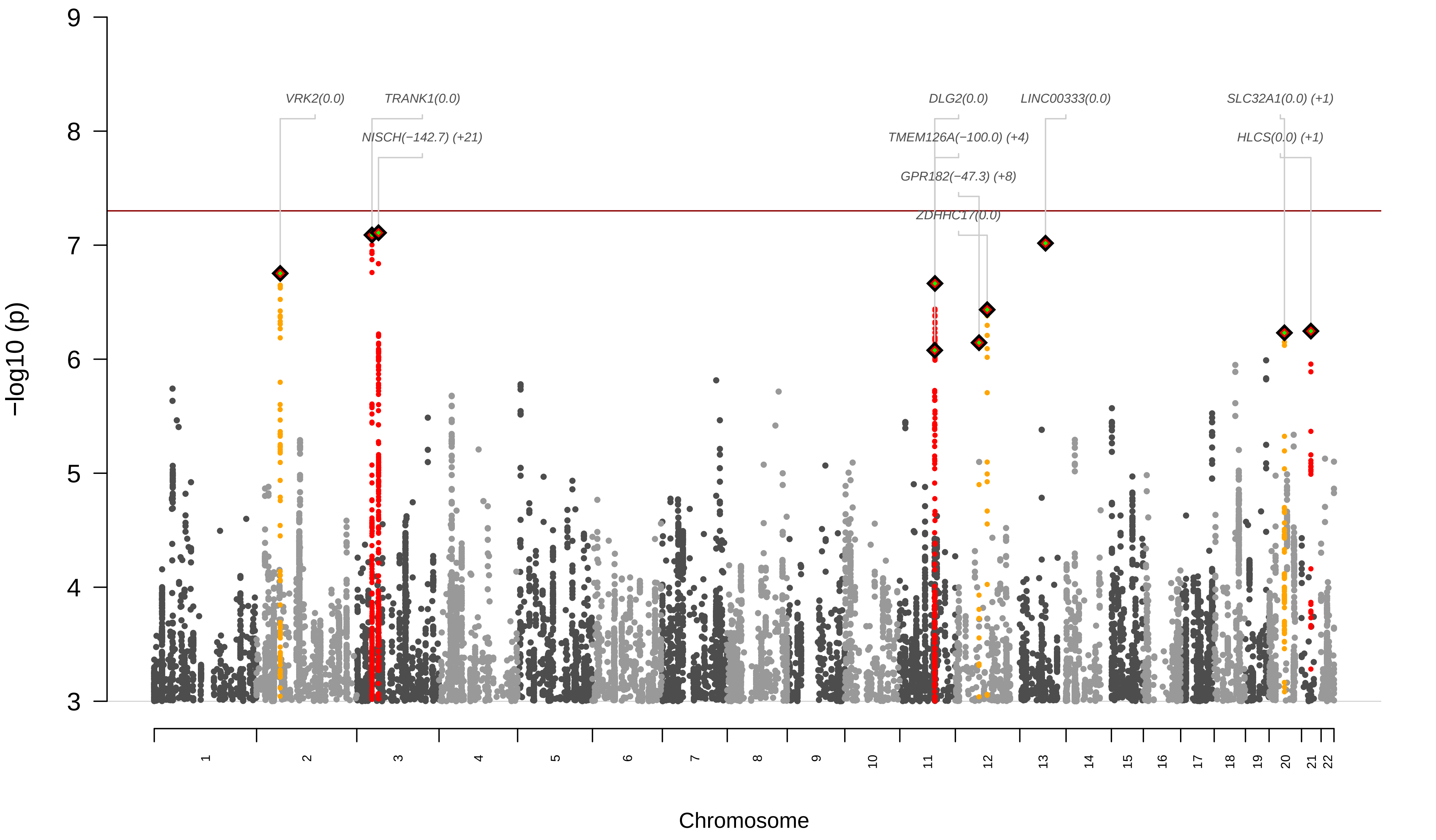

### SF2b.manhattan.v2.BD-DvsMDD.pdf

#### daner_MDD_BIP−D_run2_0519.gz.p3_GWA

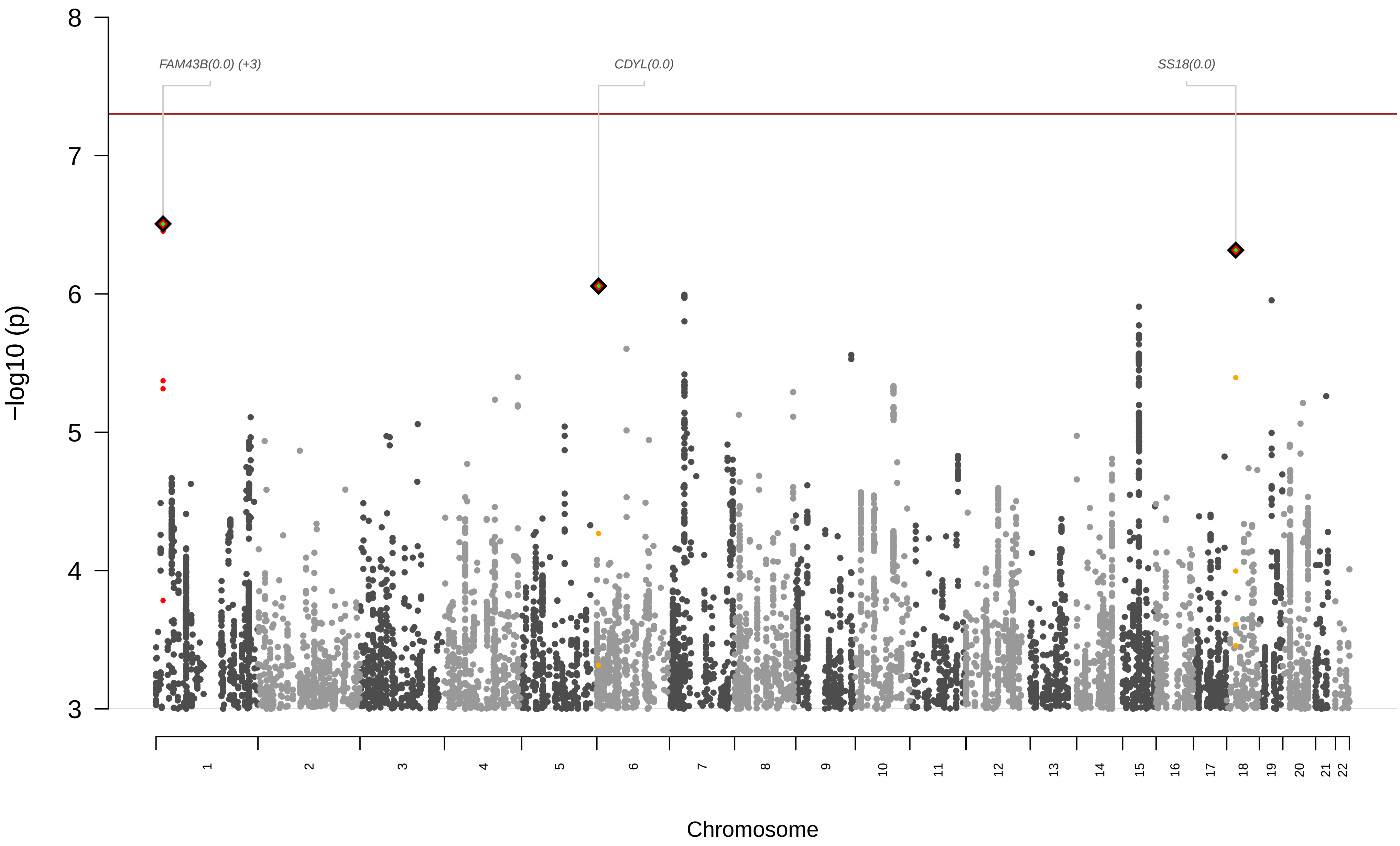

### SF2c.manhattan.v2.MetaRegr.pdf

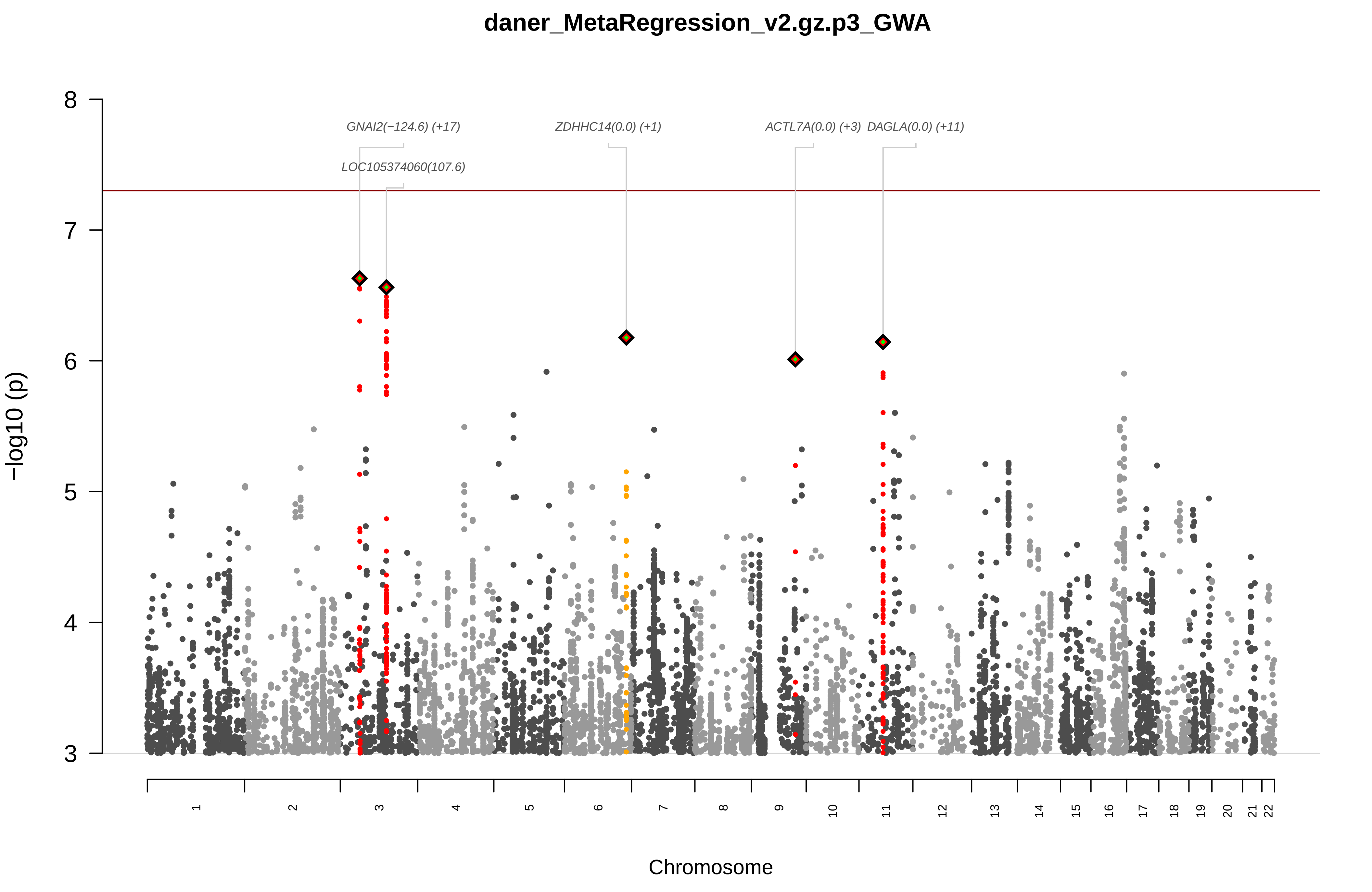

### SF2d.manhattan.v2.CCGWAS.pdf

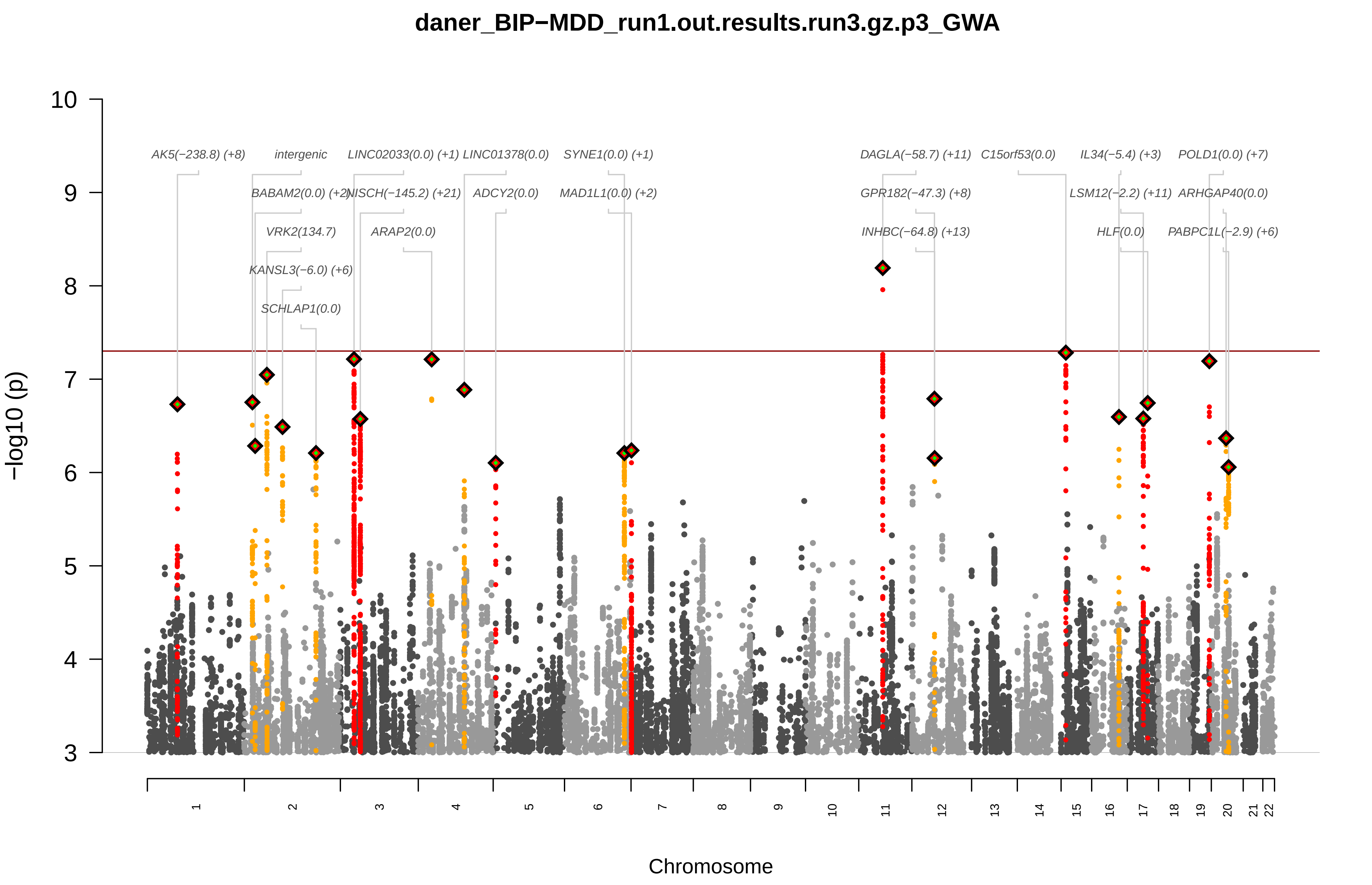

### SF3a.qq.BDvsMDD.pdf

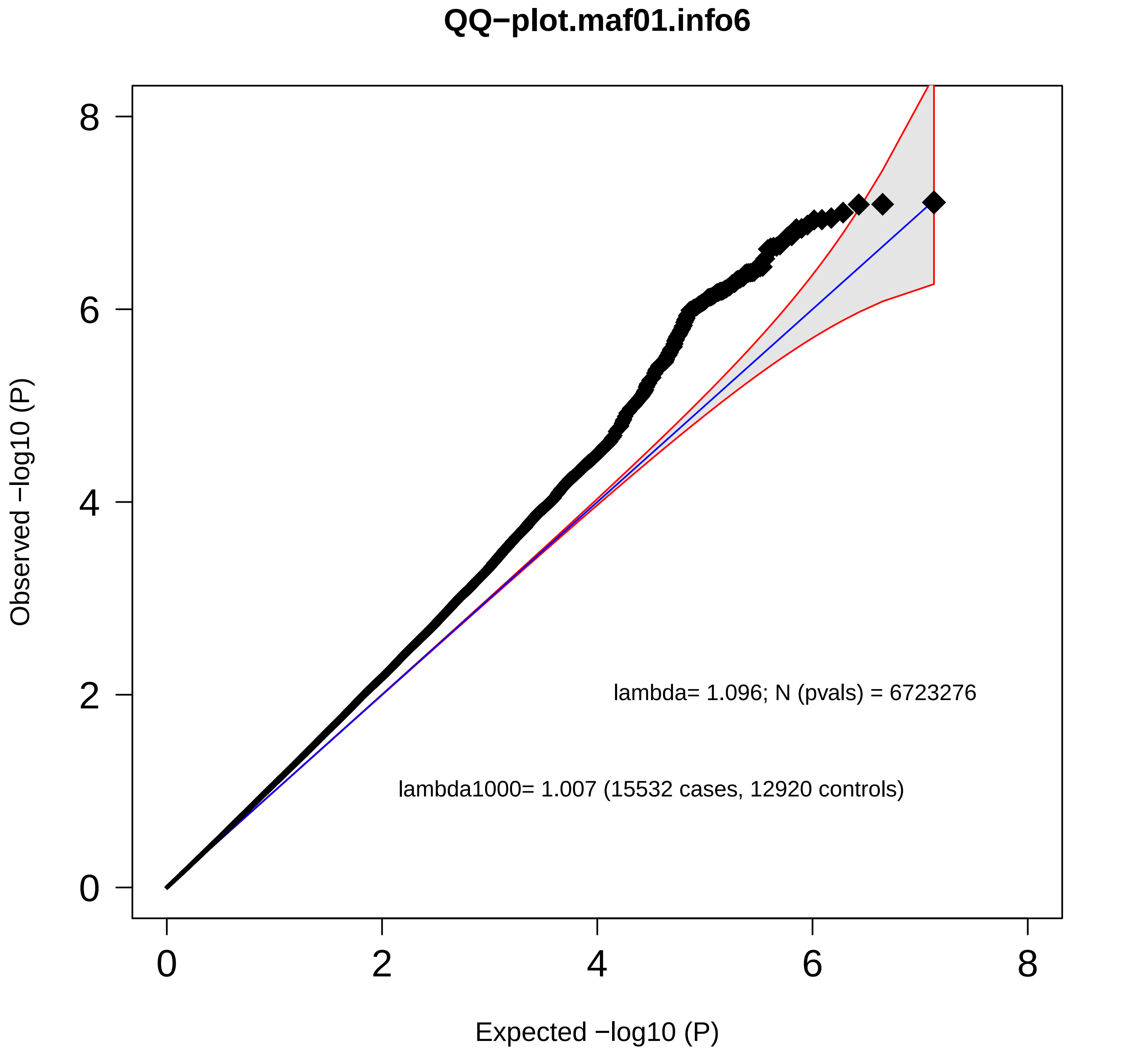

### SF3b.qq.BD-DvsMDD.pdf

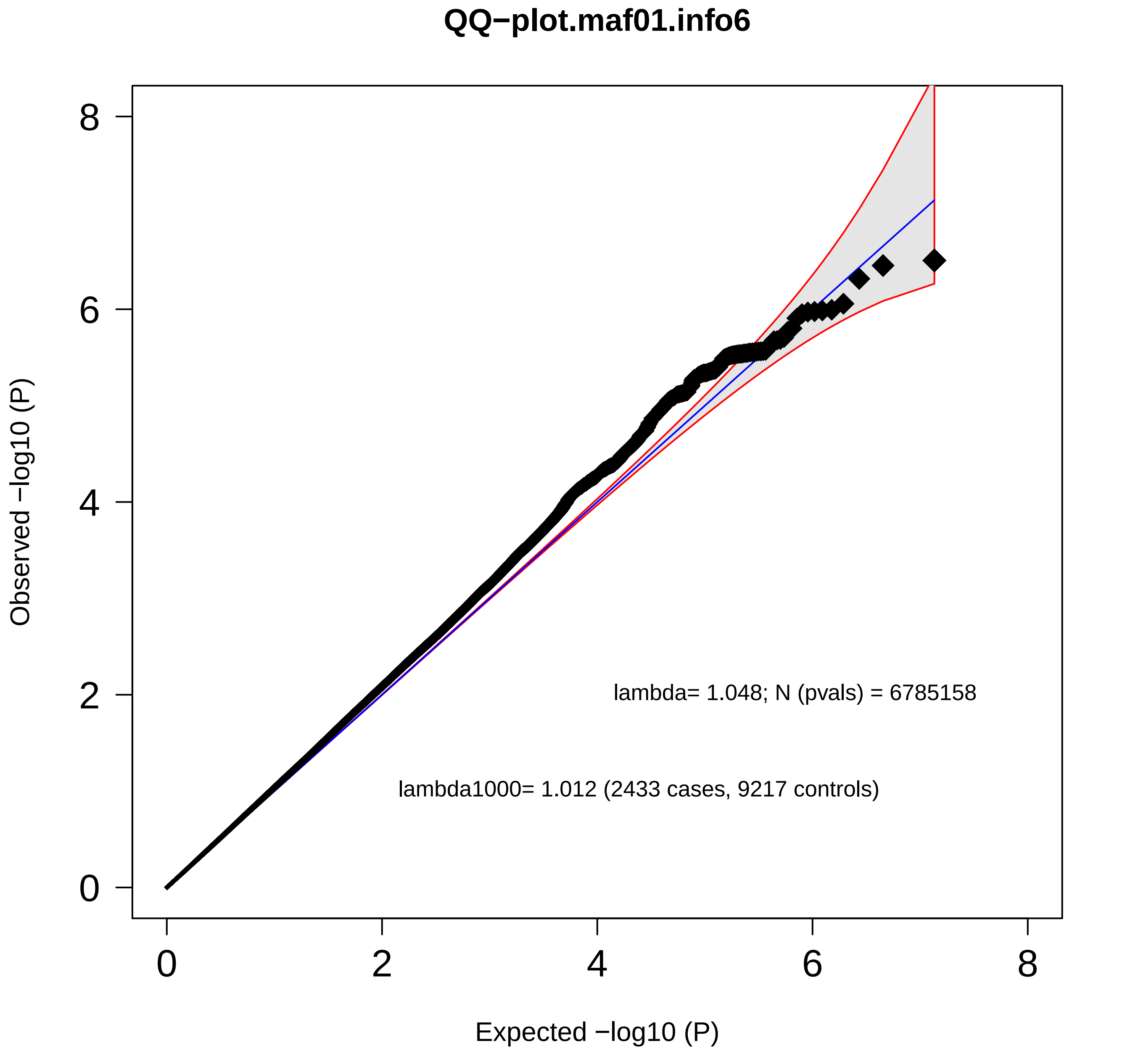

### SF3c.qq.MetaRegr.pdf

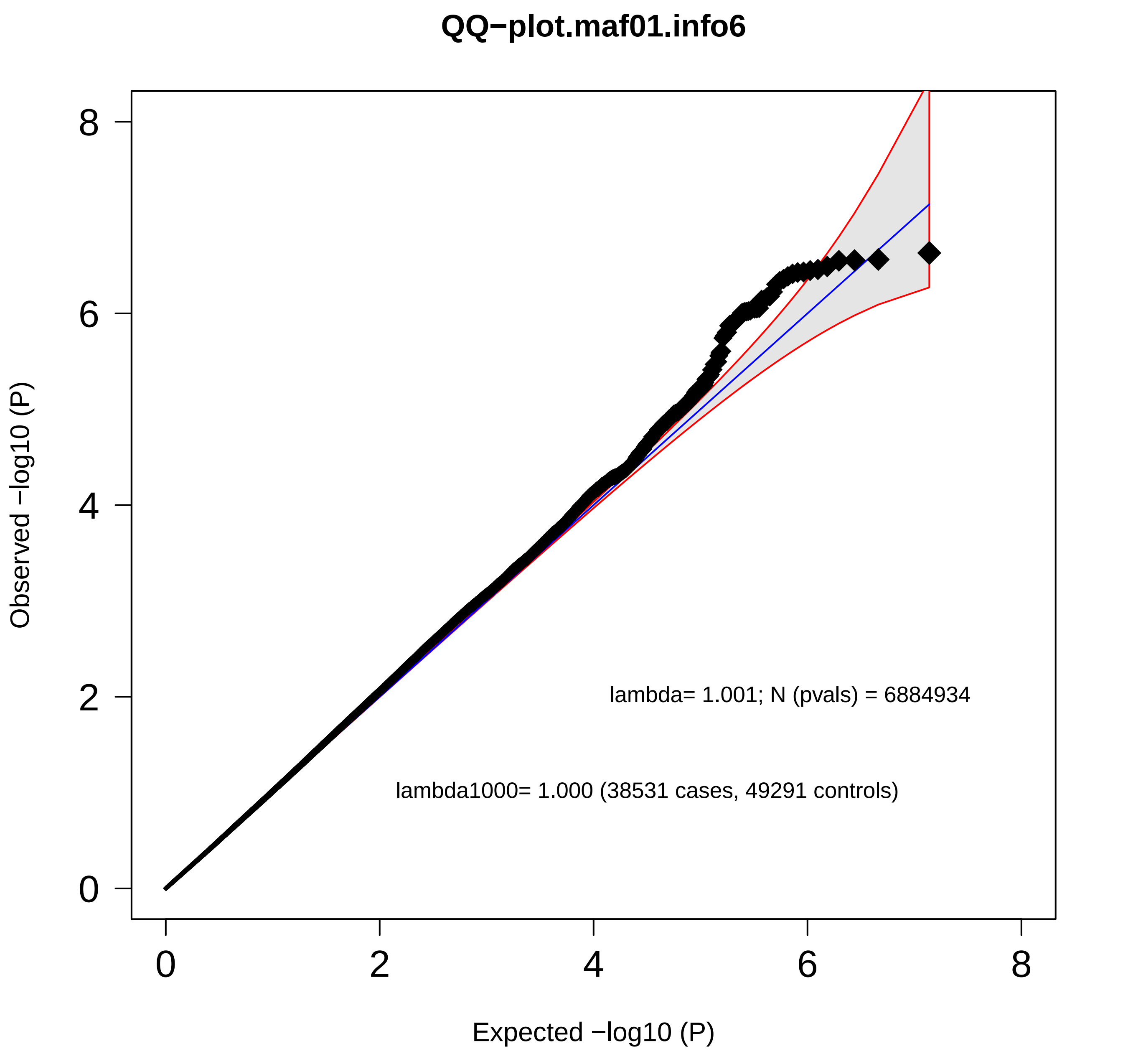

### SF3d.qq.CCGWAS.pdf

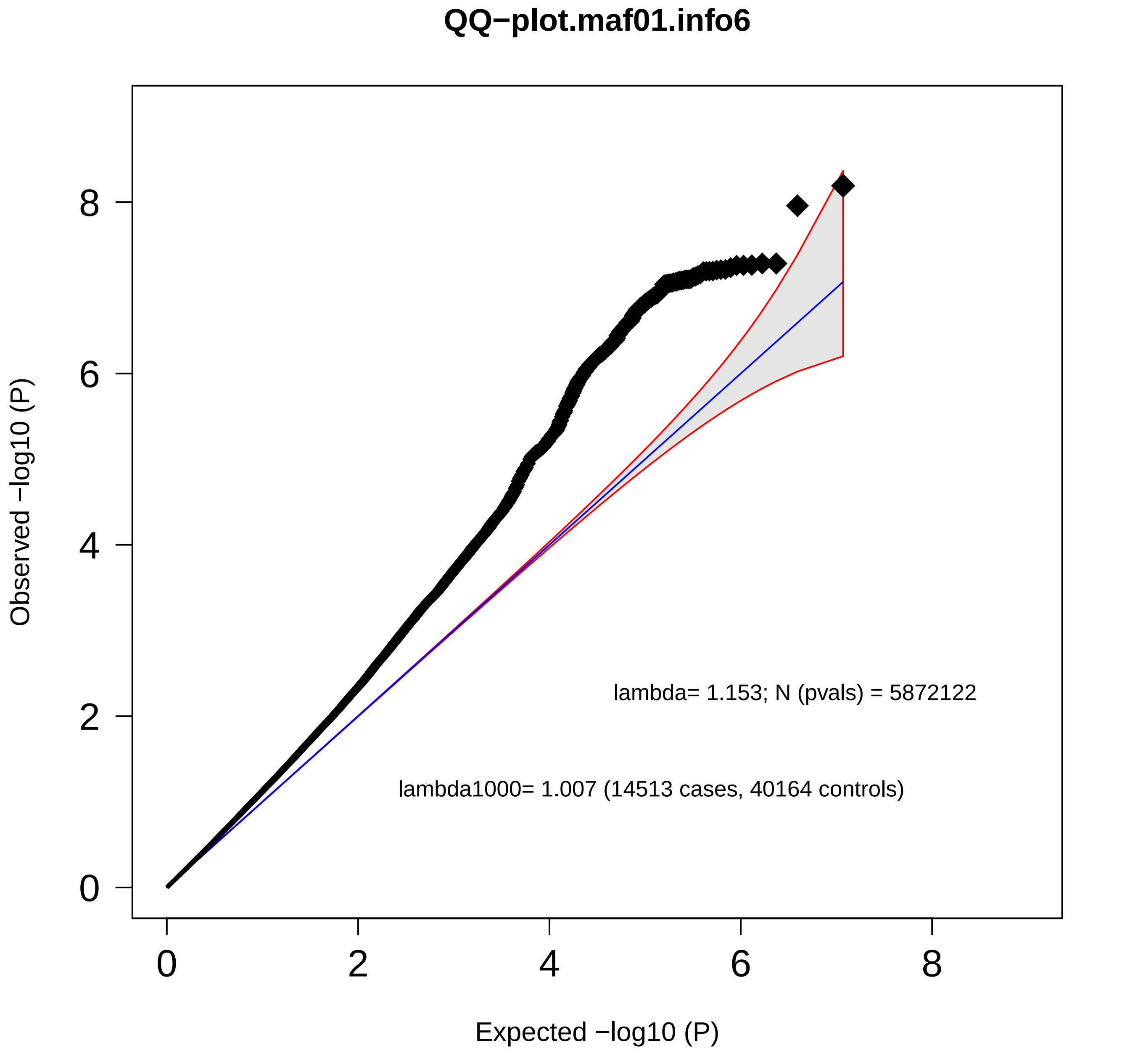

### SF4a.area_plots.BDvsMDD.pdf

#### BIP–MDD_13_run2.2.chr3

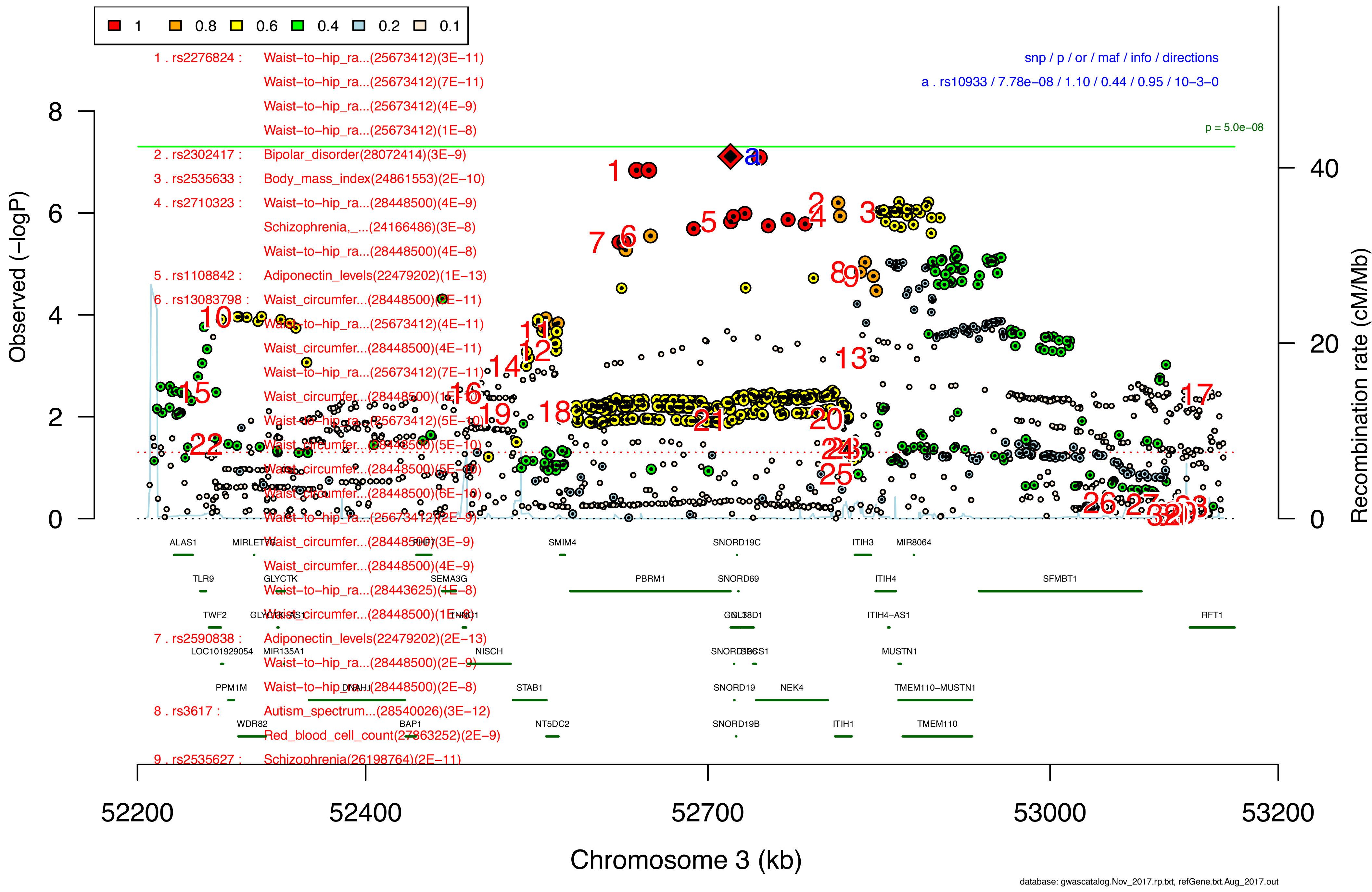

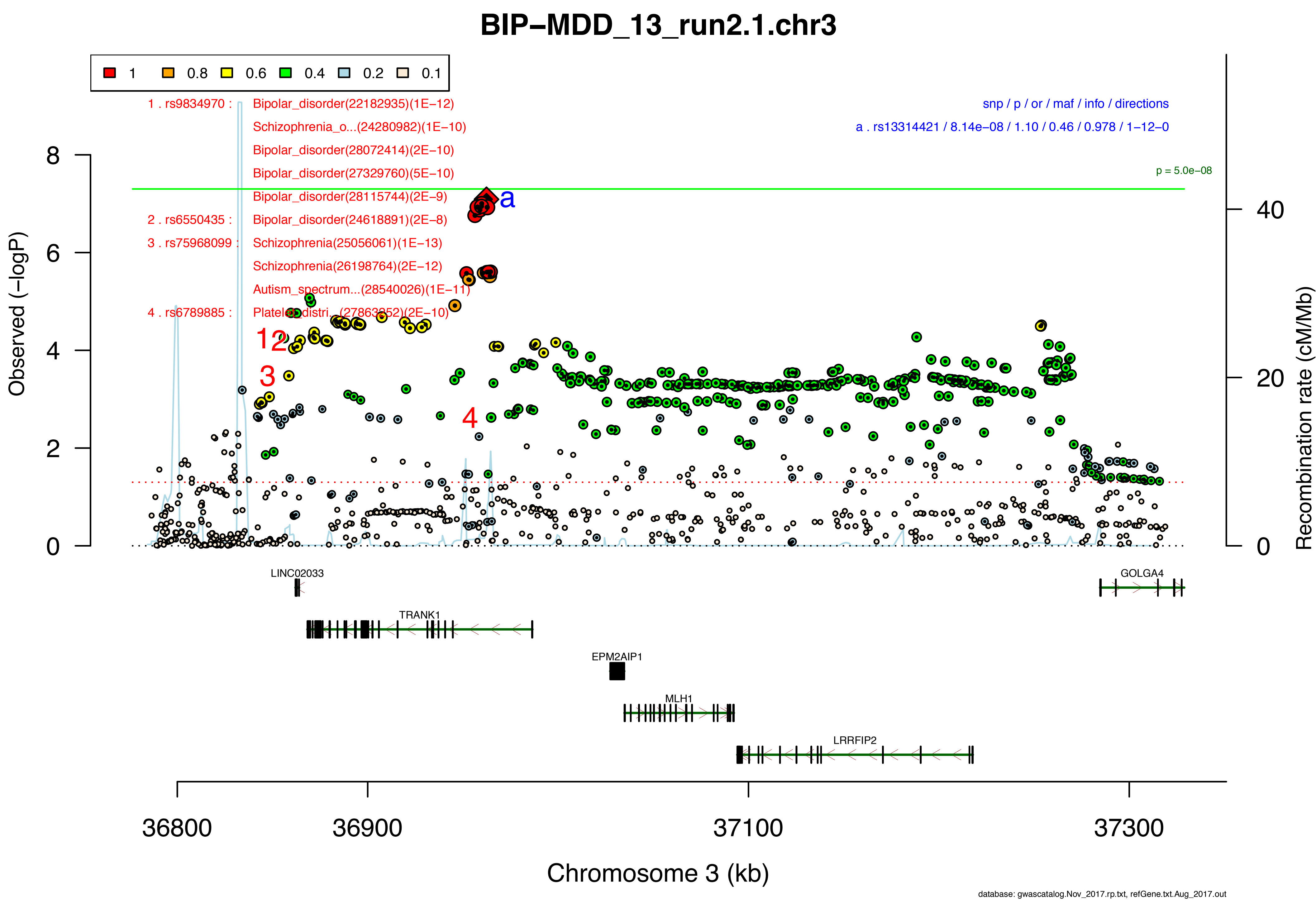

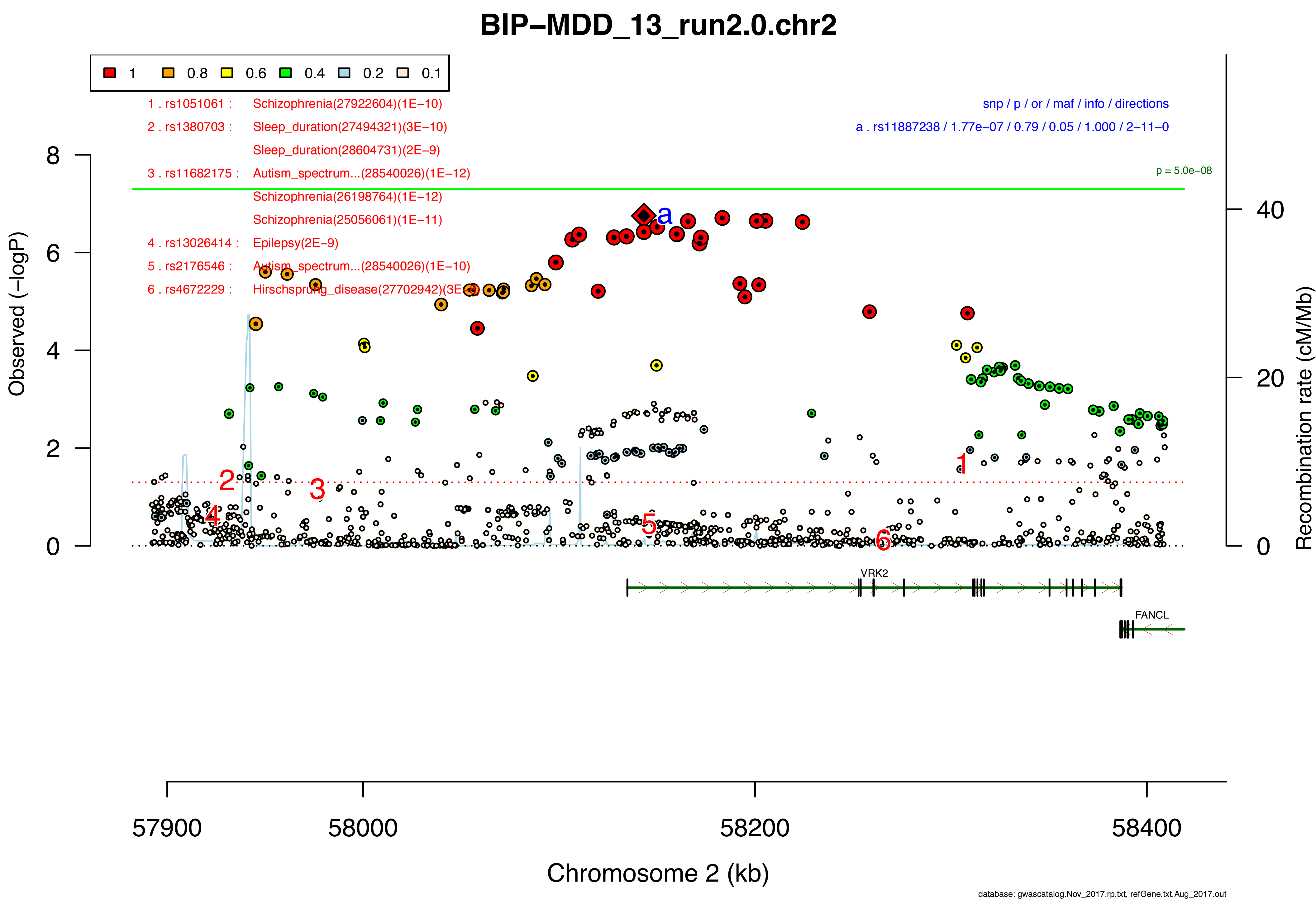

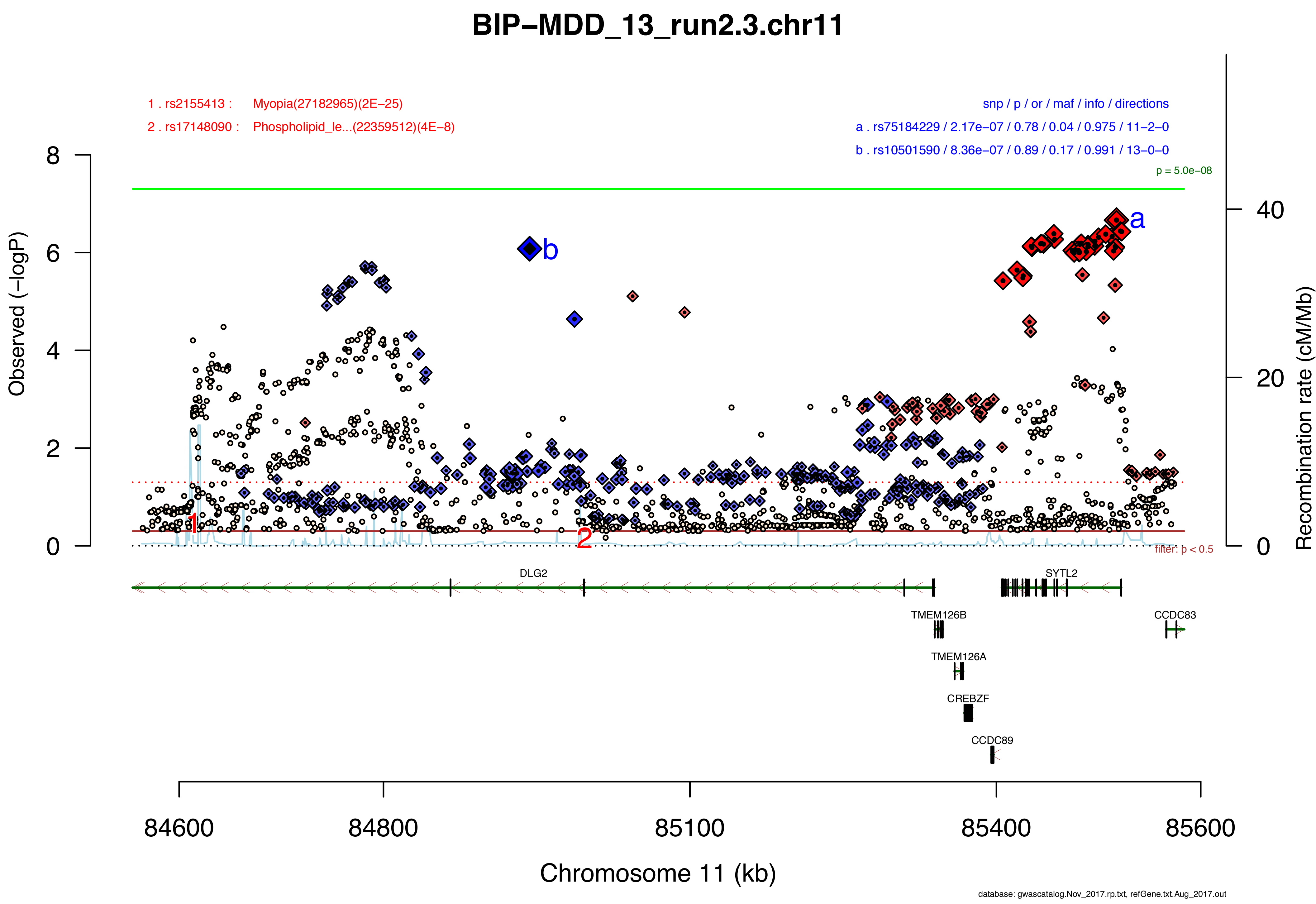

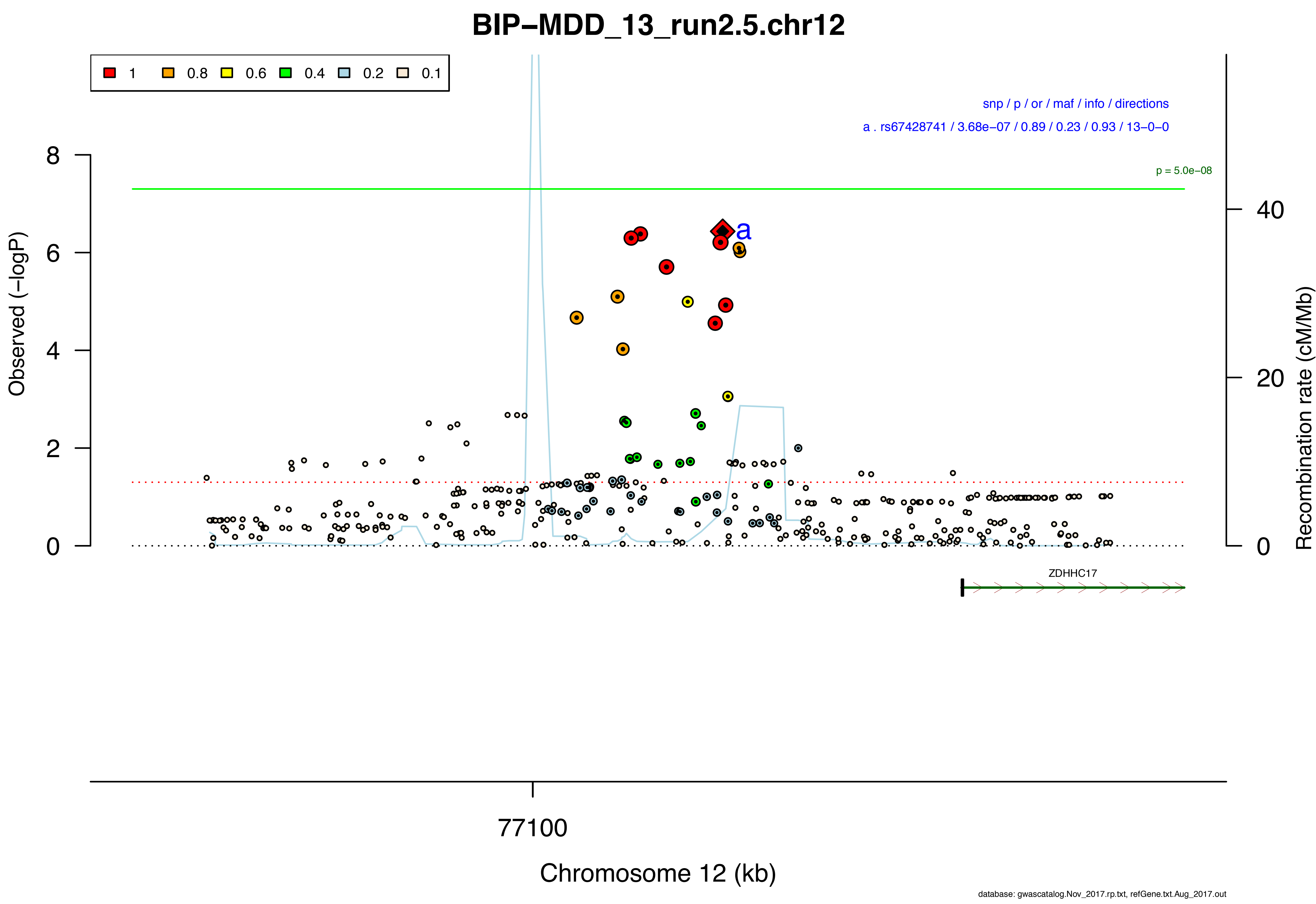

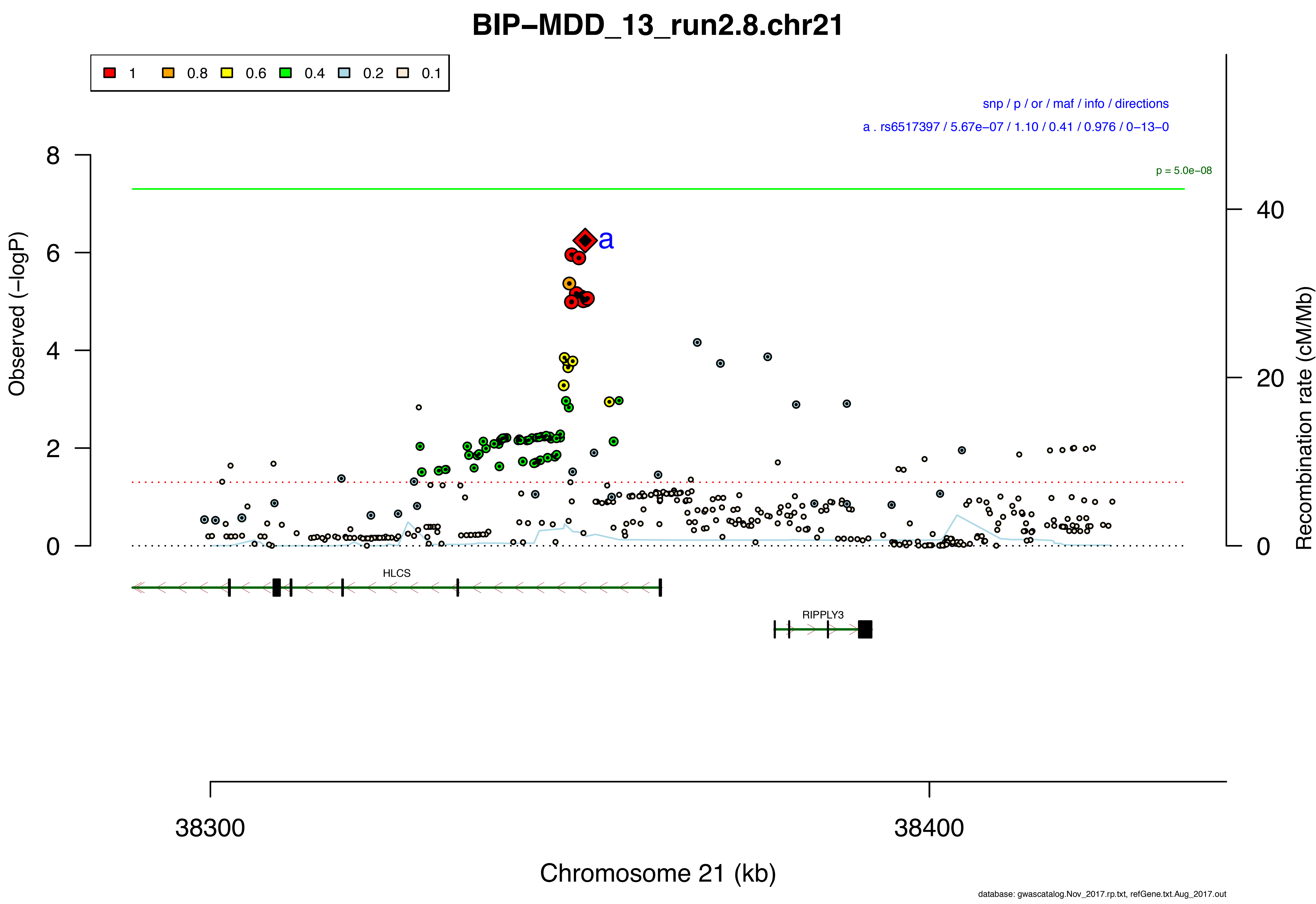

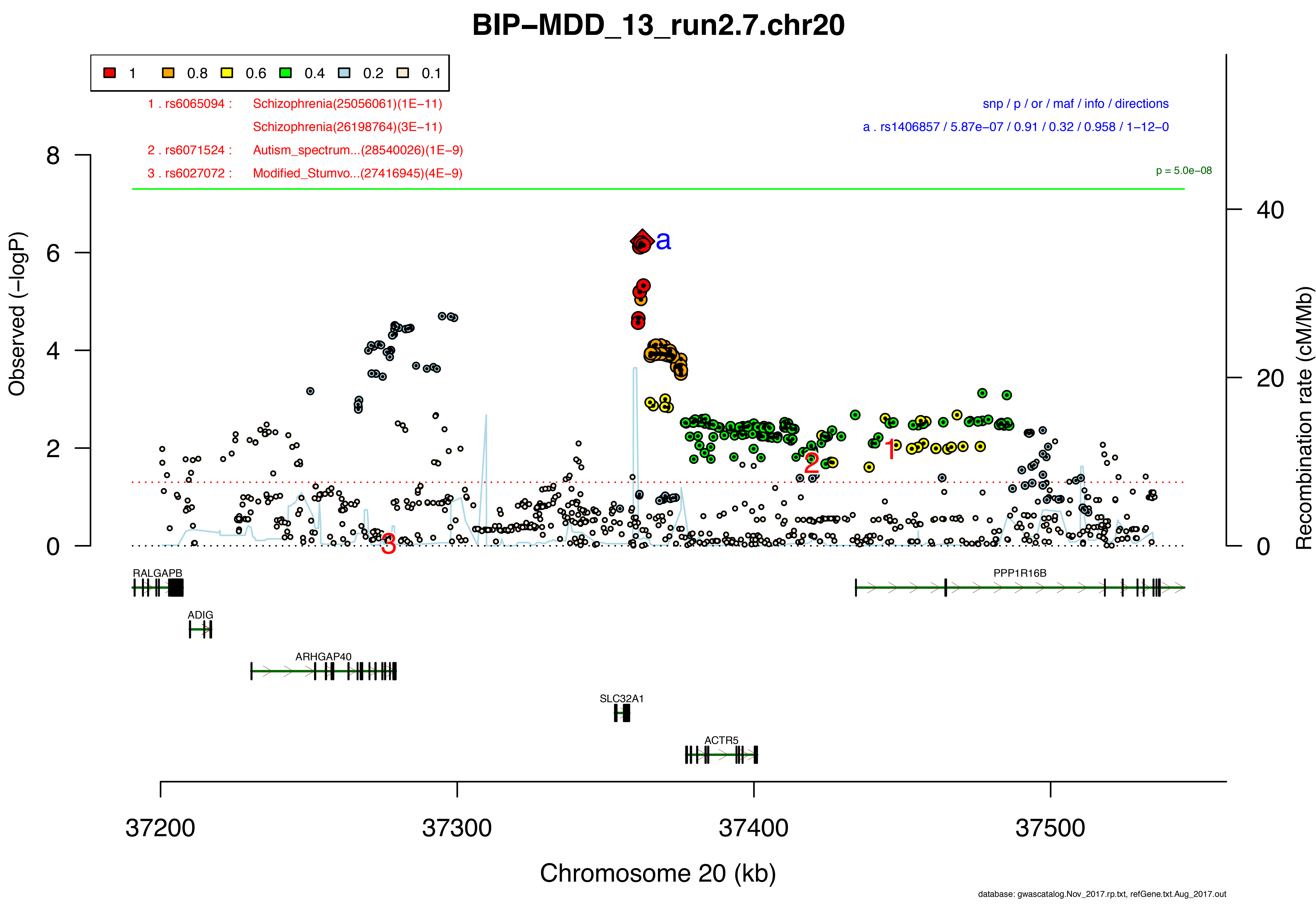

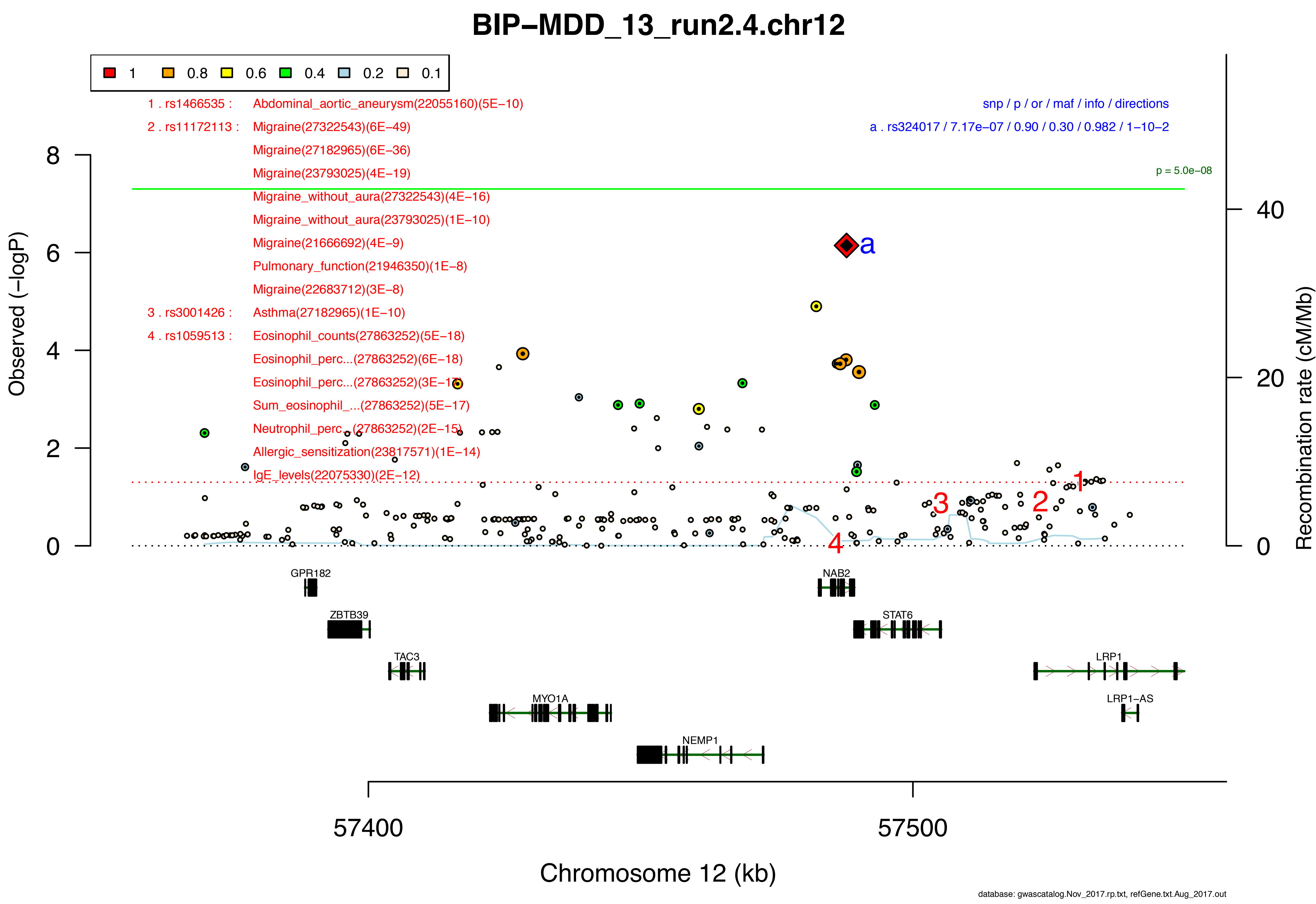

### SF4b.area_plots.MetaRegr.pdf

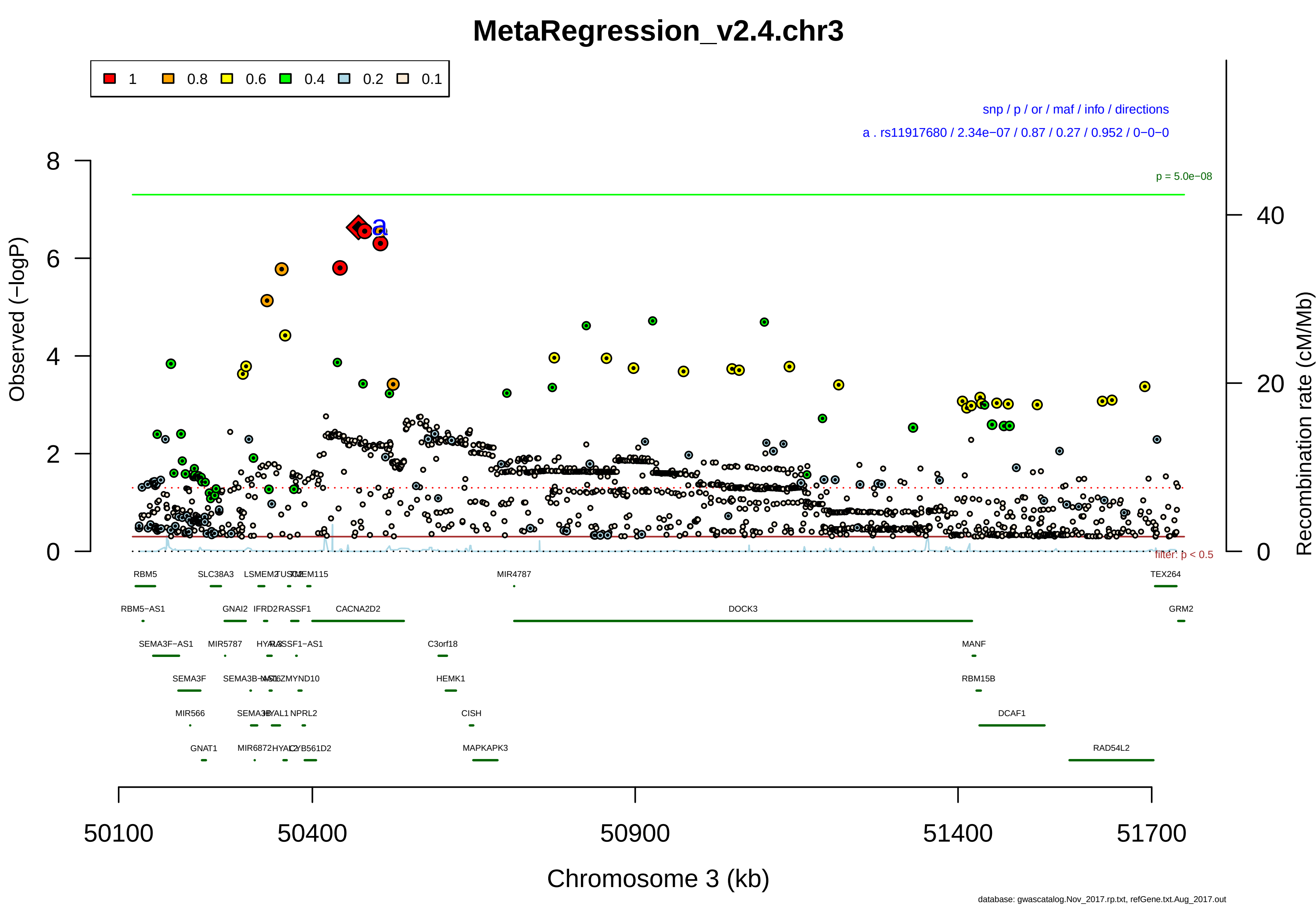

#### MetaRegression_v2.6.chr3

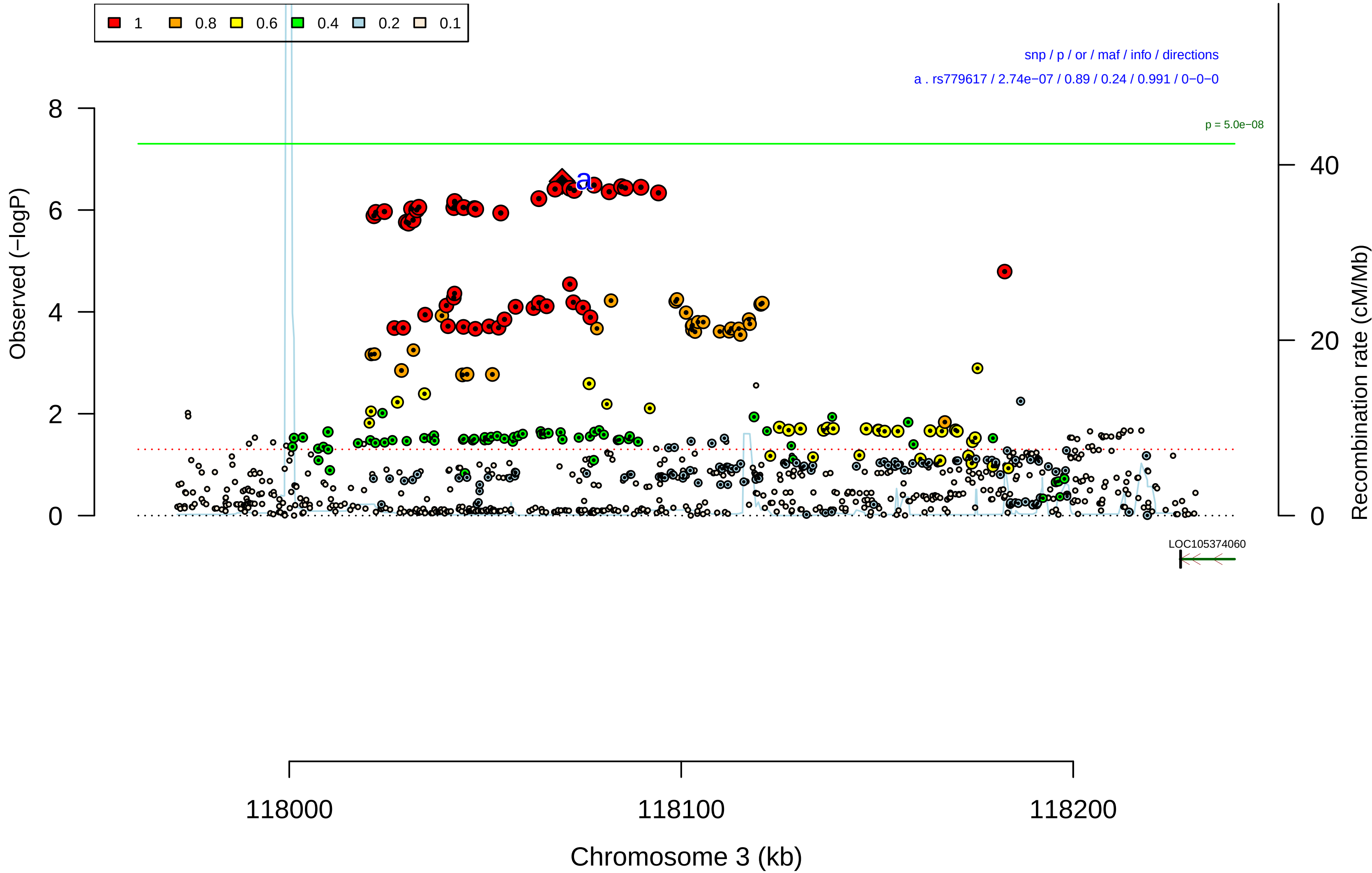


database: gwascatalog.Nov_2017.rp.txt, refGene.txt.Aug_2017.out

#### MetaRegression_v2.13.chr6

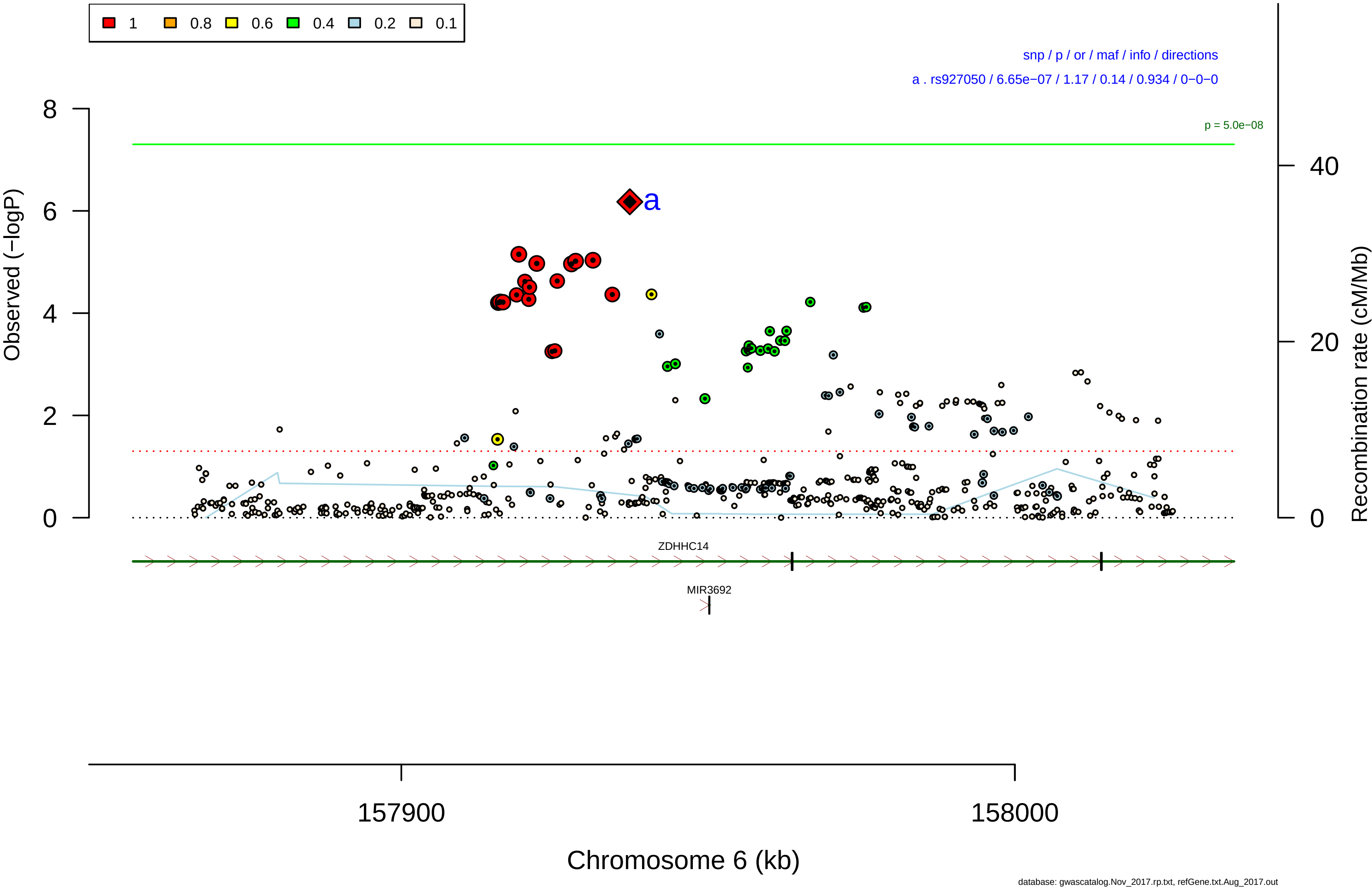

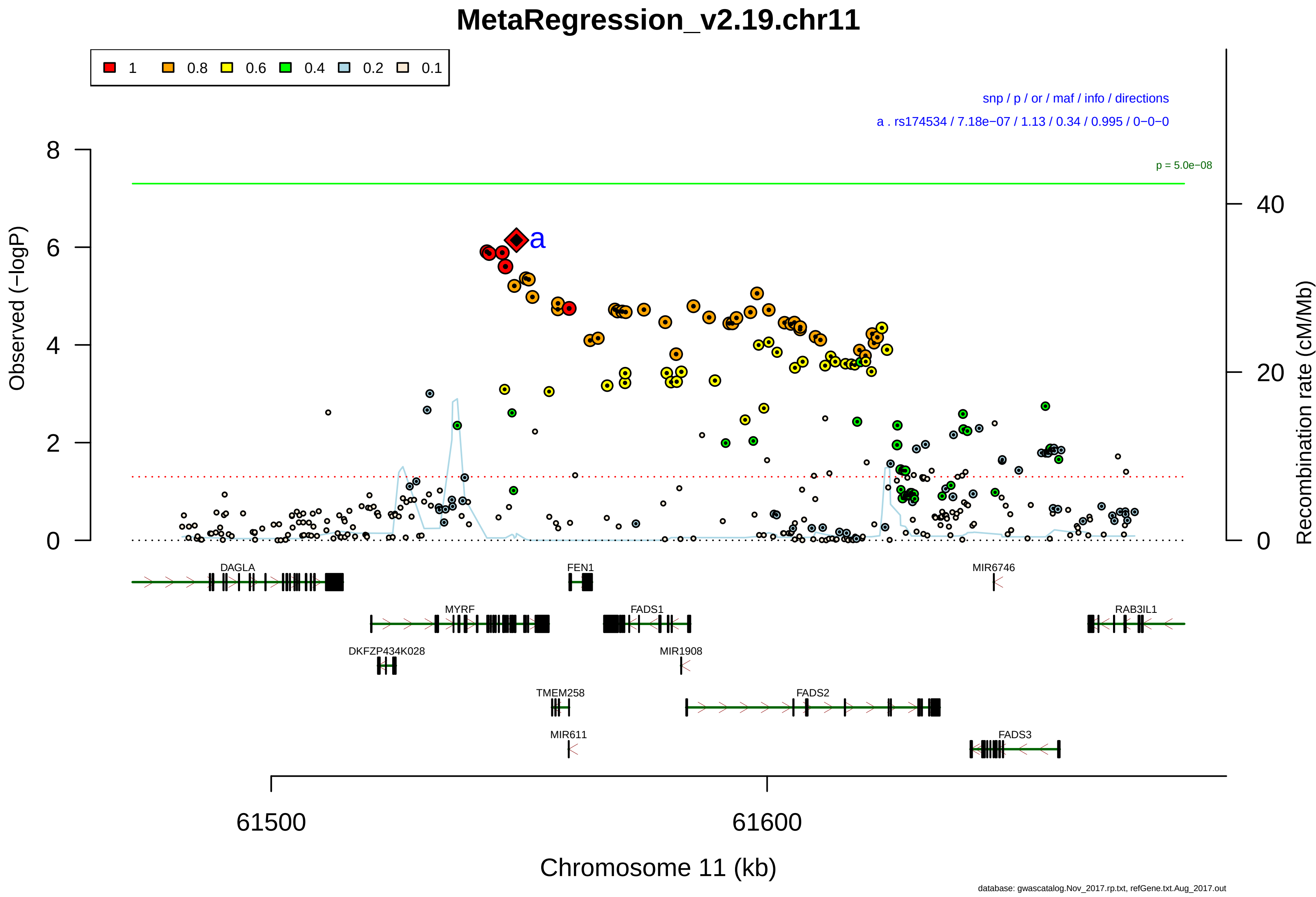

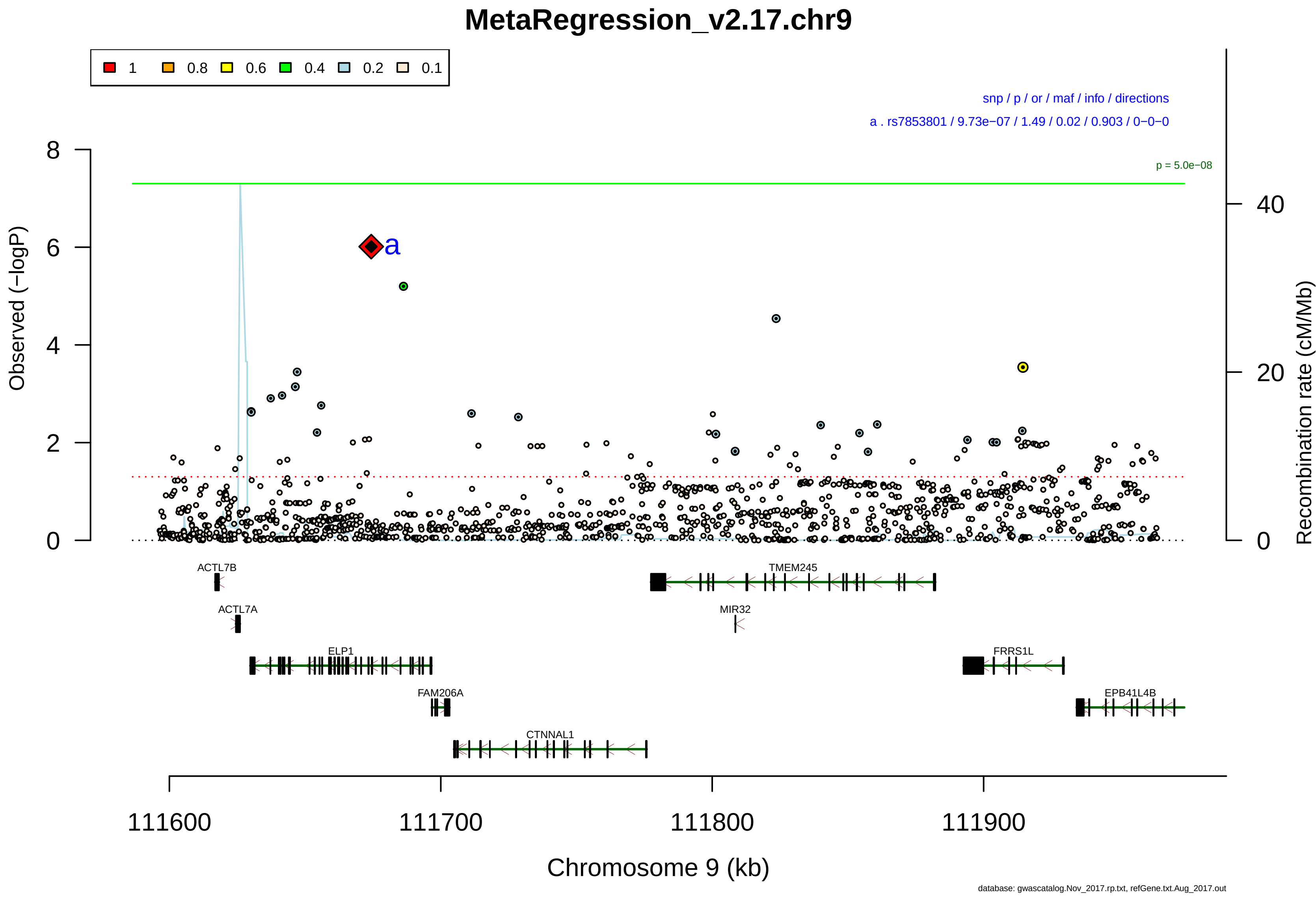

### SF4c.area_plots.CCGWAS.pdf

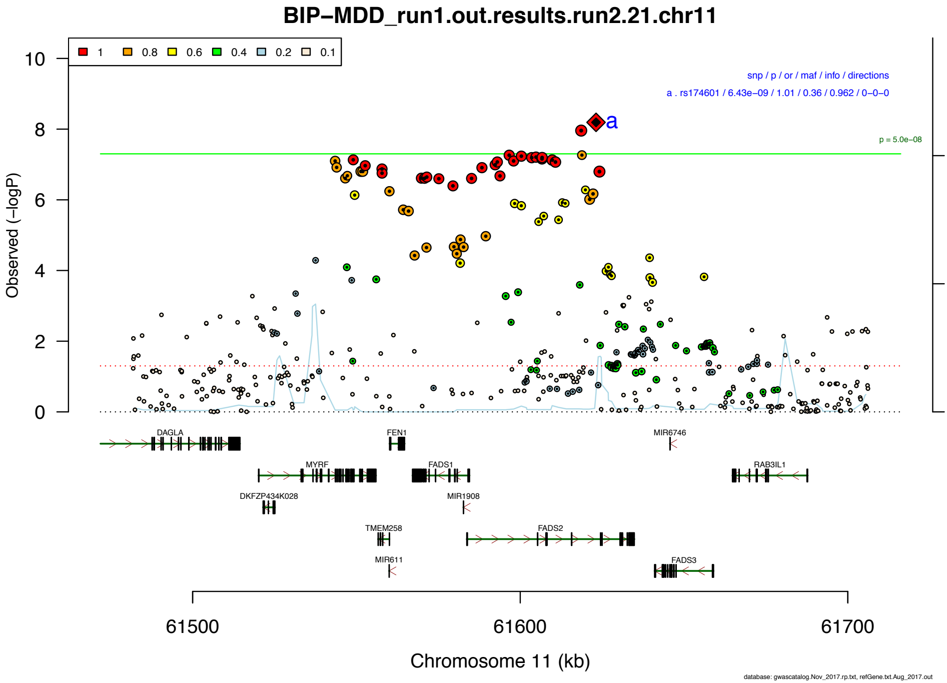

### SF6A.BD-MDD_PRS-SBayesR_BDvsMDD-BD-MDD_R2_errorbars_200323.png

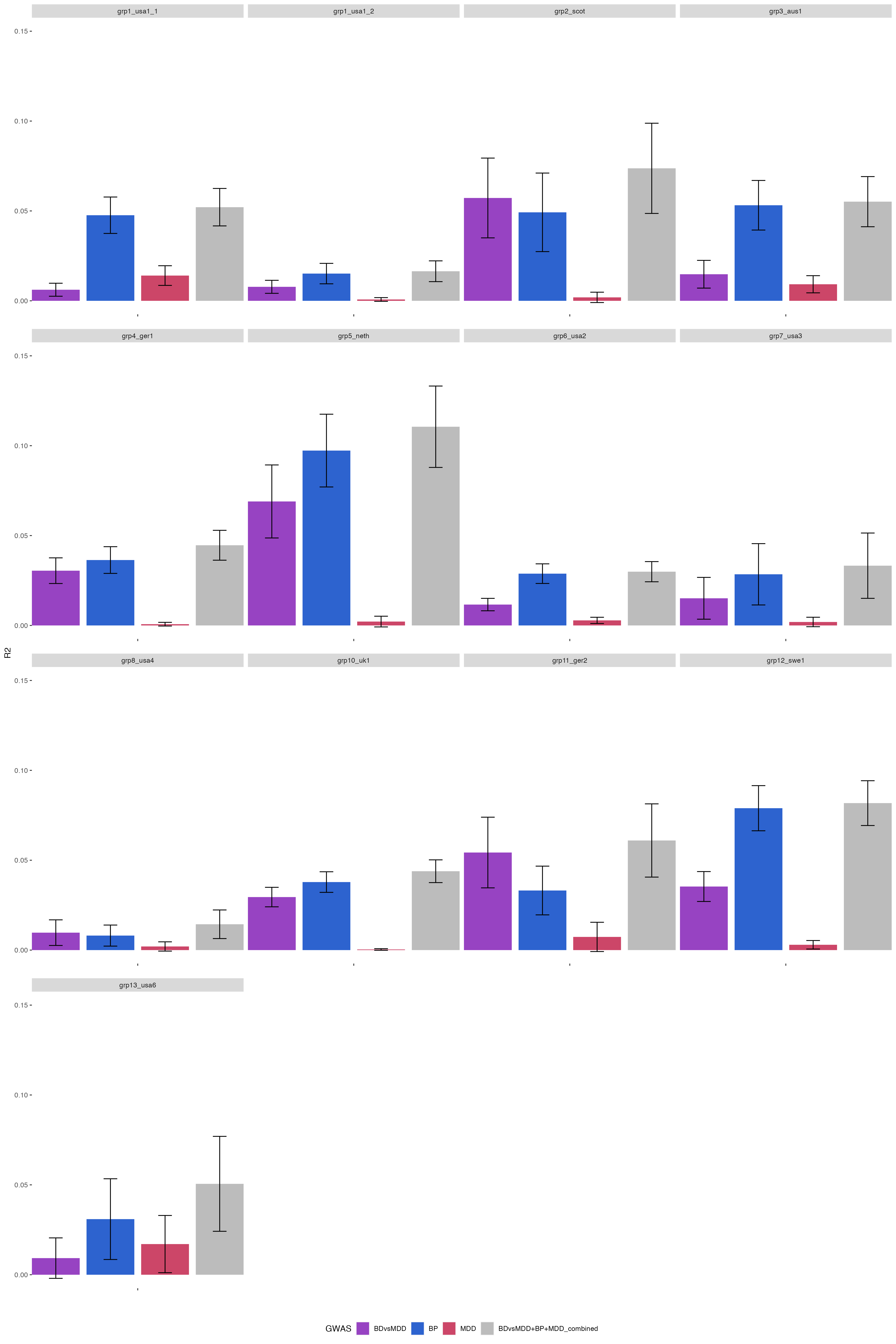

### SF6B.BD-MDD_PRS-SBayesR_BDvsMDD-BD-MDD_BD-D_R2_errorbars_200323.png

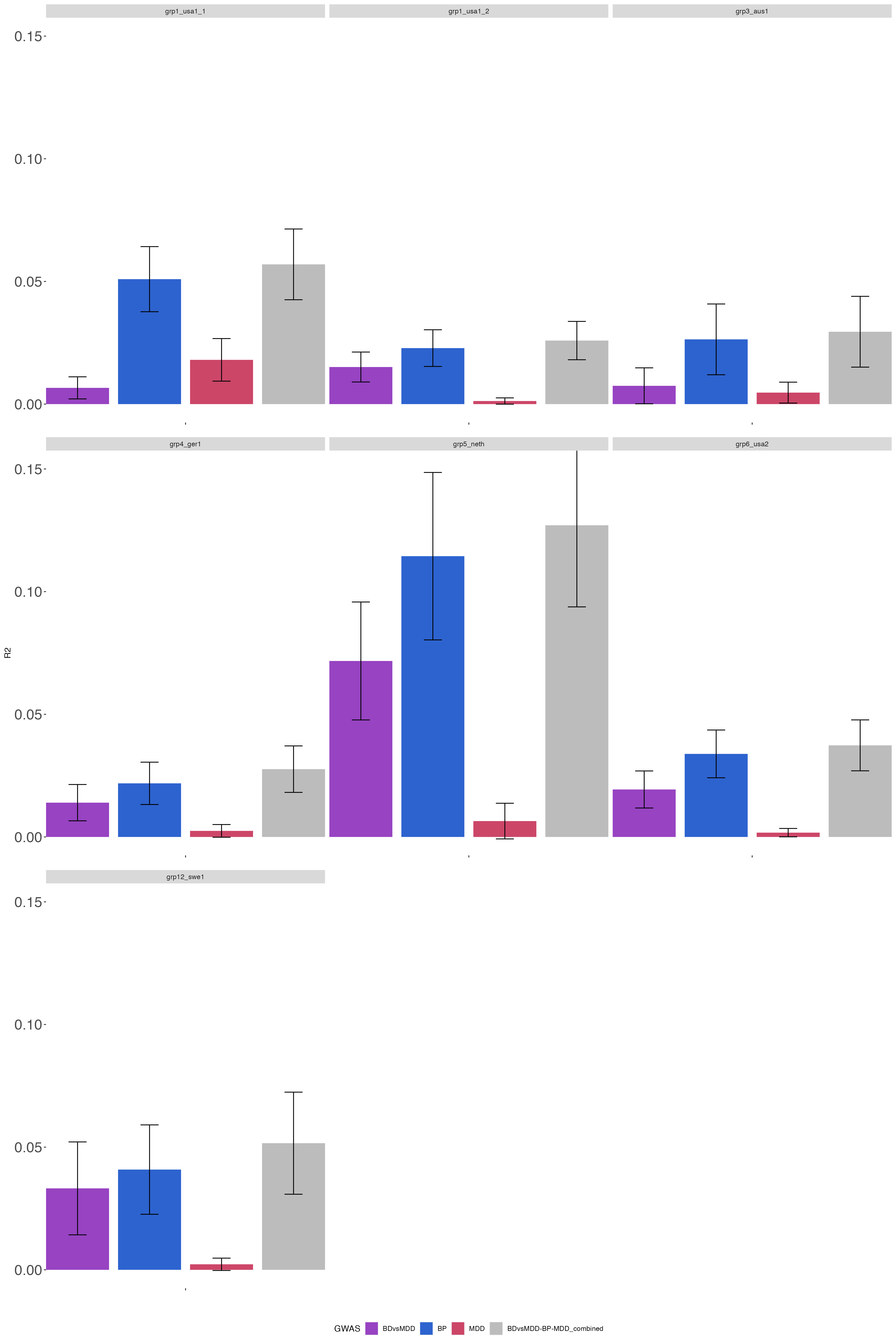

### SF7A.BDvsMDD_PRS-SBayesR_BD-BDvsMDD-BDsubN_AUC_errorbars_200323.pdf

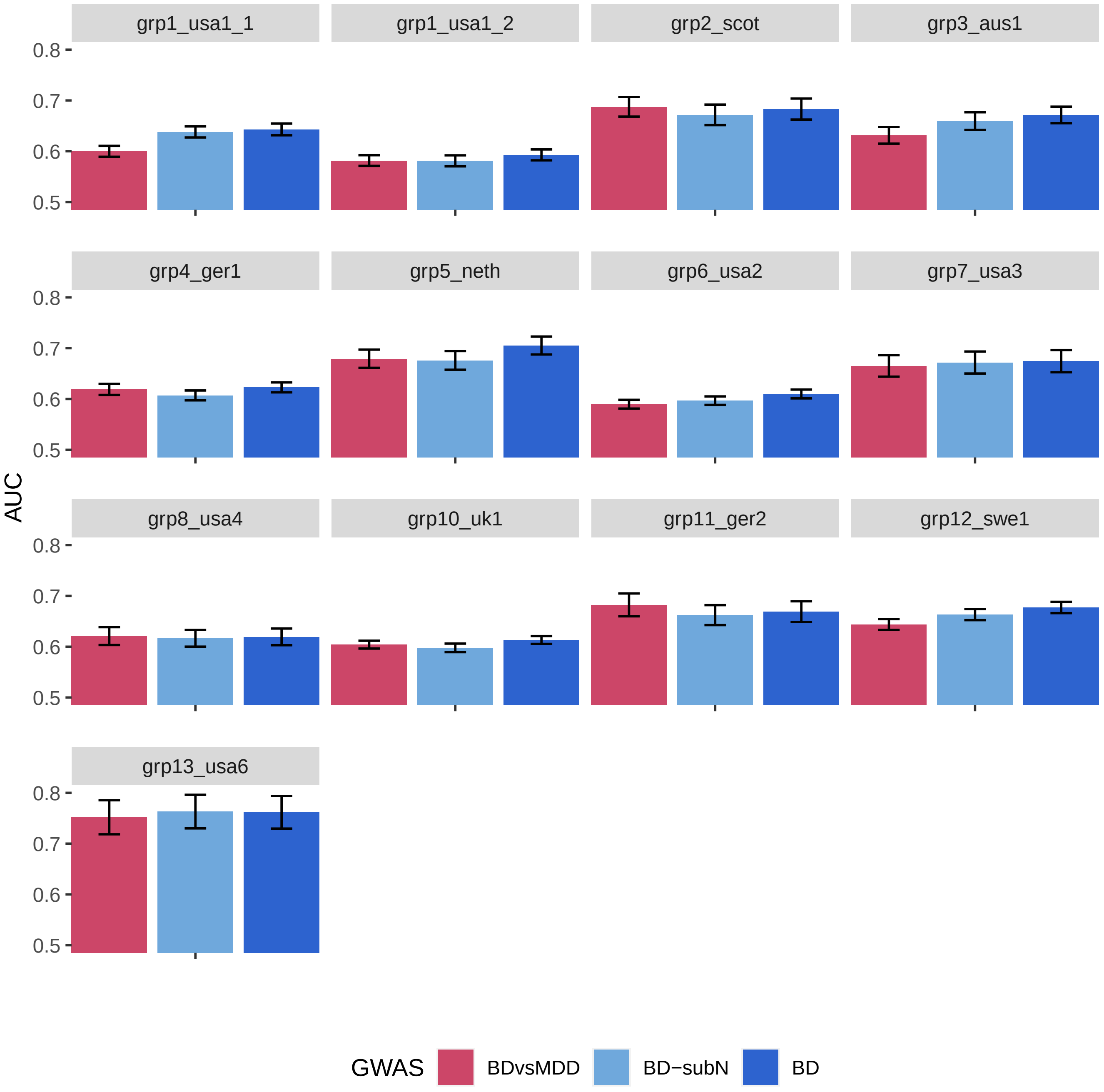

### SF7B.BD-MDD_PRS-SBayesR_BD-MDD-BDsubN_R2_errorbars_200323.png

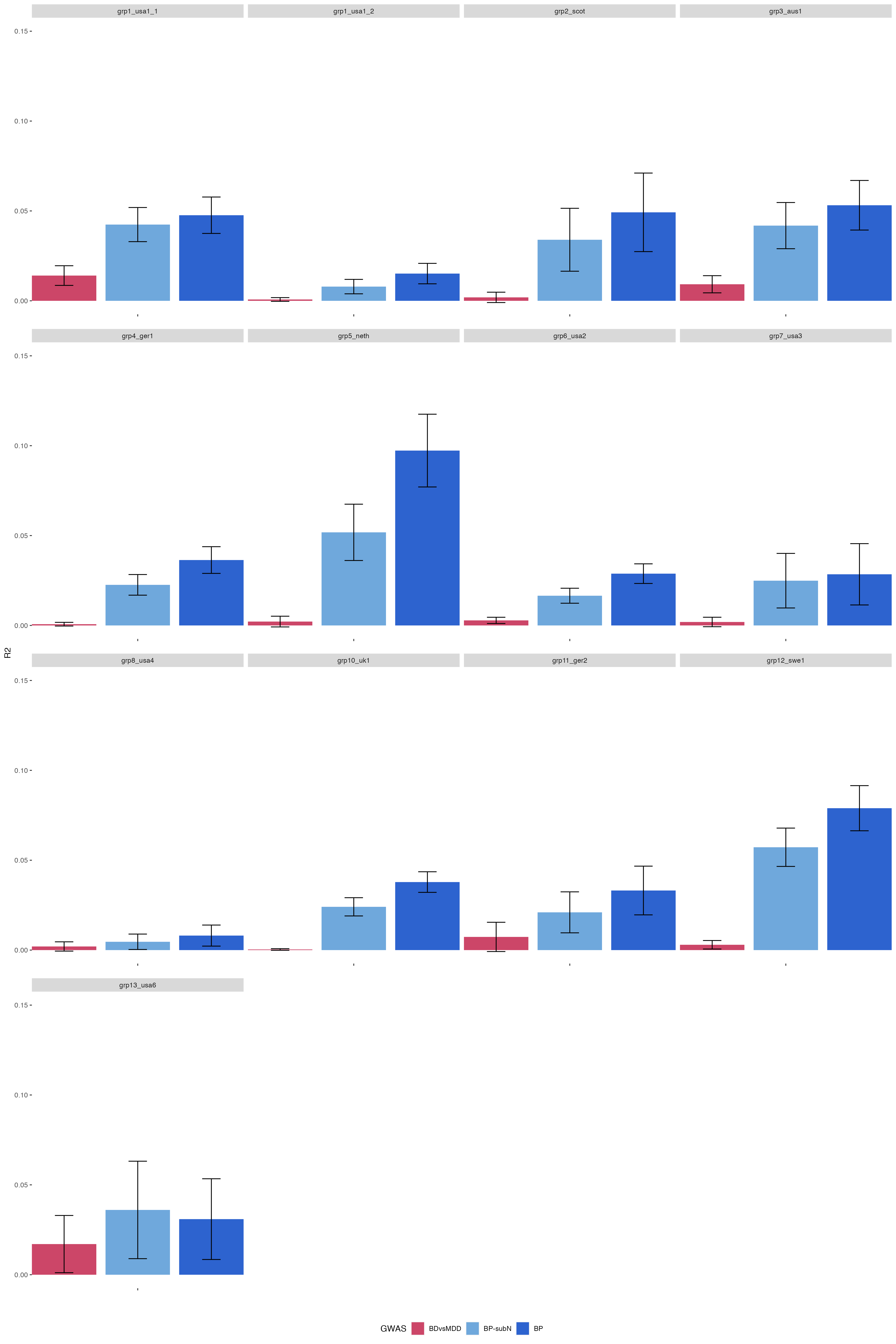

### SF7C.BD-DvsMDD_PRS-SBayesR_BD-MDD-BDsubN_AUC_errorbars_200323.pdf

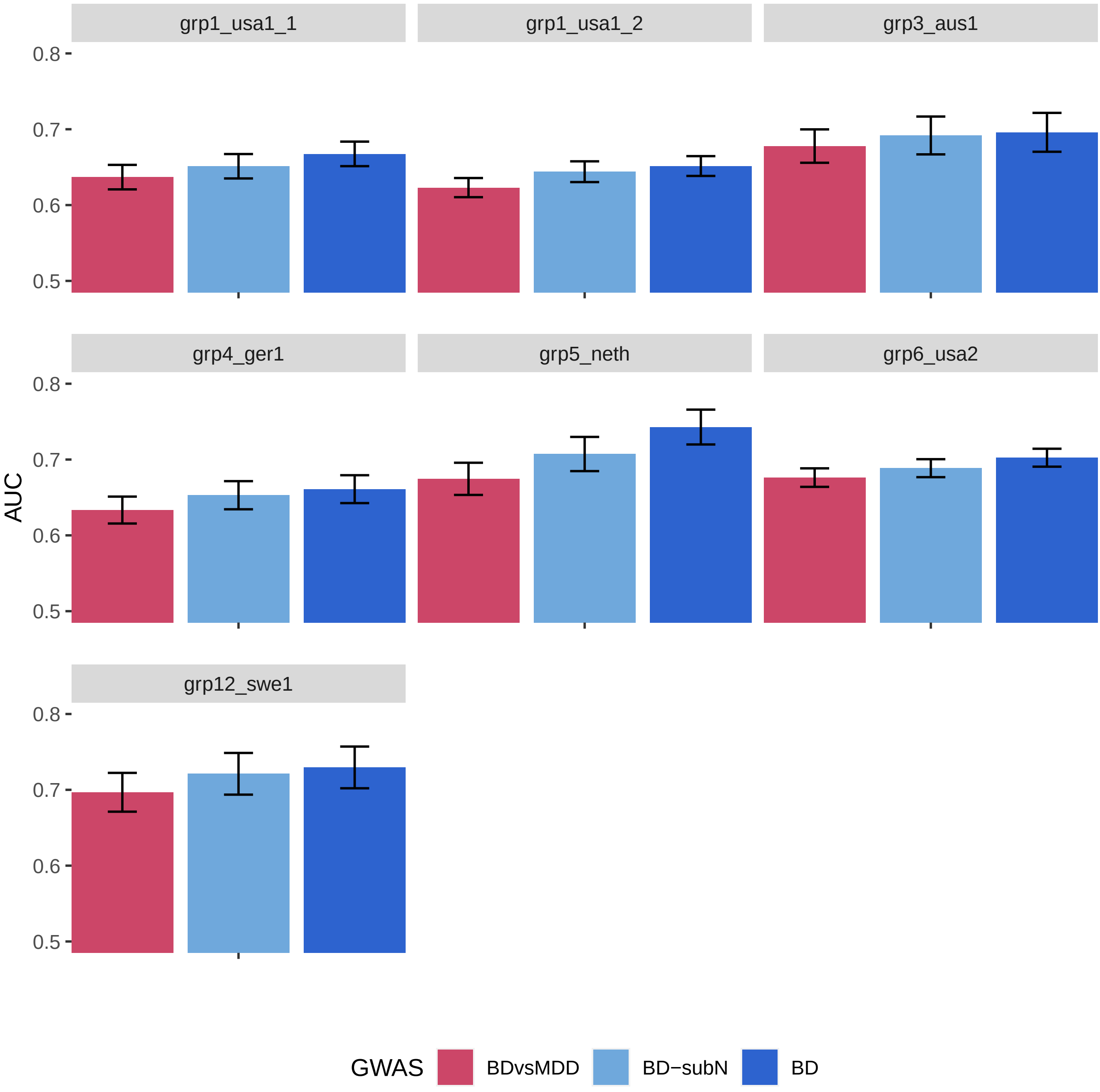

### SF7D.BD-D-MDD_PRS-SBayesR_BD-MDD-BDsubN_BD-D_R2_errorbars_200323.png

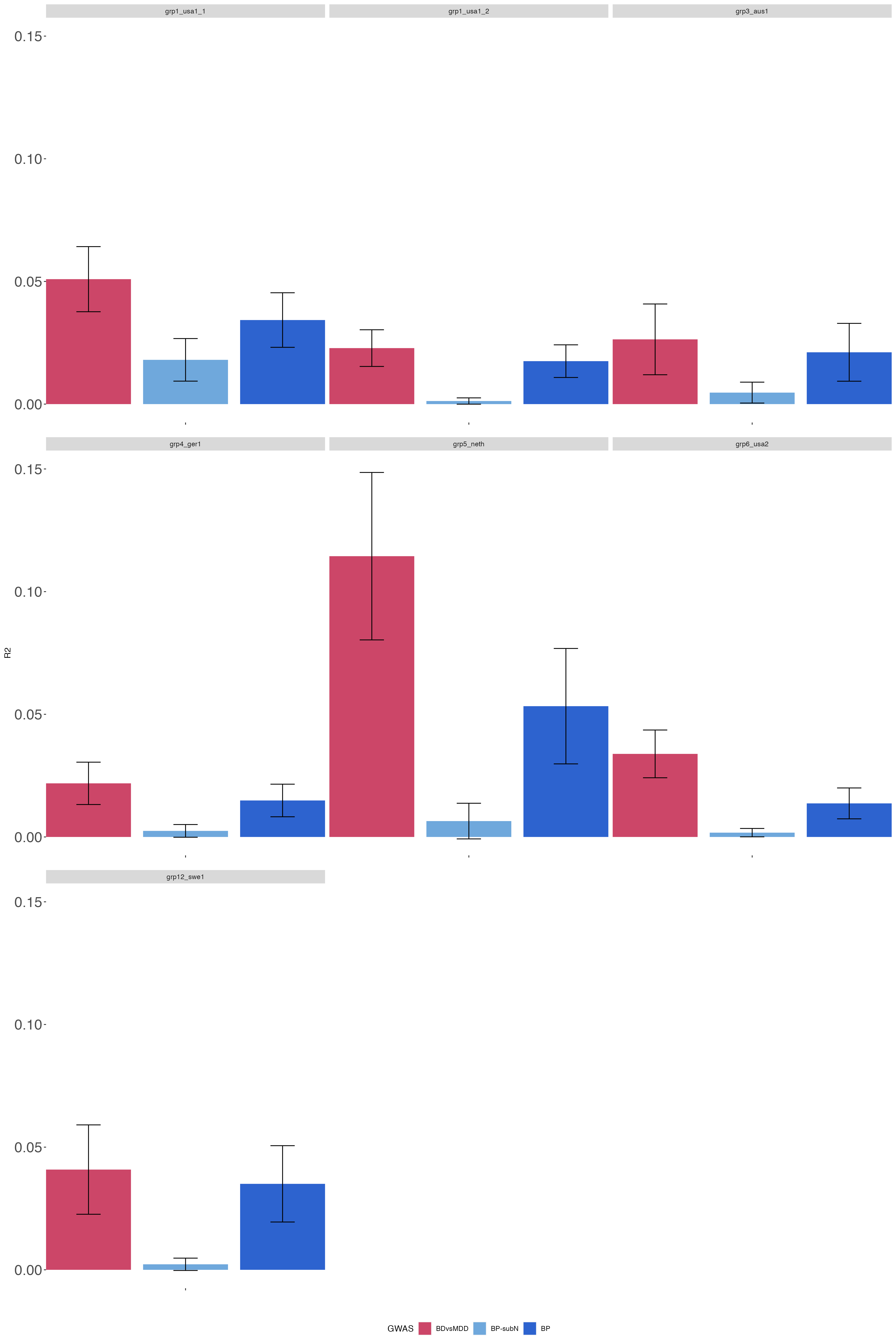

### SF8.grp5_neth.R2_all.with_BIP2019.SBayesR.pdf

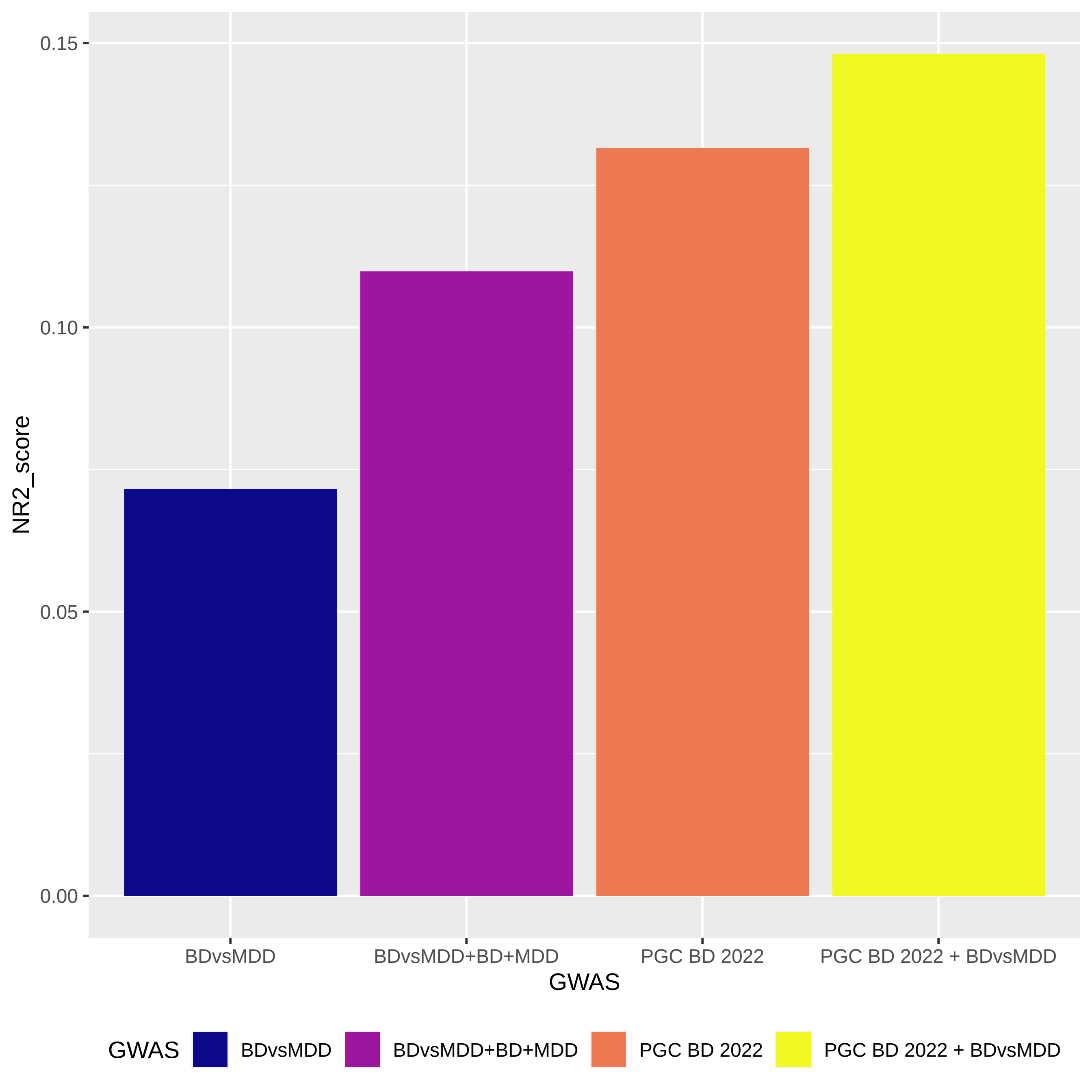

### SF9A.BDvsMDD_PRS-SBayesR_SI_AUC_errorbars_200323.pdf

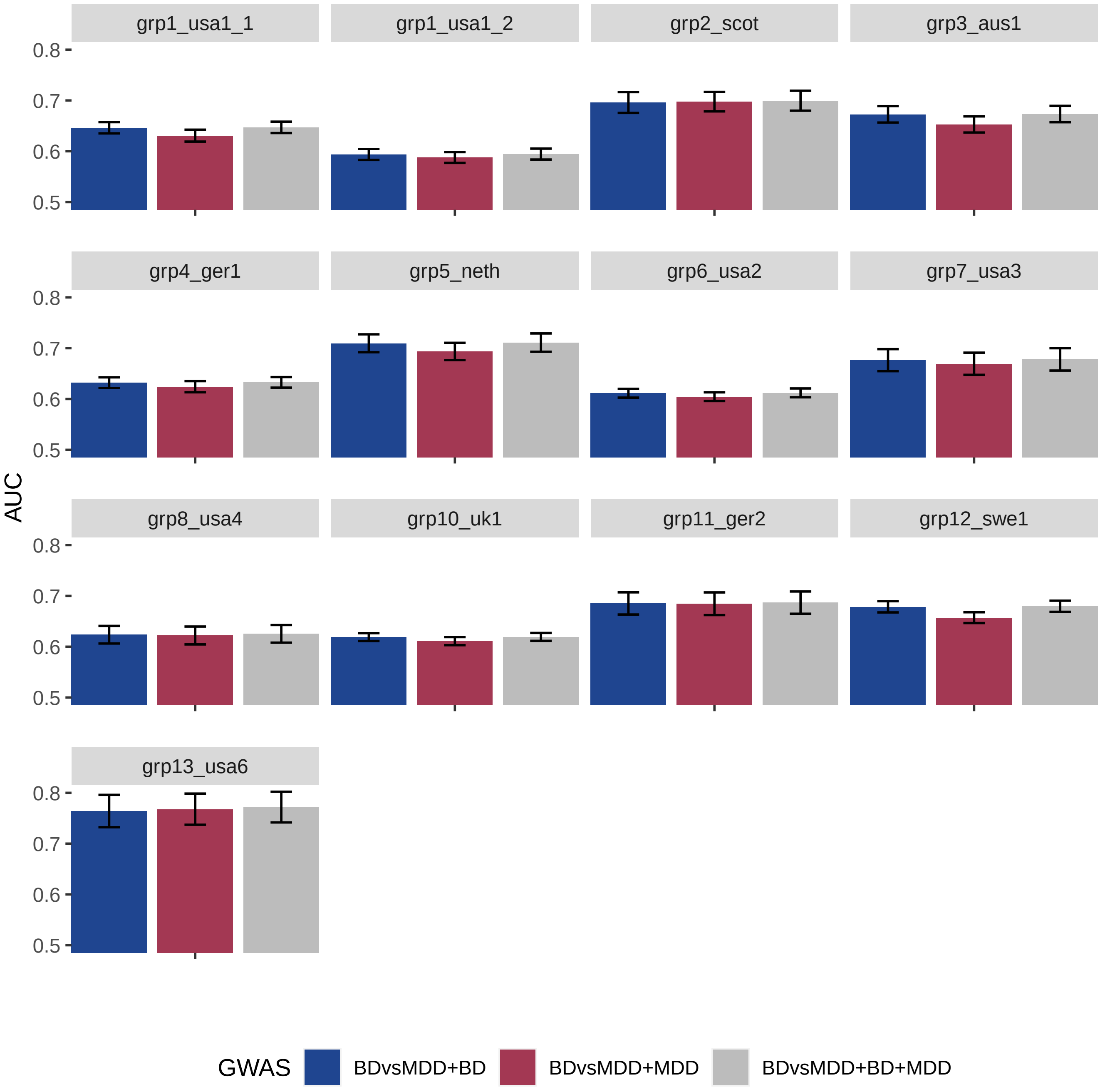

### SF9B.BD-MDD_PRS-SBayesR_SI_R2_errorbars_200323.png

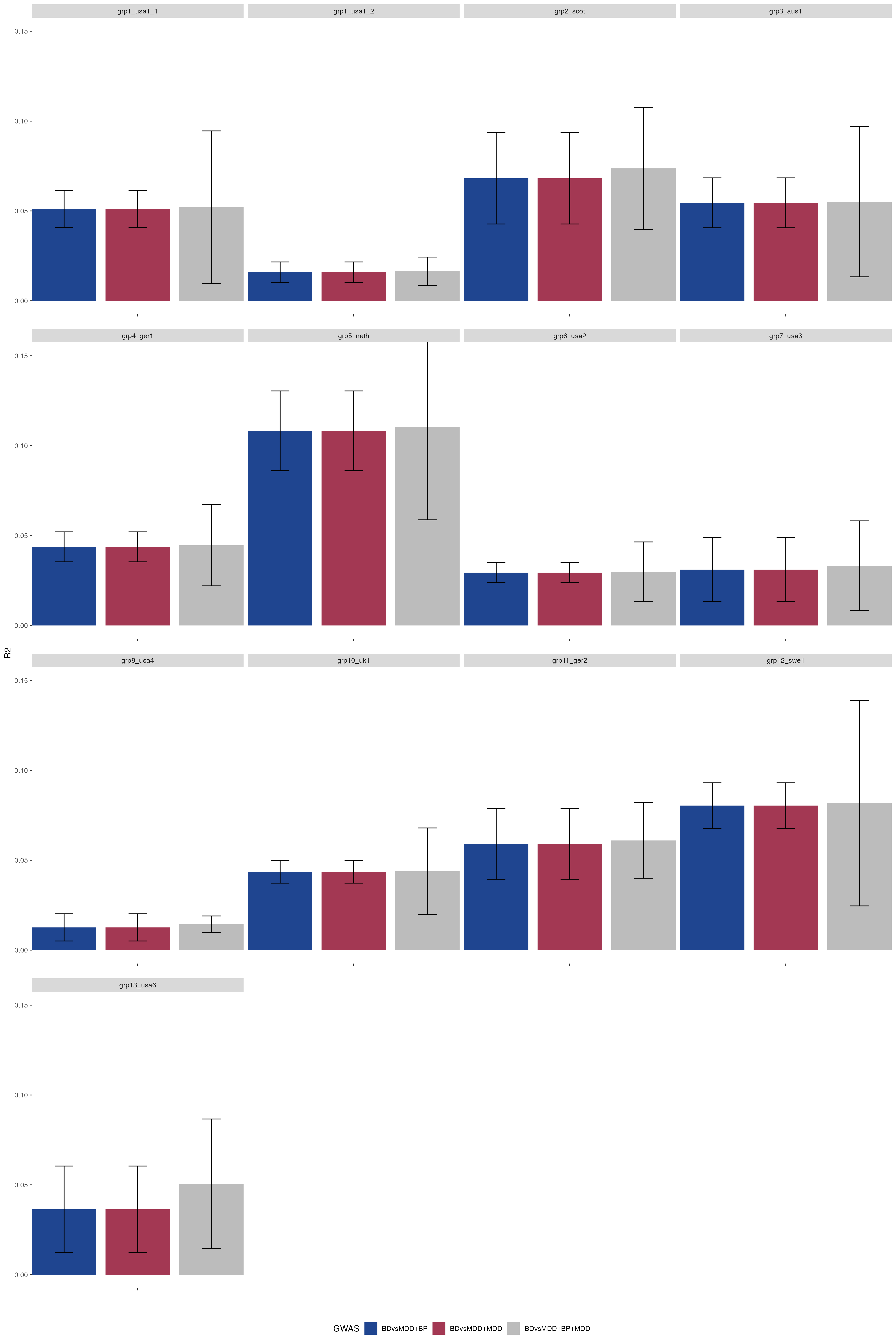

### SF9C.BD-DvsMDD_PRS-SBayesR_SI_AUC_errorbars_200323.pdf

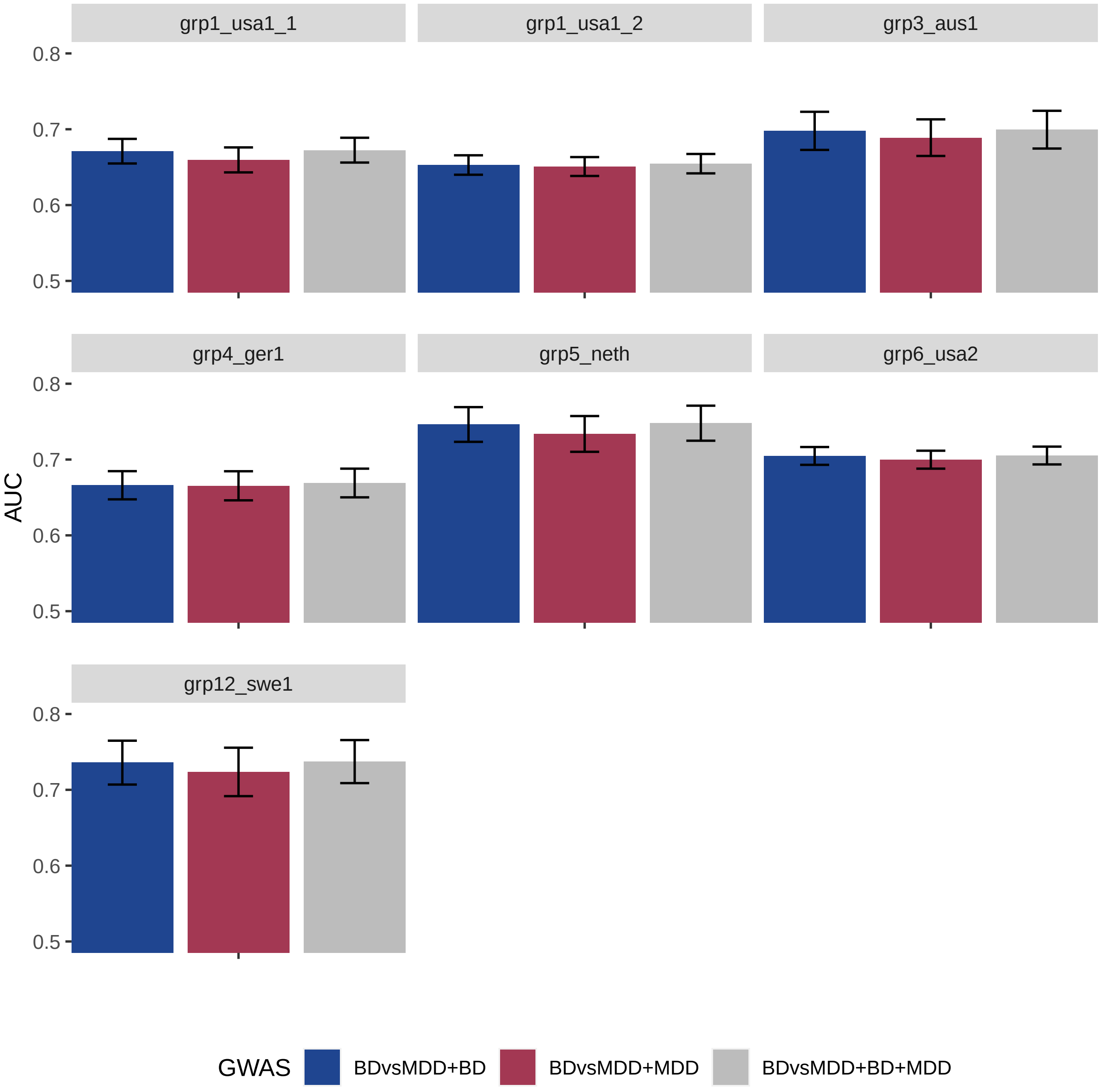

### SF9D.BD-DvsMDD_PRS-SBayesR_SI_R2_errorbars_200323.png

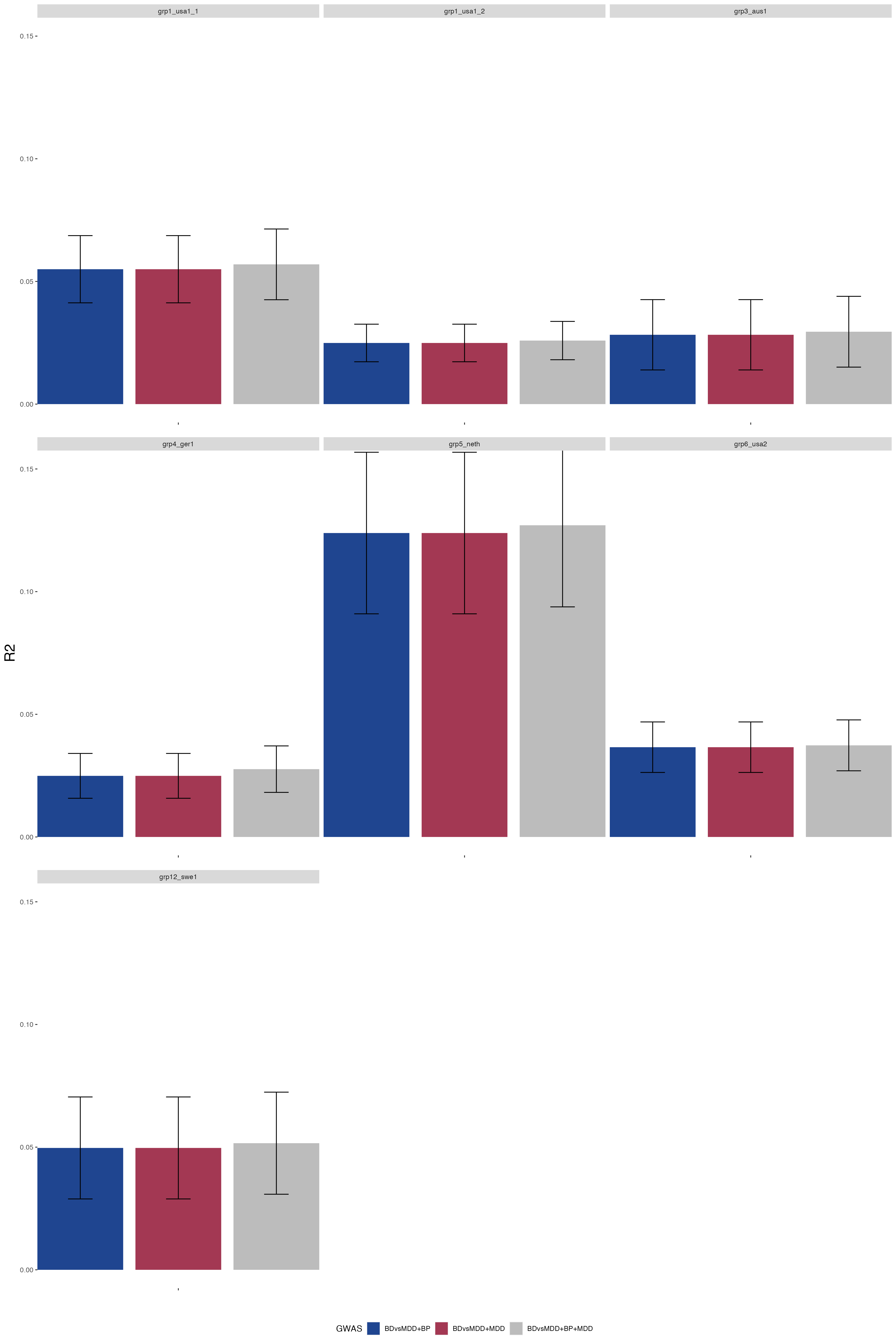
